# Leveraging perturbations to infer the population dynamics of human rhinovirus and interaction of influenza A virus

**DOI:** 10.64898/2026.03.23.26348908

**Authors:** Wakinyan Benhamou, Emily Howerton, Sang Woo Park, Ćecile Viboud, C. Jessica E. Metcalf, Bryan T. Grenfell

**Affiliations:** Department of Ecology and Evolutionary Biology, Princeton University, Princeton, NJ, USA; High Meadows Environmental Institute, Princeton University, Princeton, NJ, USA; School of Biological Sciences, Seoul National University, Seoul, Korea; Institute for Data Innovation in Science, Seoul National University, Seoul, Korea; Fogarty International Center, National Institutes of Health, Bethesda, MD, USA; Princeton School of Public and International Affairs, Princeton University, Princeton, NJ, USA

**Keywords:** Rhinovirus, Influenza A virus, Population dynamics, Virus-virus interaction, Viral interference, Non-pharmaceutical interventions, Bayesian inference

## Abstract

Many respiratory pathogens co-circulate within human populations. Yet, how pathogen community structure shapes the dynamics of infectious diseases remains poorly understood. At the population level, investigating polymicrobial dynamics, with potential underlying competitive or cooperative interactions, is challenging, because of confounding factors such as differing seasonality. This is particularly true for endemic pathogens which typically exhibit stable periodic dynamics. Their disruption due to the implementation of non-pharmaceutical interventions during the COVID-19 pandemic thus represents a unique large-scale natural experiment that can be leveraged to provide valuable insights into the complex interplay between respiratory pathogens. Here, we focus on the population dynamics of human rhinovirus (common cold) and on the potential viral interference of influenza A virus (flu A), which is hypothesized to account for their asynchronous circulation. Using a Bayesian framework, we first show based on simulations that exogenous perturbations can be a powerful tool to disentangle the contribution of pathogen interaction from other epidemiological factors. We then apply our framework to surveillance time series from the US and Canada spanning the COVID-19 pandemic. We estimate key parameters of rhinovirus but find no conclusive support for an influence of influenza A virus at the population level.

**Author summary:** A wide range of respiratory viruses (influenza, coronaviruses, respiratory syncytial virus, . . .) circulate in the same communities and can infect the same individuals. Yet, the extent to which these pathogens interact and shape each other’s spread remains under-explored and represents an emerging frontier in public health. We focus here on the dynamics of human rhinovirus, a leading cause of the common cold, particularly among young children. Rhinovirus and influenza A virus typically peak at different times of the year, raising the possibility of some negative interactions. For viruses that peak at consistent times each year, it is however challenging to determine whether differences in their timing are due to true virus-virus interactions or simply reflect their own, independent, seasonal trends. We develop a mathematical modeling approach that leverages the disruption of these dynamics due to the COVID-19 pandemic as a large-scale natural experiment. Our work demonstrates how such epidemiological perturbations can be used to uncover interactions among co-circulating pathogens. Using multi-year surveillance incidence data from the United States and Canada, we estimate key parameters governing rhinovirus transmission and immunity, but find no substantial impact of influenza A virus.

## 1 Introduction

Human rhinoviruses (RVs) are one of the most prevalent pathogens responsible for the common cold, accounting for more than 50% of upper respiratory tract infections, especially among young children [1, 2, 3, 4]. They are are non-enveloped, positive-sense, single-stranded RNA viruses belonging to the family *Picornaviridae* and genus *Enterovirus* (EV) [1, 3]. RVs are ubiquitous, circulating globally and year-round [5, 1, 3, 6], usually with seasonal peaks in spring and fall. Transmission occurs via direct person-to-person contacts, aerosols, and fomites [7, 8, 9, 10]. RVs are currently classified into three species (RV-A, B, and C), encompassing over 170 subtypes [11, 3, 1]. Importantly, cross-reactivity between subtypes appears to be weak (i.e., offering little protection against heterotypic infection), such that this wide antigenic diversity has hampered the development of antiviral treatment or vaccines [3, 1, 4, 12]. While infections are typically mild, they may occasionally lead to more severe diseases like asthma exacerbation and chronic obstructive pulmonary disease [1, 4]. Additionally, the associated global health and economic burden is substantial (albeit difficult to assess precisely [11]), costing billions of US dollars annually due to medical visits and work absenteeism [1, 3].

One mystery about RV remains its potential interaction with influenza virus (IV), particularly influenza A virus (IAV). We refer here to negative heterologous virus-virus interactions (or viral interference), whereby one respiratory virus suppresses another, for example by triggering an antiviral defense or through the competition for susceptible cells [13, 14, 15, 16, 17, 18, 19]. A range of observational and experimental studies supports the existence of viral interference between RV and I(A)V at different scales, although with some conflicting findings:

*•* **Uncertain directionality or bidirectional interaction**: At the population level, RV and IAV dynamics are typically asynchronous [17, 18, 15, 20, 21] – though this could also reflect other factors such as differences in seasonality [18] –, and can also exhibit divergent spatial distribution patterns [22]. Statistical analyses in [17] found a strong negative correlation between RV and IAV monthly prevalence, even after adjusting for seasonality. Reduced likelihood of co-detection have also often been reported [23, 17, 18, 15, 20, 24], but co-detection prevalence studies may be unreliable for the inference of interaction [25, 26]. In [27], the authors found a mutual reduction in infection risk.
*•* **RV affecting IAV**: Epidemiological data suggested that a major RV outbreak may have delayed the 2009 IAV (H1N1pdm09) pandemic in Europe [28, 29, 30]. Similarly, [22] found a suppressing effect of RV on influenza outbreaks. At the within-host level, experimental studies showed that prior RV infection can inhibit infection with IAV [18], reduce disease mortality and enhance IAV clearance [31], or inhibit IAV replication [20].
*•* **IAV affecting RV**: Conversely, analyses of virus detection frequency in [32] suggested interference of RV infection by IV. Experimental evidence found that IAV H1N1 can inhibit RV replication, whereas RV does not inhibit subsequent IAV H1N1 infection [33]. Besides, mathematical simulations supported the idea that a transient immune-mediated refractory period induced by IAV could account for the observed decline in RV infections during peak IAV activity [17]. This asymmetric interaction is further consistent with longitudinal (individual-level) data from 2009 showing that IAV H1N1 infection reduced the subsequent risk of RV infection the following week, while RV infection did not confer a protective effect against IAV H1N1 [34, 35, 36].

In summary, although various studies support its existence, RV-IAV interaction still largely remains unclear, in particular whether it might be mutual or more asymmetric. This viral interference has mostly been experimentally associated to interferons (IFNs) [31, 18, 33]; although IFN signaling is part of the host innate (non-specific) immunity, asymmetric interactions may still arise due to differences in response timing and/or magnitude, as well as in the sensitivity of each virus. Besides, ecological (as opposed to immunological) mechanisms [37], such as changes in contact rates after a primary infection, may also play a role. In this work, we adopt a phenomenological approach to examine the potential influence of IAV on RV population dynamics.

The recent implementation of non-pharmaceutical interventions (NPIs) during the coronavirus disease 2019 (COVID-19) pandemic significantly disrupted the dynamics of respiratory pathogens [38, 39, 40, 41, 42]. The COVID-19 pandemic is particularly relevant to our analysis for two reasons. First, in most locations, RV continued to circulate at appreciable levels. At the onset of the pandemic, RV infections declined in many countries across the globe during periods of strict NPIs, though to a lesser extent than other respiratory pathogens [43, 39, 44, 45, 46, 47, 48, 40, 49, 41, 50, 51], and rebounded sharply following NPI relaxation [43, 39, 46, 47, 50, 52]. IAV, on the other hand, remained largely undetected during periods of strict NPIs as well as periods of gradual NPI relaxation, finally rebounding in 2022 or later. Interactions between respiratory pathogens such as RV and IAV were often cited to have potentially played a role during the COVID-19 pandemic, e.g., [39, 32, 50]. One question would thus be: is the persistence of RV during this period enhanced by the absence of IAV? Second, endemic pathogens typically exhibit stable endemic cycles, complicating the possibility of teasing apart the contributions of different transmission drivers. Extrinsic shocks – such as sudden changes in the recruitment rate of susceptibles (e.g., baby booms) or vaccination campaigns – have previously been used to better understand pathogen dynamics [53, 54], and NPIs represent another form of such shocks that could be leveraged opportunistically [55].

Here, we focus on the population dynamics of RV and on the potential influence of IAV. In contrast to other respiratory pathogens, population-level model fitting remains rare for RV. Using a Bayesian inferential framework, we develop a simple mathematical model to capture RV incidence dynamics, including IAV detection as an external input to test the hypothesis of IAV interference on RV dynamics. We first conduct a simulation study to validate our approach, and in particular to assess how exogenous perturbations can help to quantify the strength of interaction between endemic pathogens. We then apply our framework to historical national and regional time series data in the US and Canada. Using COVID-19 pandemic NPI perturbations, we estimate key parameters of RV and evaluate the strength of the effect of IAV.

## 2 Results

### 2.1 RV and IAV population dynamics across the US and Canada

We collected time series of weekly tests and detections for RV/EV and IAV from longstanding respiratory virus surveillance systems in the US and Canada at both the national and regional level (a map is shown in **Fig. S1**). Rapid laboratory diagnostic testing does not differentiate RVs from other EVs, though RVs are expected to account for most detections [56, 57]. For brevity, we thus refer to RV/EV simply as RV. We plot longitudinal data at the national level in **Fig. 1** (see **Fig. S2** and **S3** for time series at the regional level).

**Figure 1:**
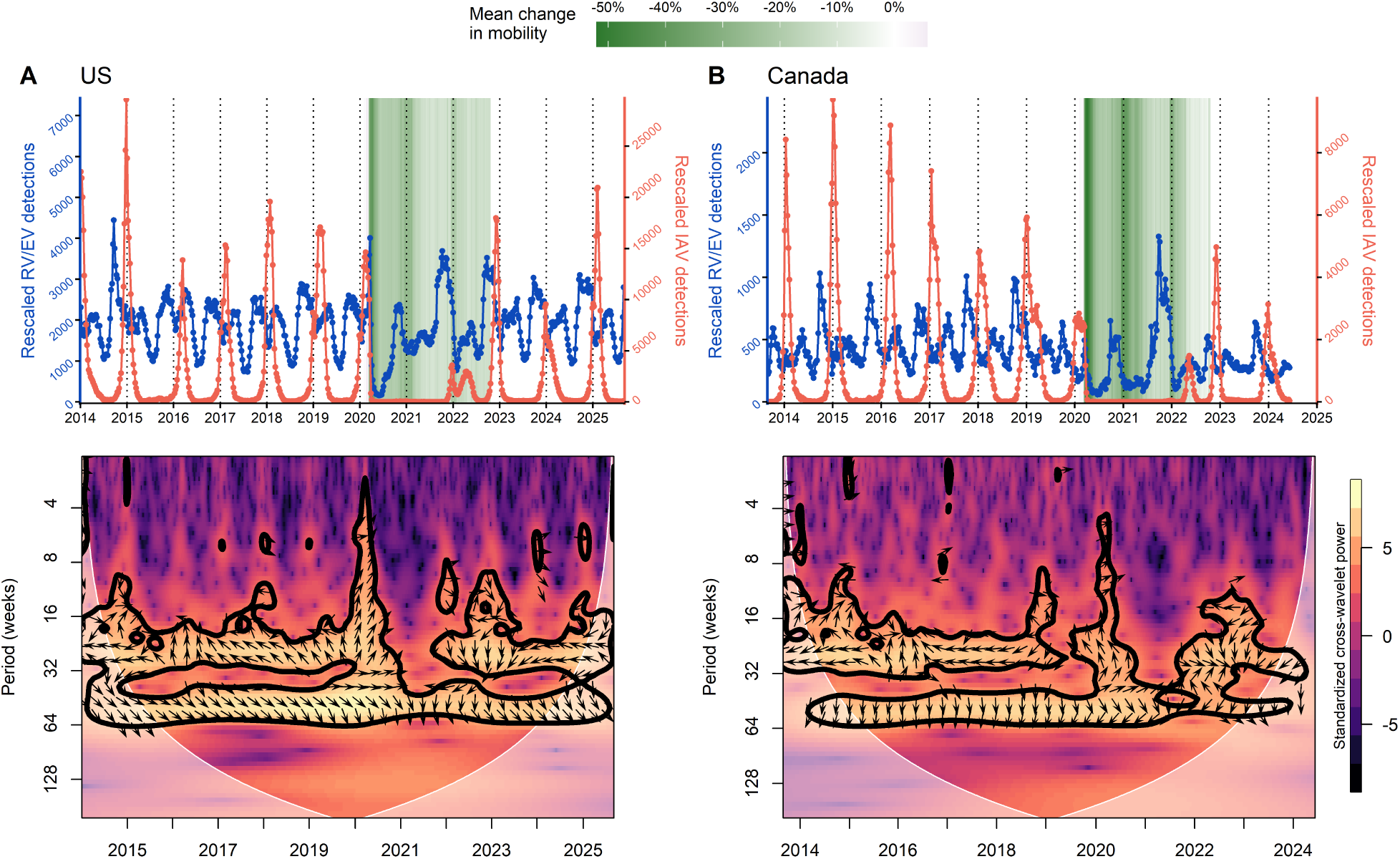
RV and IAV circulation at the national level (US and Canada). Top: RV (blue) and IAV (red) surveillance detections in (A) the US and (B) Canada; data have been rescaled to mitigate biases due to circulation of other respiratory pathogens and changes in testing effort (see **Materials and Methods §4.1**). Mean change in mobility (colored background) during the COVID-19 pandemic were computed from Google COVID-19 Community Mobility Reports [83]. See **Fig. S2** and **S3** for time series at the regional and provincial level. Bottom: Cross-wavelet transform of IAV and RV detections in (A) the US and (B) Canada. We used the function xwt from the R package biwavelet [93] (see details in **supporting information §S1**). Colors indicate standardized cross-wavelet power, with 95% significance region outlined by a contour. Arrows pointing left (resp. right) suggest IAV and RV detections are out-of-phase (resp. in phase) and arrows pointing down (resp. up) suggest IAV detections lead (resp. lag) RV detections (with vertical arrows indicating a phase difference of *π/*2 (quarter cycle)). Here, results confirm the asynchronous circulation of IAV and RV at the national scale: we find that detections are predominantly out-of-phase at subannual periods (≈ 16 − 32 weeks), as well as an anti-phase behavior with RV lagging at more annual periods (≈ 40 − 64 weeks). This pattern is transiently disrupted during the COVID-19 period (2021-2022), but is restored in the subsequent years. See **Fig. S6** and **S7** for plots at the regional and provincial level.

Across these locations, RV exhibit biannual outbreaks before the pandemic, with a peak in fall and a (usually smaller) peak in spring. Pre-pandemic IAV dynamics are annual with a peak (of varying magnitude) in winter. We used cross-wavelet transform analyses to confirm and complement the initial visualization and description of the time series. Results in **Fig. 1** confirm that national IAV and RV detections are predominantly out-of-phase at subannual periods (≈ 16 − 32 weeks), except in 2016 when the IAV outbreak seems delayed; and indicate an anti-phase behavior with RV lagging at more annual periods (≈ 40 − 64 weeks). We recover similar patterns at the regional level, though with some variability across locations (**Fig. S6** and **S7**).

Importantly, IAV effectively disappeared following NPI implementation at the onset of the COVID-19 pandemic. In the US, this period was followed by a small rebound late 2021/early 2022 and a bigger rebound in the following season, similar to pre-pandemic levels) (**Fig. 1-A** and **S2**). IAV rebounds occurred later in Canada, between mid-2022 and early 2024 across regions of Canada (**Fig. 1-B** and **S3**). Conversely, while RV detections dropped at the very beginning of the COVID-19 pandemic, they continued to circulate during the pandemic at approximately similar levels (**Fig. 1, S2** and **S3**).

In the following, we use a seasonally-forced Susceptible-Infectious-Recovered-Susceptible (SIRS) model to fit RV incidence time series (see **Materials and Methods §4.2** and **Table 1**). Briefly, the transmission rate is decomposed into three terms: (i) a periodic term *β* consisting of weekly transmission rates that can handle any seasonal pattern within a year, similar to [58], (ii) a timevarying term that captures contact changes due to COVID-19 pandemic NPIs and (iii) a timevarying term that phenomenologically captures potential effect of viral interaction due to IAV. We use IAV incidence data as an external input (or covariate, as in [59]), which is assumed to force RV transmission by some factor *ϕ*. Our aim is to detect whether signatures of an interaction of IAV on RV dynamics exist at the population level. These effects could be derived from within-host viral interference mechanisms (e.g., IFNs in the context of innate immunity) or ecological interaction mechanisms (e.g., changes in contact rates after a primary infection) [37, 55]. While viral interaction effects can be characterized by their direction, intensity and duration, however our approach cannot easily estimate the duration of these effects. We start by using the concurrent weekly incidence of IAV, assuming the duration of interaction is short-term (on the order of days to one week consistent with e.g., interactions mediated primarily by IFNs).

**Table 1:**
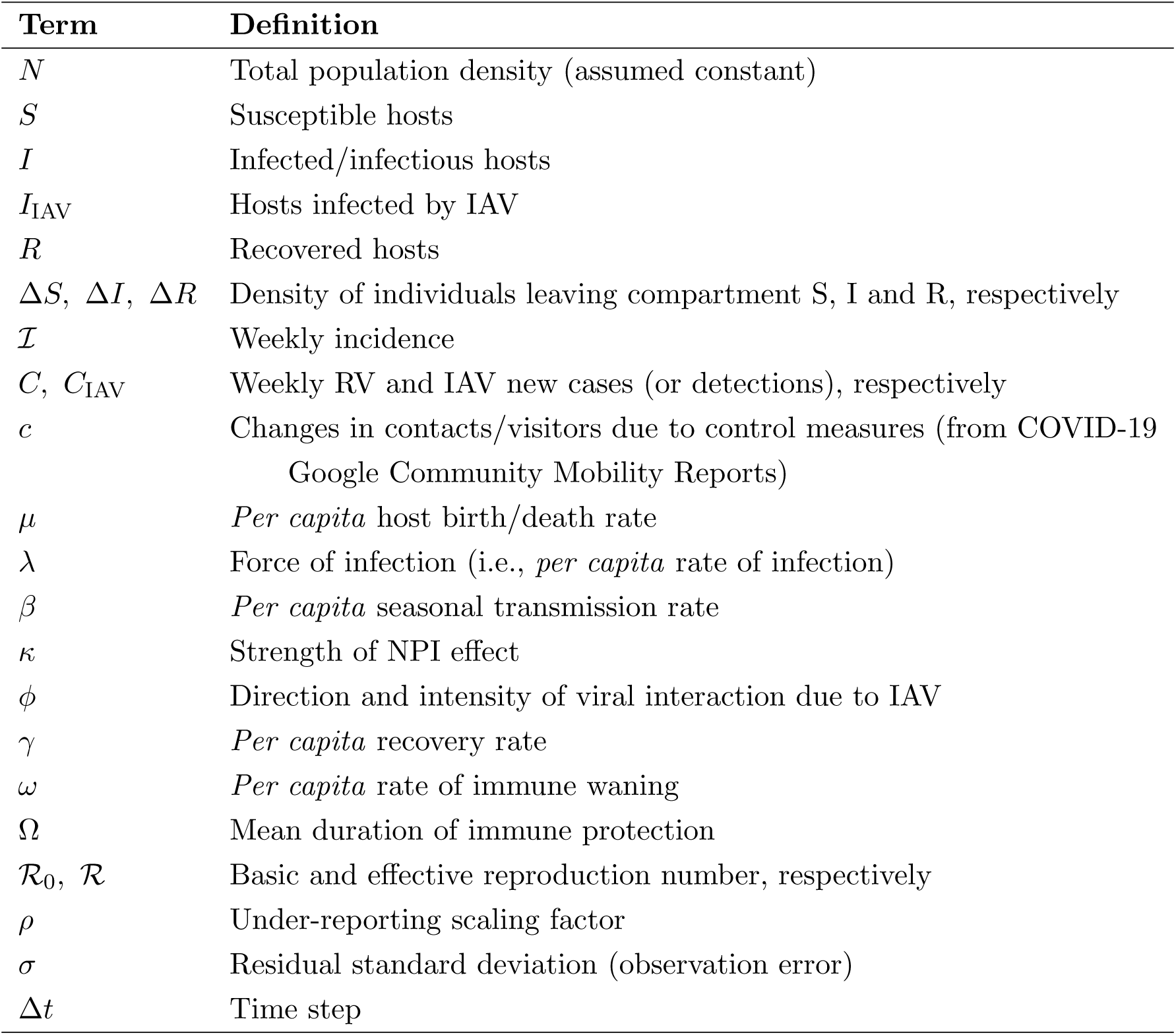
Main notations. For the sake of simplicity, pathogen-associated state variables and parameters correspond to RV, unless otherwise specified with the subscript IAV.

| Term | Definition |
| --- | --- |
| $N$ | Total population density (assumed constant) |
| $S$ | Susceptible hosts |
| $I$ | Infected/infectious hosts |
| $I_{\text{IAV}}$ | Hosts infected by IAV |
| $R$ | Recovered hosts |
| $\Delta S, \Delta I, \Delta R$ | Density of individuals leaving compartment S, I and R, respectively |
| $\mathcal{I}$ | Weekly incidence |
| $C, C_{\text{IAV}}$ | Weekly RV and IAV new cases (or detections), respectively |
| $c$ | Changes in contacts/visitors due to control measures (from COVID-19 Google Community Mobility Reports) |
| $\mu$ | *Per capita* host birth/death rate |
| $\lambda$ | Force of infection (i.e., *per capita* rate of infection) |
| $\beta$ | *Per capita* seasonal transmission rate |
| $\kappa$ | Strength of NPI effect |
| $\phi$ | Direction and intensity of viral interaction due to IAV |
| $\gamma$ | *Per capita* recovery rate |
| $\omega$ | *Per capita* rate of immune waning |
| $\Omega$ | Mean duration of immune protection |
| $\mathcal{R}_0, \mathcal{R}$ | Basic and effective reproduction number, respectively |
| $\rho$ | Under-reporting scaling factor |
| $\sigma$ | Residual standard deviation (observation error) |
| $\Delta t$ | Time step |

### 2.2 Exogenous perturbations as a powerful tool to infer pathogen-pathogen interaction

We first conducted a simulation study to assess our ability to recover model parameter values, and in particular the direction and intensity of the viral interaction of IAV on RV, *ϕ*. Using a two-pathogen transmission model, we first simulated epidemiological trajectories for RV and IAV (**Fig. S8**), from which we generated synthetic time series; we then fitted RV simulated data while IAV simulated data was only used as an external input. The primary confounder of the population-level effects of viral interactions is seasonality in transmission, so here we generated simulated data under three main scenarios, each consistent with the sub-annual patterns of RV epidemics. In scenario 1, RV transmission corresponds to a bimodal seasonality with no interaction (*ϕ* = 0); in scenario 2, IAV negatively impacts RV (*ϕ <* 0) but we adjust the seasonal forcing so that the overall transmission rate matches scenario 1 (“masked interaction”); in scenario 3, RV seasonality corresponds to a classic unimodal, sinusoidal forcing, and the two annual RV outbreaks arise from the interference by IAV. We plot the different seasonal transmission profiles in **Fig. 2-AB**. Simulated data include a pre-pandemic period of 6 years and a (post-)pandemic of 4 years, aligned with our empirical surveillance data. See more details on model structure and fitting in **Materials and Methods §4.2-4.3** and simulation experiment in **Materials and Methods §4.4**.

**Figure 2:**
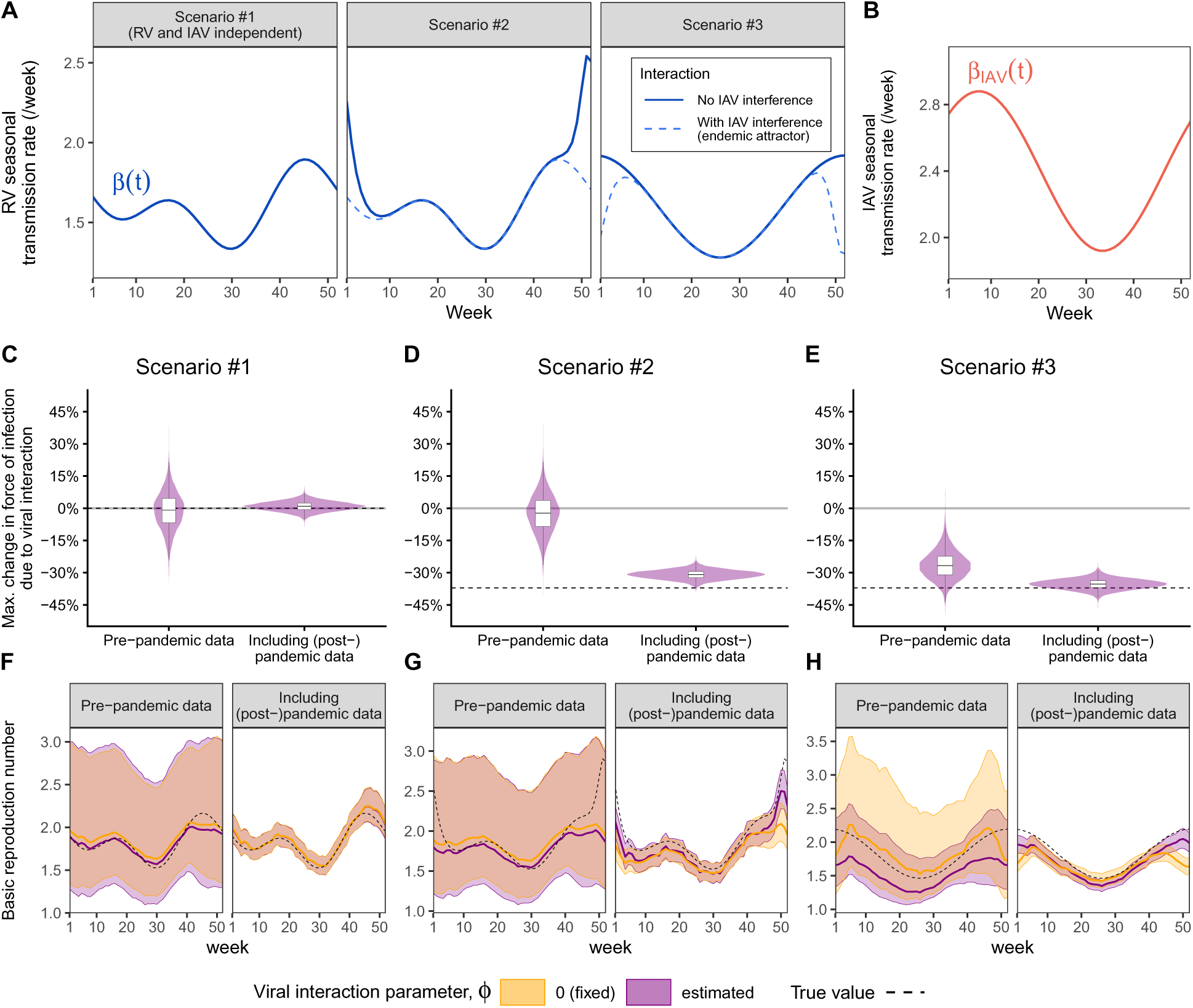
Perturbations can be a powerful tool for estimating pathogen-pathogen interactions. We conducted a simulation study to validate our ability to estimate model parameters. For simplicity, we only display here the results obtained with the main simulations of each scenario. (A) RV seasonal transmission (solid blue lines) and effect of the interference due to IAV when the latter is at its endemic attractor (i.e., no perturbation, dashed blue lines). Importantly, RV and IAV are independent in scenario 1 (no interaction) and this seasonal forcing is mimicked in scenario 2 in the presence of IAV. (B) IAV seasonal transmission (same across scenarios). (C-E) Posterior distributions of the estimated viral interaction term. (F-H) Estimated profiles of RV basic reproduction number (median values (lines) and 95% CrIs (shaded envelopes)), reflecting seasonal forcing terms. Black dashed lines represent true values used in the simulations. See **Table S1** for parameter values and see **Fig. S9**, **S10** and **S11** for the full results.

Simulation study results showed that including exogenous perturbations can enable us to infer pathogen-pathogen interaction. In scenario 1 (no interaction), the model consistently inferred that RV dynamics is independent of IAV (**Fig. 2-C** and **S9-A**). In scenario 2 (“masked interaction”), the model failed to detect the presence of viral interaction when using the pre-pandemic data alone, but including pandemic perturbations substantially improved the inference, enabling both the viral interaction and the seasonal forcing terms to be estimated correctly (**Fig. 2-DG** and **S10-A**). In scenario 3 (simple seasonality with interaction), model fit using pre-pandemic data correctly detected a negative interaction but did not accurately estimate its magnitude, which was underestimated with a wide CrI (**Fig. 2-EH** and **S11-A**). Here, viral interaction was only partially captured by other parameters (in particular the seasonal forcing), likely because the reduction in the force of infection coincident with the increase in IAV was more abrupt than in scenario 2. However, fitting the model with and without interaction (i.e., estimating *ϕ* or fixing *ϕ* = 0, respectively) yielded similar fits to the pre-pandemic data (**Fig. S11-A**). Including the (post-)pandemic period resulted in much better estimates of both the viral interaction effect and seasonal forcing. Across scenarios, restricting the inference to the pre-pandemic period resulted in wider CrIs for all model parameters, with the exception of initial conditions (**Fig. 2**, **S9-A**, **S10-A** and **S11-A**); in particular, the mean duration of immune protection, Ω, and the under-reporting factor *ρ*, were more difficult to estimate precisely and were usually positively correlated. Our results were robust to observation noise (**Fig. S12**). As a sensitivity analysis, we also varied the chosen values of some key parameters of the viruses, showing that most estimates remained robust across the explored range of values (**Fig. S13**).

For each scenario, we also examined two extensions (we refer to the previous cases as the main simulations). First, we considered a six-month shift in the timing of NPIs, such that interventions implemented in winter (typically stricter) occurred in the preceding summer, leading to earlier postpandemic rebounds of IAV. Overall, varying the timing of NPIs resulted in similar results (**Fig. S9-B**, **S10-B** and **S11-B**). Second, we introduced a one-off exogenous perturbation in the dynamics of IAV in the pre-pandemic period (a “kick”, transferring 35% of the susceptibles to IAV to the recovered compartment, e.g., due to a vaccination campaign). Here, in scenarios with viral interference (2 and 3), fitting the model to the pre-pandemic period alone yielded more accurate parameter estimates than for the main simulations, which only featured regular cycles (**Fig. S9-C**, **S10-C** and **S11-C**). Overall, in the absence of viral interference, the model performed consistently across all cases. In the presence of viral interference, including any form of perturbations enabled accurate estimation of the viral interaction and seasonal forcing terms. Estimation of seasonal forcing was improved in particular for weeks of high IAV circulation, which peaks around week 52, while errors remained consistently smaller during periods of low IAV circulation (trough centered around week 30) (**Fig. S14**).

In this section, we used simulations to illustrate how perturbations can be a powerful tool for estimating pathogen-pathogen interactions. This analysis indicates that COVID-19 pandemic NPIs can be used as a large-scale natural experiment for examining such interactions, which would not be not possible when available data only include endemic limit cycles. In contrast to perturbations solely affecting IAV, NPIs represent a distinct class of perturbations that, in addition to potential indirect effects mediated through IAV dynamics, also directly affects RV transmission. One major limitation of NPIs may be the unknown *shape* of the perturbation through time. Additional simulations showed that, when the model also had to infer this shape without constraints, it tended here to underestimate the magnitude of the interaction effect (though the estimated direction remained correct here) while overestimating seasonal transmission rates (**Fig. S15**). Constraining the shape of these perturbation seems thus essential for accurately estimating the different drivers of the force of infection. In practice, the true shape is unknown, and we used mobility data as a proxy.

### 2.3 Modeling of empirical surveillance data does not support an impact of IAV on RV dynamics at the population level

We then fitted our model to RV detection time series from the US and Canada, including both preand (post-)pandemic period. The model converged for all locations except the US HHS region 1 and the Canadian Atlantic province; examples of MCMC chains and posterior distributions are provided in **Fig. S16**. Our model captures most of the RV outbreaks (**Fig. 3**, **S17** and **S18**) – though it underestimates the 2021 fall outbreaks in several locations. Using the current incidence of IAV, the posterior distributions of *ϕ* – and thus of the change in transmission due to viral interaction, with negative values suggesting that IAV epidemics suppress RV activity – are either not significantly different from 0 or relatively weak in magnitude across locations (**Fig. 4**). For example, at the national level, we obtained a median *ϕ̂* = −0.024 (95% CrI: [−0.051, +0.003]) for the US and *ϕ̂* = +0.035 (95% CrI: [−0.002, +0.071]) for Canada. A sensitivity analysis varying the prior constraint on seasonal transmission variability yielded similar results (**Fig. S19**). Model comparisons of expected log-predictive density against a null model with no viral interaction (fixing *ϕ* = 0) did not support an effect of IAV on RV dynamics in any location (**Fig. S20**). We also explored the possibility that interaction might be better captured by considering longer-lasting effects (i.e., if interaction effects would last more than 1 week), so that prior IAV incidence could also affect RV dynamics. Refitting the model using a lagged sum of IAV incidence, rather than current incidence, with a lag of 1, 3, or 5 weeks (see **Materials and Methods §4.3**) led to similar results (**Fig. 4** and **S20**). Surprisingly, viral interaction estimates *ϕ̂* tend to be more positive for Canadian locations and more negative for American locations. Whether these small differences are meaningful is unclear and otherwise difficult to explain. The different surveillance systems and the usually earlier timing of IAV rebounds in US locations seem to be the two main differences between the two countries. These results were obtained when fitting the model independently to each location. We explored how fitting the model simultaneously to all regions/provinces impacted the results, constraining *ϕ* to be shared across locations within a given country, and the rate of immune waning *ω* across all locations (see **supporting information §S3**). In this case, estimates of viral interaction *ϕ̂* remained centered around 0 for both US and Canada, with no difference in sign, but the Canadian estimate was highly uncertain (**Fig. S21**). We also considered different extensions of the main analysis, showing that our results remained robust to: (i) post-pandemic behavioral changes (**Fig. S22**), (ii) the use of total IV incidence, i.e., including influenza B virus (IBV) detections, (**Fig. S23**) and (iii) the addition of an exposed RV stage, i.e., infected but not yet infectious, (**Fig. S24**).

**Figure 3:**
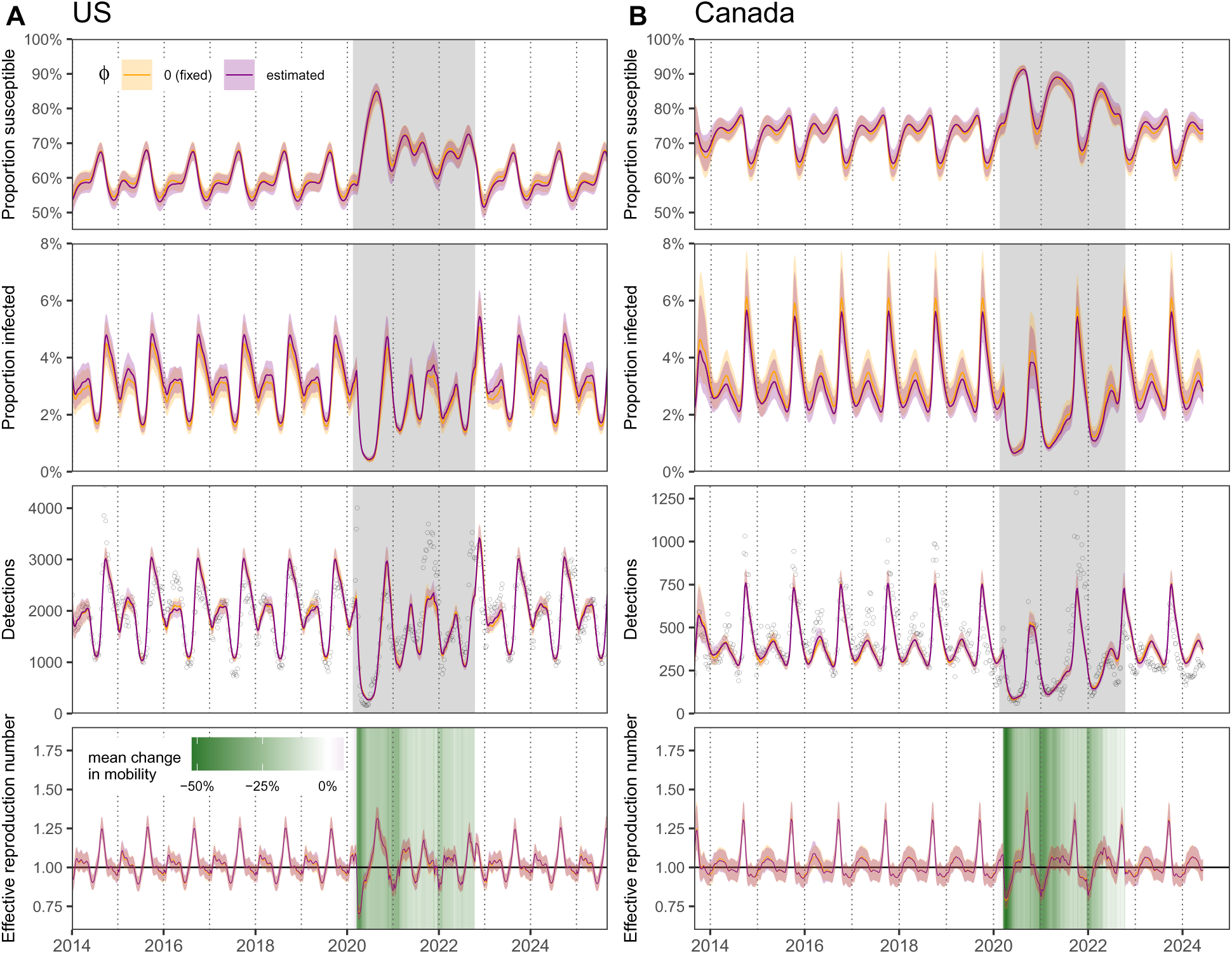
Fitted model results in the US and Canada. We independently fitted model (3)-(4) with seasonal transmission (8) (see **Materials and Methods §4.2-4.3**) to rescaled RV detections (black points) in (A) the US and (B) Canada. Detections were rescaled to account for changes in testing patterns. We compare here results of the model where IAV can affect the transmission of RV by estimating *ϕ* (yellow) or not by fixing *ϕ* to 0 (purple). We plot posterior median values (lines) and 95% CrIs (shaded envelopes) for: from top to bottom, (i) the proportion susceptible *S/N*, (ii) the proportion infected *I/N*, (iii) the number of detections and (iv) the effective reproduction number R (the epidemic grows as soon as R *>* 1 (black horizontal line)). Colored backgrounds indicate the pandemic period (mean change in mobility during the COVID-19 pandemic were computed from Google COVID-19 Community Mobility Reports [83]). See **Fig. S17** and **S18** for fitted model results at the regional/provincial level.

**Figure 4:**
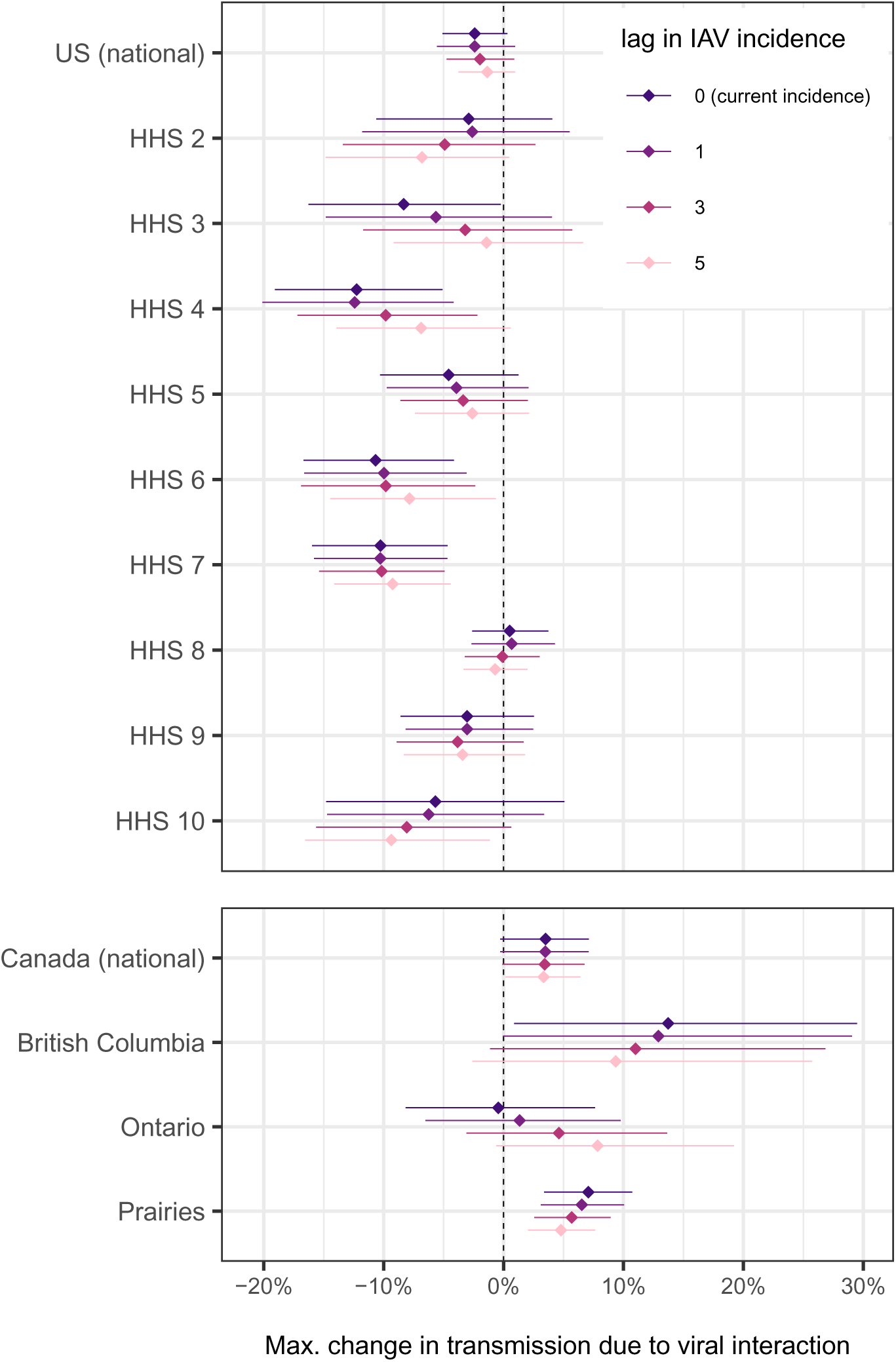
Viral interaction estimates for the US and Canada. Posterior median values (points) and 95% CrIs (segments) of the maximum change in RV transmission due to IAV interaction. We recall that, for the purpose of model fitting, the effect of viral interaction is modeled as *ϕ C*_IAV_[*t*]*/* max *C*_IAV_[*t*], where *C*_IAV_[*t*] denotes the observed incidence of IAV, so that *ϕ* represents the maximum change in the force of infection of RV due to viral interaction (i.e., during periods of highest IAV circulation). If *ϕ* is positive, it means IAV facilitates RV transmission, if negative, it suppresses RV transmission, and if zero, there is no interaction. We also considered lagged sums of IAV incidence, substituting *C*_IAV_[*t*] by 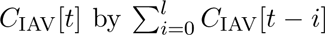 with a maximum lag *l* of 1, 3 or 5 weeks (*l* = 0 refers to current incidence only).

We plot in **Fig. 5-A** and **C** the estimated weekly transmission rates of the more parsimonious model (i.e., without viral interaction). Both the magnitude and seasonal profile vary across locations (in particular in the US) but, as expected, a common seasonal pattern includes a drop in transmissibility around the summer (typically between week ≈ 20 and 35), followed by an increase in fall. We also plot in **Fig. 5-B** and **D** the posterior distributions of the mean duration of immune protection to RV reinfection, Ω. Immune protection was estimated to be quite short (typically around 1-3 months), despite some variability across locations (e.g., a median of 20 weeks (95% CrI: [16.41, 24.43]) in the HHS region 9). Posteriors of the remaining parameters are displayed in **Fig. S18** and summarized in **Table S3**. Note that estimates of the under-reporting factor *ρ* cannot be interpreted quantitatively as *ρ̂* can also compensate for the arbitrary rescaling of detections. Using our estimates, the median of the basic reproduction number R_0_ was estimated between ≈ 1.15−3.35 for the US (**Fig. 5-A**) and ≈ 1.21 − 1.97 for Canada (**Fig. 5-C**), which is consistent with existing literature suggesting a range between 1.2 and 2.7 [60, 61, 62, 63]. Averaged over the entire seasonal profile, mean values range from 1.16 to 2.04 for the United States and from 1.24 to 1.42 for Canada. The effective reproduction number R ranges between approximately 0.8 and 1.5 in classic seasons, following the biannual pattern of RV (primary peak in fall and a smaller secondary peak in spring). During the first implementation of NPIs at the onset of the COVID-19 pandemic (March-May 2020), R shows its biggest decline, but followed by a rapid return to historical levels (**Fig. 3**, **S26** and **S27**).

**Figure 5:**
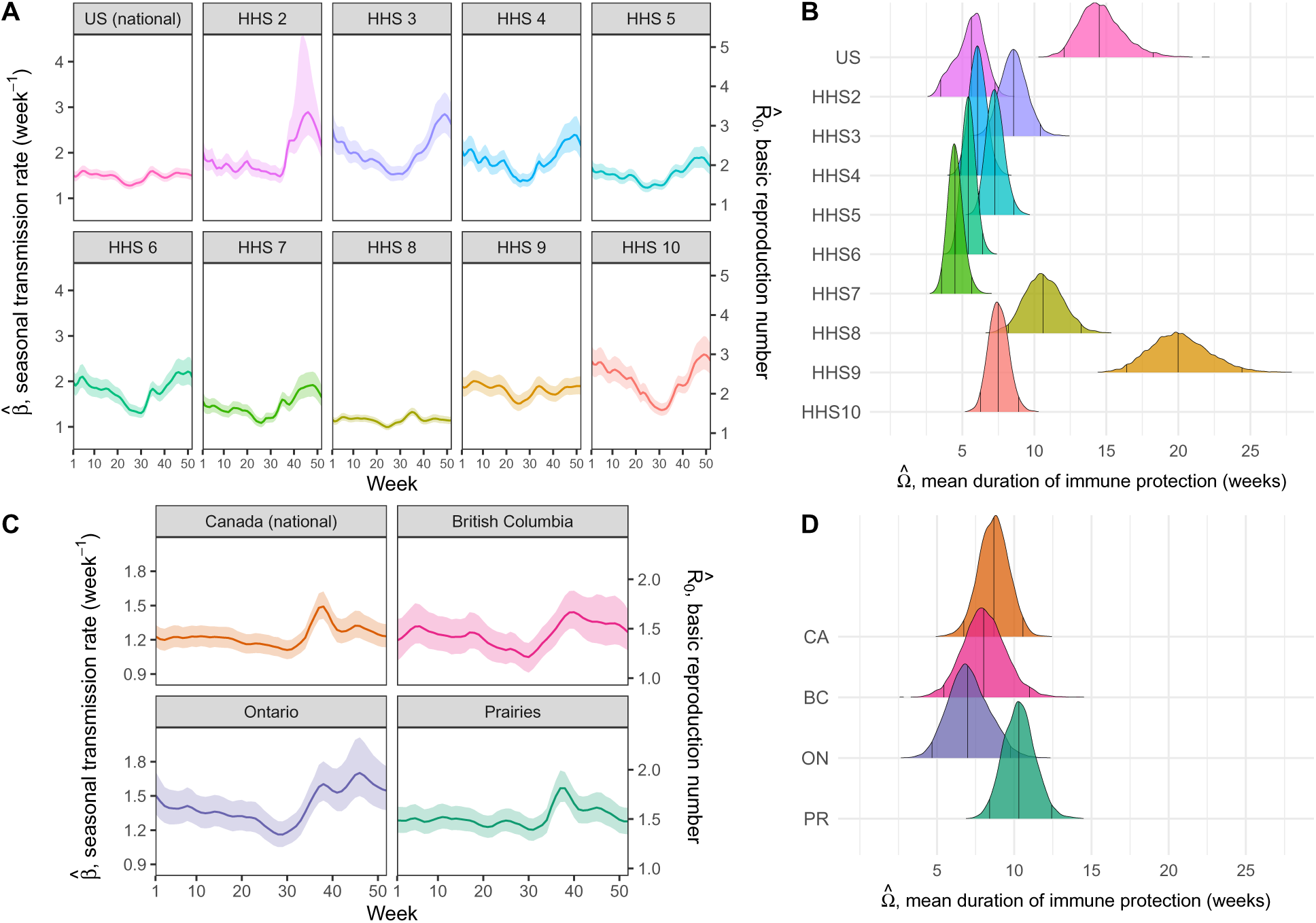
Estimated transmissibility and duration of immune protection of RV. We plot results obtained with the model with no viral interaction (*ϕ* = 0). First column: estimated profiles of RV weekly transmission rate *β* (left y axis) and basic reproduction number *R*_0_ (right y axis) in (A) the US and (C) Canada; the lines indicate the median values and the envelopes represent and 95% CrIs of the inferred posterior distributions. Second column: posterior distributions of the mean duration of immune protection Ω in (B) the US and (D) Canada; vertical black lines represent 2.5%, 50% (median) and 97.5% quantiles, respectively. Hat symbols denote estimated values. CA=Canada (national), BC=British Columbia, ON=Ontario, PR=Prairies.

## 3 Discussion

Here, we investigated the population dynamics of human RVs and aimed to characterize the presence of potential viral interaction due to IAV. Historical time series from the US and Canada (at both the national and regional level) confirmed the asynchronous dynamics of RV and IAV, as well as their contrasting responses to COVID-19 pandemic perturbation, during which IAV disappeared while RV continued to circulate. In line with other studies, we hypothesized that IAV-RV viral interference may (partly) explain these epidemiological patterns; and we leveraged pandemic perturbations as a large-scale natural experiment to put this hypothesis to the test.

We modeled RV population dynamics using a seasonally-forced SIRS model, using IAV incidence as an external input. Note that this does not imply that RV cannot also influence IAV dynamic. We first conducted a simulation study to evaluate our ability to accurately estimate model parameters in various settings, and especially viral interaction due to IAV. Simulation results demonstrated that including exogenous perturbations such as COVID-19 pandemic NPIs can be key to accurately estimate endemic pathogen interaction. Using only stationary oscillatory dynamics hinders the identifiability of different epidemiological factors on the force of infection, such as seasonality and virus-virus interaction, but perturbations can provide additional information to disentangle these different drivers. The utility of such perturbations may depend on characteristics such as their intensity, duration, and timing. In contrast to perturbations that only affect the dynamics of one infectious disease (e.g., vaccination, antigenic shift), NPIs affect the transmission of both diseases. The shape of these perturbations is usually unknown, which can hamper the estimation process in the absence of additional constraints. Here, we relied on Google mobility data, although this approach still remains imperfect. We then fitted our phenomenological model to historical RV time series, including both preand (post-)pandemic periods. Results did not support an effect of IAV on RV dynamics at the population level in any location, although this does not completely rule out the possibility of an interaction. For example, available data may lack sufficient statistical power to detect more subtle effects – in particular, RV dynamics are extremely fast due to rapid loss of immunity, IAV is likely to have (if any) only a short-lived interaction, and IAV circulation is low most of the year. Even strong individual-level effects can be masked at the population level [59]. Viral interactions may also affect disease severity without altering susceptibility or transmission. For example, IAV infection could reduce the probability of hospitalization or severe disease following RV infection, an effect that we would not be able to see in our surveillance data which include many mild clinical infections. Moreover, RV seasonal forcing may be driven by other factors for which we have little data, such as a combination of climatic effects and human behavior (which is expected to vary geographically). Additionally, waning immunity can also drives sub-annual epidemics [64, 65]. More broadly, many “unknowns” may be contributing to RV dynamics, but resolving one or a few of them could substantially improve inference of interaction effects.

The mechanisms behind RV resilience to pandemic perturbations still remain unclear, and our results did not support the hypothesis that RV benefited from the absence of IAV during this period. Nevertheless, other hypotheses remain. For example, the efficacy of NPIs against RV transmission may be limited. For instance, face masks seem less effective in blocking RV particles (≈ 30 nm), smaller than IV or SARS-CoV-2 particles, [66] – although they are unlikely to be entirely ineffective, since RV circulation sometimes increased once mask mandates had been lifted [46]. As a non-enveloped virus, RV may also exhibit greater environmental stability and be less affected by hand washing and surface disinfection, thereby sustaining different transmission routes such as via fomites [50, 40, 32, 47, 44]. Yet, the concomitant decrease in the circulation of other non-enveloped viruses may challenge this hypothesis [47]. Besides, numerous studies underscore that children are a key driver of the spread of RVs [67, 68, 49, 51], especially in schools and daycare centers with subsequent transmissions to adults within households. Since children are less likely to adhere to hygiene measures, their role in maintaining RV circulation during NPIs is a plausible explanation [50]. This is supported by the increase in RV frequency among young children reported in Japan during the pandemic [32], and also consistent with the sharp increase in RV infections after schools reopened [43, 39]. RV resurgence was first observed among children in Germany [47], but RV cases began rising even before daycare centers and schools reopened in Finland [41]. Last, our estimates of the duration of immune protection support that infection only confers short-lived protection against reinfection, likely reflecting the co-circulation of many serotypes with limited cross-immunity. Specific neutralizing antibodies typically have little cross-reactivity, recognizing only one serotype [69, 4, 70]. While T-cell immunogenicity exists, with some degree of subtype cross-reactivity [69, 12], cellular immunity responses also remain poorly understood. Crucially, shorter immune protection implies a faster replenishment of the susceptible population, consistent with the better resilience of RV to pandemic perturbations [42].

Our work is closely related to [55], which estimated viral interference between two closely related pneumoviruses, respiratory syncytial virus (RSV) and human metapneumovirus (hMPV), by comparing two hypotheses: (i) the two pathogens are independent with different seasonal forcing or (ii) same seasonal forcing but RSV has a negative effect on hMPV. In this work, a two-pathogen model was first fitted to the pre-pandemic period, while post-pandemic rebounds were then used as an out-of-sample test. We could not make the assumption here that RV and IAV are subject to the same seasonal forcing, and had to use a flexible parameterization for RV seasonal transmission, which may make the other components of the force of infection more difficult to estimate. Besides, while using IAV data as an external input also directly introduces observation noise in RV transmission compared to fitting a two-pathogen model – where IAV prevalence could be reconstructed from latent variables –, surveillance time series may contain richer variability and signal capturing the complex dynamics of IAV (including antigenic variation, subtype interference, external seeding, pandemic perturbation) that could be lost if not adequately captured by the model. Our framework preserves this empirical variability, which is noisier (though we smoothed IAV data) but may also be more informative for inferring the potential effect of IAV on RV dynamics. Although our approach is statistically less powerful, it is also more general, and can be more easily extendable to other pathogens. More broadly, viral interference is fundamentally a within-host interaction mechanism that can subsequently shape population-level pathogen transmission dynamics. While a detailed multi-pathogen model (as in, e.g., [17]), may provide a more satisfying mechanistic description of immunity and co-circulation patterns, it would be difficult to fit to incidence data alone without additional information and assumptions. In contrast, our approach is intentionally phenomenological. Our model is intentionally simple and does not aim to explicitly resolve within-host interactions; instead, it provides an effective description of observed incidence dynamics and allows us to probe for epidemiological signatures consistent with altered transmission patterns that integrate both withinhost viral interference (immunological) mechanisms and ecological mechanisms of interactions [37].

There are several limitations to our work. First, we assumed a homogeneously mixing population. However, children are likely to be a natural reservoir for RV infections, and a key driver of transmission to adults [67, 68, 49] – infections are usually symptomatic among children but asymptomatic among adults [68]. Heterogeneity in mixing patterns may have profound consequences on pathogen transmission, but accounting for it would also require age-stratified data, alongside contact matrices. Age was not available for the surveillance data used in this study. It may however contribute to residual variation that is not explicitly captured by the model, and the use of aggregated data (despite the advantage of larger sample sizes) may obscure some important age-dependent patterns. Addressing how age-specific differences affect our results would require an extension of the current modeling framework and would therefore constitute a natural future direction to evaluate how this could improve model fit and parameter estimation. It would also be interesting to investigate if RV dynamical patterns could be better captured by school and daycare center openings and closings, which were also disrupted during the pandemic period. Reductions in RV infections in the summer and in the winter may reflect changes in children mixing due to school holidays.

Second, we modeled RVs as a single-strain pathogen, neglecting the different RV species and their wide antigenic diversity. This is important because our estimates therefore represent some averages across multiple strains – whereas quantities such as the basic reproduction number should in theory be defined as the strain level. Strain composition may differ between the US and Canada (and potentially also between regions/provinces), such that the observed differences in estimates might potentially partly reflect differences in strain composition. Understanding the complex RV population-level dynamics and immunity landscape would greatly benefit from a better characterization of this underlying diversity. This underscores the need for improved and continuous measurement of RV epidemiology and genomics, notably through sequencing and seroprevalence studies. Co-circulation of numerous subtypes of the three species is common [71, 72], with subtype prevalence and age patterns appearing to remain stable over decades [73]. RV phylogenetic analyses are scarce. Still, in [74, 6], RV infections are characterized by many *localized subtype-specific ‘mini-epidemics’*, with multiple co-circulating RV subtypes and changes over time within a social structure (e.g., school, hospital); [75, 76] also reported a high genotype diversity throughout the year (especially RV-A and C), with significant seasonal variation in [76] (tropical climate), and pandemic levels comparable to pre-pandemic in [75]. Other studies observed a shift in dominance before and during the pandemic, e.g., [72].

Third, we focused on the potential viral interference of IAV on RV, but we did not examine the opposite direction which is also supported by several studies (e.g., [31, 18, 20, 22]). The resilience of RV during the COVID-19 pandemic has notably been hypothesized to have contributed to a prolonged suppression of IV circulation [50] and, more generally, shaped the spread of IV [11]. Here, fitting the model to IAV poorly captured the data, indicating that further work is needed to better tailor the model to IAV biology (e.g., antigenic changes, seeding and subtype interference). Revisiting data of the 2009 flu A pandemic in Europe – where the interference of RV on IAV was initially hypothesized [28, 29, 30] – may provide further insights into these potential interactions. RV may also interact with other viruses (including SARS-CoV-2) [11, 77, 78, 79, 19, 27].

We estimated key parameters of RV population dynamics, yet substantial gaps remain in our understanding of the underlying drivers, underscoring the need for better continuous epidemiological and genomic surveillance. More broadly, understanding pathogen-pathogen interactions is an important emerging frontier in public health, with implications for the prediction and anticipation of future outbreaks. Most studies, including this work, have focused on pairwise pathogen interactions, whereas real-world pathogen ecology may be shaped by more complex polymicrobial/multistrain interaction networks. Identifying when, where and how pathogen-pathogen interactions influence infectious disease dynamics will thus be essential in the future.

## 4 Materials and Methods

### 4.1 Data

We collected time series of weekly detections for RV/EV and IAV in the US (**Fig. S2**) and Canada (**Fig. S3**). US data were collected from the Centers for Disease Control and Prevention (CDC), as reported through the National Respiratory and Enteric Virus Surveillance System (NREVSS). Data were downloaded at both the national and regional level (10 Health and Human Services (HHS) regions, **Fig. S1**) and range from January 4, 2014 to June 28, 2025 (regional level) or September 6, 2025 (national level). Canadian time series were reconstructed from publicly available reports provided by the Respiratory Virus Detection Surveillance System (RVDSS, Public Health Agency of Canada [80]). We analyzed data at the national and regional level – 4 provinces/regions: Atlantic, British Columbia, Ontario and Prairies (which includes Alberta, Manitoba and Saskatchewan) (**Fig. S1**) – from August 31, 2013 to June 8, 2024. NREVSS and RVDSS are decade-long laboratory-based surveillance systems that monitor respiratory virus activity across the US and Canada, respectively. These systems involve a network of participating laboratories across each country testing patients with respiratory symptoms in outpatient and inpatient settings. Testing is conducted by PCR; in recent years the use of multiplex PCR has become more frequent. We note that rapid laboratory diagnostic testing does not differentiate RVs from other EVs (and, *a fortiori*, does not distinguish RV species and subtypes), and results are typically reported as a combined RV/EV category. While there is no strong evidence for viral interference between IAV and other EVs, RVs are expected to account for most detections, in line with [56, 57]. We therefore considered RV/EV patterns to be primarily driven by RV circulation and, for brevity, we refer to RV/EV simply as RV.

Positivity rates (i.e., proportion of detections among tests performed) can yield a biased measure of pathogen activity levels depending on the co-circulation of other respiratory pathogens [81, 58]. We thus calculated an incidence proxy to mitigate this bias and to account for changes in testing effort over time. Following an approach similar to [82, 55, 42], we rescaled incidence data by multiplying raw cases by a weekly testing factor equal to the average number of tests (over the entire studied period) divided by the one-year moving average of the number of tests (see testing patterns in **Fig. S4** for RV and **S5** for IAV).

We also used Google COVID-19 Community Mobility Reports [83] as a proxy to capture reductions in transmission due to COVID-19 pandemic NPIs. Google mobility Reports provide percentage changes in the number of visitors relative to baseline days (median value from the 5-week period from January 3 to February 6, 2020). As in [58, 55], we defined a weekly mobility metric, *c*, by averaging mobility data across four categories (grocery & pharmacy, retail & recreation, transit stations and workplaces), computing a population-weighted average across states or provinces. Google mobility data were only reported between February 15, 2020 and October 15, 2022 and we assumed *c* = 0 (i.e., same as baseline) outside of this range (we relaxed this assumption in **Fig. S22**). We approximated population sizes using 2021 census data for Canada [84] and 2020 census data for USA [85].

### 4.2 Transmission model

We modeled RV epidemiological dynamics using a seasonally-forced SIRS model (details of notation are provided in **Table 1**). We assumed a homogeneously mixed population in which susceptible hosts (*S*) become infected and infectious (*I*) at a *per capita* rate *λ*(*t*) (force of infection, where *t* denotes the current time). Infected hosts then enter the recovered state (*R*) at a *per capita* rate *γ*. Infection provides full immunity against reinfection but immunity naturally wanes at a *per capita* rate *ω* after recovery (in the following we denote Ω the mean duration of immune protection, i.e., the mean sojourn time in *R*). Last, we define *µ* as the *per capita* host birth and death rate, such that the population size *S*(*t*) + *I*(*t*) + *R*(*t*) = *N* remains constant over time. These epidemiological trajectories can be written as the following system of ordinary differential equations (ODEs):

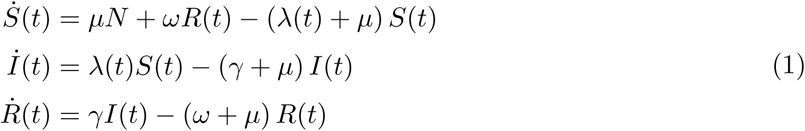

where the current incidence I(*t*) is computed by tracking the number of new infections *λ*(*t*)*S*(*t*). As with NPIs, potential effect of IAV is assumed to affect susceptibility or transmissibility, modeled phenomenologically as a scalar effect on the transmission term. The force of infection *λ* is then given by:

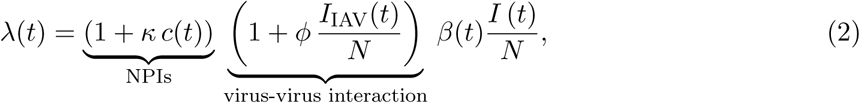

where *β*(*t*) is the seasonal transmission rate, *κ*, the strength of the effect of NPIs (assuming this effect scales linearly with the changes in contacts/visitors, *c*(*t*), due to control measures), and where *ϕ* captures the strength and direction of the effect of IAV (assuming this effect scales linearly with IAV prevalence, *I*_IAV_(*t*)*/N*).

For the purpose of model fitting, we use in practice the efficient discretization scheme described in [86]:

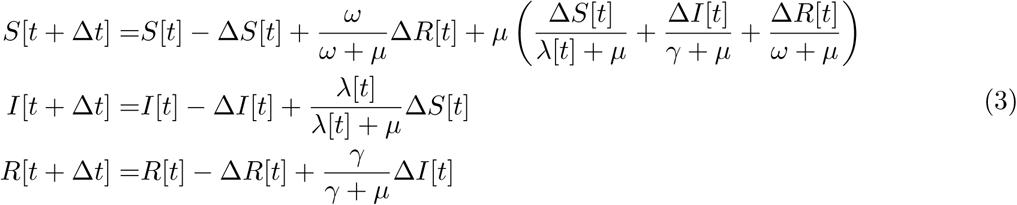

Δ*S*[*t*], Δ*I*[*t*] and Δ*R*[*t*] represents the density of individuals leaving compartment *S*, *I* and *R*, respectively, between time *t* and *t* + Δ*t*:

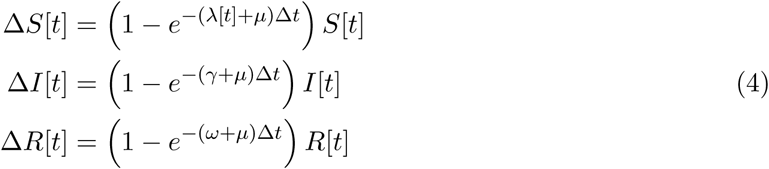

where 1 − exp (−*r*Δ*t*) is the probability of leaving a given compartment when *r* is the sum of rates out of this compartment in the ODE model. Consequently, the corresponding mean sojourn time is given by Δ*t/* (1 − exp(−*r*Δ*t*)), which, taking the limit as Δ*t* → 0, yields the sojourn time in the corresponding ODE, 1*/r*. Besides, the incidence I (i.e., number of new cases per time step) now corresponds to the term:

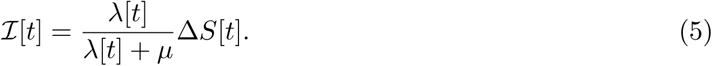

Last, we determine the effective reproduction number R (i.e., the expected number of secondary infections by a single infectious individual), as well as the basic reproduction number *R*_0_ (defined in an otherwise wholly susceptible population), of the discrete model (3). Multiplying the incidence I[*t*] by the mean number of time steps an individual stays in the the infected compartment, 1*/*(1 − exp(−(*γ* + *µ*)Δ*t*)), and linearizing around *I*[*t*] = 0 yields:

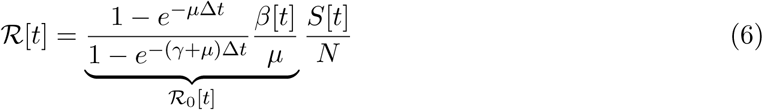

(we drop the potential effects of NPIs and viral interaction). Note that *R*_0_ converges towards *β*[*t*]/(*γ* + *µ*) when Δ*t* → 0.

### 4.3 Bayesian inference

Using a Bayesian framework, we fitted the model (3)-(4) to our RV detection proxy *C*[*t*]. Because *C*[*t*] is a positive, continuous variable, we relied on a log-Normal likelihood:

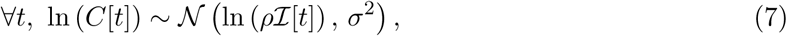

where I[*t*] is the incidence given in equation (5), *ρ* ∈ [0, 1] accounts for under-reporting and *σ* is the residual standard deviation. The log-Normal distribution implies multiplicative observation noise. Poisson and negative binomial likelihoods are also commonly used in similar epidemiological settings, but require discrete case counts.

To estimate the seasonal transmission rate *β*[*t*], we used 52 weekly transmission rates *β*_k_ (with *k* ∈ [1, 52]], the *k*^th^ week of the year), modeled as a cyclical random-walk on the log scale:

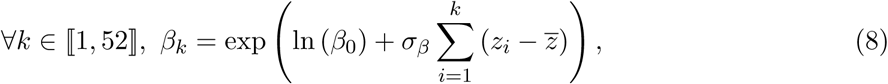

with *z_i_*, some independent step increments (centered by their arithmetic mean *z* to enforce cyclicity), whose magnitude is scaled by *σ_β_*, and where *β*_0_ is a reference transmission rate (by construction, *β*_52_ = *β*_0_).

IAV incidence was directly input from surveillance data, similarly to [59]. Specifically, we substituted the unknown IAV prevalence in the viral interaction term (1 + *ϕ I*_IAV_[*t*]*/N*) by the observed weekly IAV incidence, *C*_IAV_, which was assumed to be a reasonable proxy as IAV infectious period is also of the order of one week. Values were slightly smoothed using a geometric three-week moving average to reduce the influence of observation noise. We then scaled the time series between 0 and 1 by dividing by its global maximum (across all seasons of the study period) to enforce a lower bound of -1 for *ϕ*, ensuring that the force of infection is always positive while preserving any relative differences across seasons such as epidemic size. When fitting the model to real data, we also considered a lagged sum of IAV incidence instead of current incidence, substituting *C*_IAV_[*t*] by 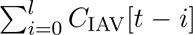 with a lag *l* of 1, 3 or 5 weeks. This approach assumes a more sustained effect, where individuals infected during the past *l* weeks remain protected, and is intended to approximate varying durations of potential immune protection or reduced susceptibility. Interaction mechanisms mediated by interferon responses (typically resolve within days to a week) are more consistent with short-term effects (lags 0 or 1), whereas ecological mechanisms could potentially last longer.

We fitted models with a weekly time step (Δ*t* = 1 week) using a Hamiltonian Monte Carlo algorithm (no-U-turn sampler) implemented in Stan [87] and its R interface `rstan` [88]. RV infections are typically associated with an infectious period between 1 or 2 weeks [3, 89, 90, 62], so we assumed infected individuals to recover at rate *γ* = 2 week*^−^*^1^, that is a mean infectious period of 1*/* (1 − exp(−2)) ≈ 1.16 week (around 8 days). We estimated the rate of immune waning *ω* because of lack of information, noting that elapsed time between two infections of the same individual is likely to be relatively short due to the wide antigenic diversity of RVs and the low level of cross-immunity [69, 4]. We also fixed the mortality rate, assuming *µ* = 1*/*80 year*^−^*^1^. Let *θ* be the vector of parameters to estimate and 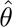 its estimator. Using the seasonal transmission rate (8), we have 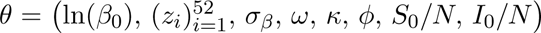; priors can be found in **Table S2**. For each estimation process, we independently ran 4 MCMC chains of 4,000 iterations (including a burn-in period of 2,000 iterations). We assessed the convergence of posterior distributions by the absence of warnings from Stan, including no divergent transitions, sufficient maximum tree depth, Gelman-Rubin statistics (R-hat) below 1.05 and effective sample size (ESS) above 400 (100 × number of chains) for each parameter.

### 4.4 Simulation study

We conducted a simulation study to validate our approach, and in particular our ability to quantify pathogen-pathogen interaction. Simulated data were generated by extending the ODE model (1) to account for two pathogens (**supporting information §S2**). ODEs were solved using the deSolve package [91] in the R programming language [92]. We considered 3 main scenarios:

*•* **Scenario 1 (bimodal seasonality with no interaction):** RV seasonal forcing peaks twice a year, and transmission is independent from IAV (*ϕ* = 0);
*•* **Scenario 2 (“masked interaction”):** Building upon the previous scenario, IAV now negatively impacts RV (*ϕ <* 0), but we adjust RV seasonal forcing so that its force of infection mimics scenario 1 when IAV is at its endemic attractor (i.e., in the absence of perturbation);
*•* **Scenario 3 (unimodal seasonality with negative interaction):** RV seasonality follows a simple sinusoidal forcing (one peak per year), and the two annual RV outbreaks arise from the interference by IAV (*ϕ <* 0).

Seasonal transmission profiles are shown in **Fig. 2-AB** and parameter values are listed in **Table S1**; virus parameters are based on existing literature and/or to reproduce the observed epidemiological characteristics of RV and IAV (see caption for more details).

In the last two scenarios, the maximum change in transmission due to viral interference lies around −35%. Starting from initial conditions on the endemic attractor of both pathogens, we simulated the model for 10 years, corresponding to a 6-year pre-pandemic period (endemic limit cycles) followed by a 4-year (post-)pandemic period. We used Google mobility data from Canada to model more realistically the pandemic period. We also considered two extensions: (i) one with a 6-month shift in the timing of NPIs (e.g., NPIs in winter now occurs in the summer), and (ii) one where we introduced a one-off exogenous perturbation in IAV dynamics (a “kick”, moving 35% of the susceptibles to IAV to the recovered compartment) at *t* = 1.5 year during the pre-pandemic period. All simulations are presented in **Fig. S8**.

Simulated pathogen detections were generated using a log-Normal distribution with a residual standard deviation *σ* of 0.25 on the log scale. We fitted the model (3) to RV detections using the seasonal transmission (8) and compared the results with those obtained when the model was only fitted to the pre-pandemic period. Additionally, we also contrasted these fits with a null model with no interaction (fixing *ϕ* = 0). To account for model discretization, the recovery rate was fixed as − ln (1 − *γ*Δ*t*) */*Δ*t*, where *γ* is the recovery rate used in the continuous ODE model. Likewise, we did not directly compare the true and estimated rates of immune waning *ω*, but the mean durations of immune protection Ω: 1*/ω* (continuous ODE model) vs. Δ*t/* (1 − exp(−*ω*Δ*t*)) (discretized version).

## Data availability statement

Data and codes are publicly available at: https://github.com/WakinyanB/Rhinovirus_dynamics.

## Supporting information

S1 File. Supplementary notes, figures (Figs. S1 to S27) and tables (Tables S1 to S3) referenced in the main text.

## Author contributions

**Conceptualization:** Wakinyan Benhamou, Bryan T. Grenfell, C. Jessica E. Metcalf.

**Methodology:** Wakinyan Benhamou, Emily Howerton, Sang Woo Park, Bryan T. Grenfell.

**Data:** Wakinyan Benhamou, Sang Woo Park, Ćecile Viboud.

**Investigation:** Wakinyan Benhamou, Emily Howerton, Sang Woo Park, Ćecile Viboud, C. Jessica E. Metcalf, Bryan T. Grenfell.

**Formal analysis:** Wakinyan Benhamou.

**Visualization:** Wakinyan Benhamou, Emily Howerton, Bryan T. Grenfell.

**Supervision:** Bryan T. Grenfell, C. Jessica E. Metcalf.

**Writing – original draft:** Wakinyan Benhamou.

**Writing – review & editing:** Wakinyan Benhamou, Emily Howerton, Sang Woo Park, Ćecile Viboud, C. Jessica E. Metcalf, Bryan T. Grenfell.

## Supporting information

### S1 Cross-wavelet transform analysis

We performed cross-wavelet transform analysis to examine the relationships in time-frequency space between rescaled RV/EV case detections (*C*) and rescaled IAV case detections (*C*_IAV_).

Let (*x_t_*, *t* = 1, . . ., *N*) be a time series. Its continuous wavelet transform is given by:

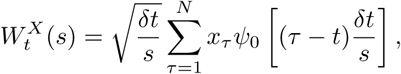

with *s*, the wavelet scale, *δt*, the time step, and *ψ*_0_, the mother wavelet (Morlet wavelet by default) [1]. The cross-wavelet transform of two time series *x_t_* and *y_t_* is then given by:

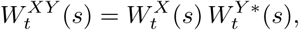

where * denotes complex conjugation. Cross-wavelet power is defined as the corresponding modulus, 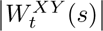, and represents the local strength of common power between the two time series in time-frequency space, while the complex argument yields their local relative phase [1].

In practice, we use the function `xwt` from the R package `biwavelet` [2]. Time series were first square-root transformed to stabilize variance and standardized to ensure comparability between time series prior to cross-wavelet analysis, that is:

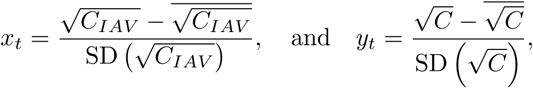

where the overline denotes the corresponding arithmetic mean and SD, the standard deviation. The cross-wavelet power was standardized to facilitate comparison across locations. Specifically, the standardized cross-wavelet power was calculated as log_2_ 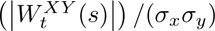, where *σ_x_* and *σ_y_* are the standard deviations of the standardized input time series *x_t_* and *y_t_*, respectively. Results were used to confirm and complement the initial visualization and description of the time series (i.e., the asynchronous dynamics of IAV and RV/EV, see **Fig. 1**), further motivating the subsequent analyses.

### S2 Two-pathogen transmission model

We used the seasonally-forced SIRS model (1)-(2) to simulate RV epidemiological dynamics, using IAV incidence as an external input. In order to generate synthetic data in the simulation study, we extended this model to explicitly simulate IAV dynamics (denoted by the subscript IAV) as follows:

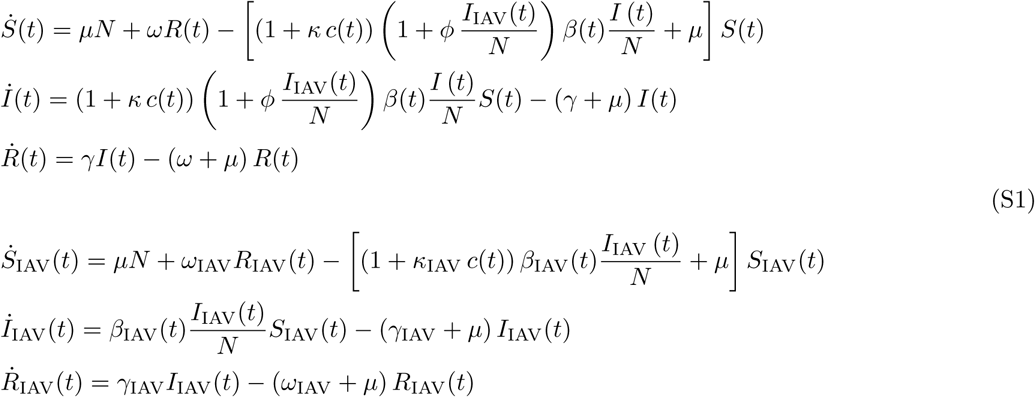

where IAV transmission rate *β*_IAV_(*t*) is modeled as a standard sinusoidal function:

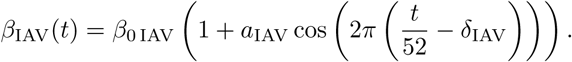

We listed parameter values in **Table S1**.

### S3 Joint model fit across all locations

In the main text, we fitted the model either independently to each region/province or to data aggregated at the national level. We additionally fitted the model simultaneously to all regions/provinces, allowing to constrain selected parameters across locations. Specifically, we assumed that the rate of immune waning *ω* was identical across all locations and that the viral interaction parameter *ϕ* was identical across locations within a given country (US vs. Canada). In our framework, the latter implicitly assumes that reporting is sufficiently similar across locations within a country, such that relative differences in IAV incidence reflect true differences in viral circulation between locations rather than spatial variation in reporting effort. In practice, both spatial heterogeneities in IAV circulation and reporting effort are likely to exist, but we consider here this assumption to be more plausible at the country scale than across different countries. Within each country (US vs. Canada), IAV incidence was thus scaled between 0 and 1 using the global maximum of IAV incidence across all seasons and corresponding locations. The model structure is otherwise similar to that presented in the main text, with the exception of the prior of the location-specific initial conditions, (*S*_0,*i*_, *I*_0,*i*_, *R*_0,*i*_), for which we specified a Dirichlet(35, 1, 4) distribution.

### S4 Supplementary figures

**Figure S1:**
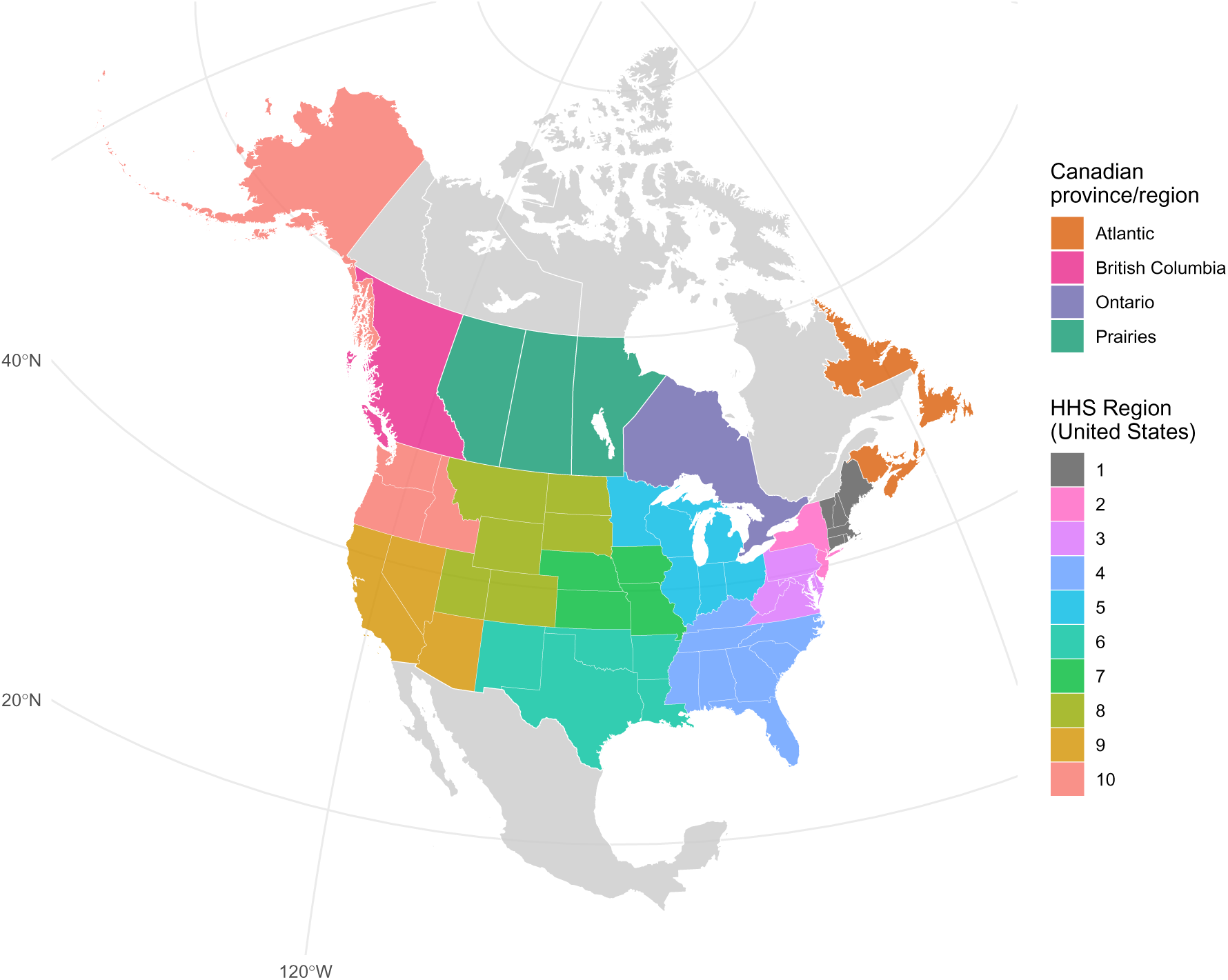
Colored map of the studied regions. We focused here on the US and Canada. At the subnational level, we considered 4 provinces/regions for Canada – Atlantic, British Columbia, Ontario and Prairies – as well as the 10 Health and Human Services (HHS) regions for the US. Not displayed: HHS region 2 also includes Puerto Rico and the Virgin Islands; HHS region 9 also includes Hawaii, American Samoa, the Commonwealth of the Northern Mariana Islands, the Federated States of Micronesia, Guam, the Marshall Islands and the Republic of Palau. Base layer of the map were obtained from Natural Earth (https://www.naturalearthdata.com/) via the `rnaturalearth` R package [3].

**Figure S2:**
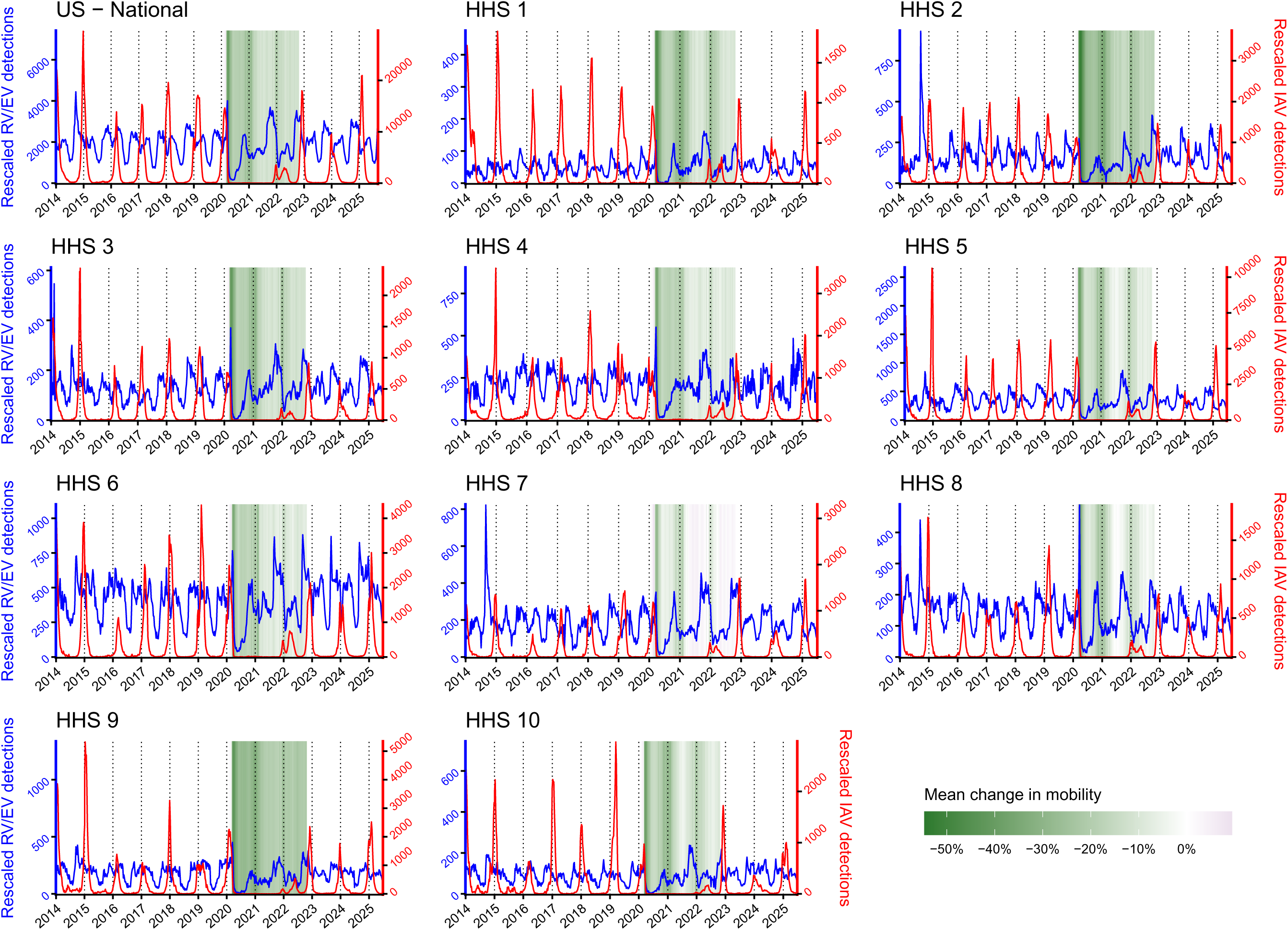
RV and IAV time series (US). RV/EV (blue) and IAV (red) detections in the US (national and 10 HHS regions). Data were rescaled to account for changes in testing patterns (see **Fig. S4** and **S5**). We plot time series from the Centers for Disease Control and Prevention (CDC), as reported through the NREVSS. Mean change in mobility (colored background) during the COVID-19 pandemic were computed from [4].

**Figure S3:**
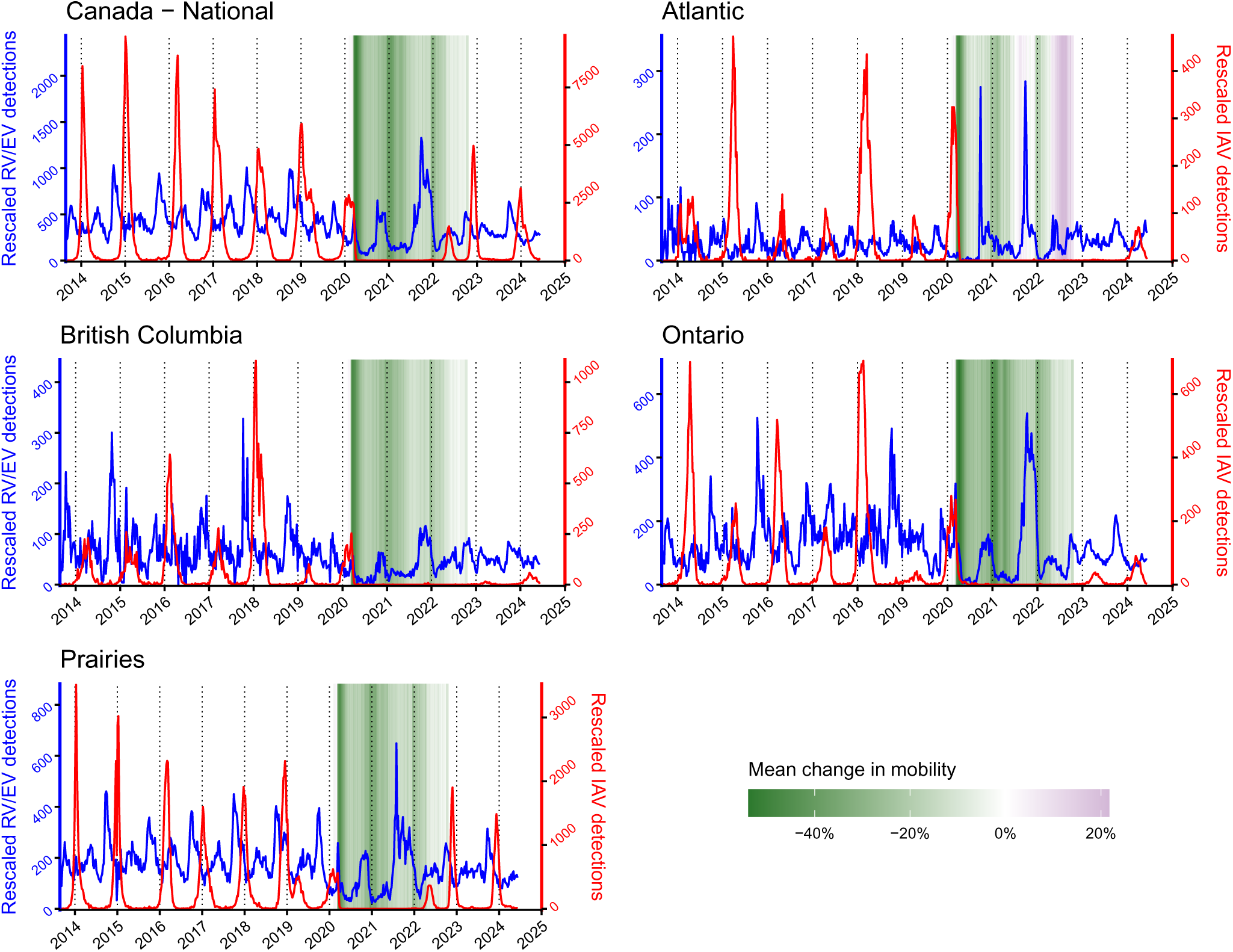
RV and IAV time series (Canada). RV/EV (blue) and IAV (red) detections in Canada (national and 4 provinces). Data were rescaled to account for changes in testing patterns (see **Fig. S4** and **S5**). Time series were reconstructed from publicly available reports by the RVDSS. Mean change in mobility (colored background) during the COVID-19 pandemic were computed from [4].

**Figure S4:**
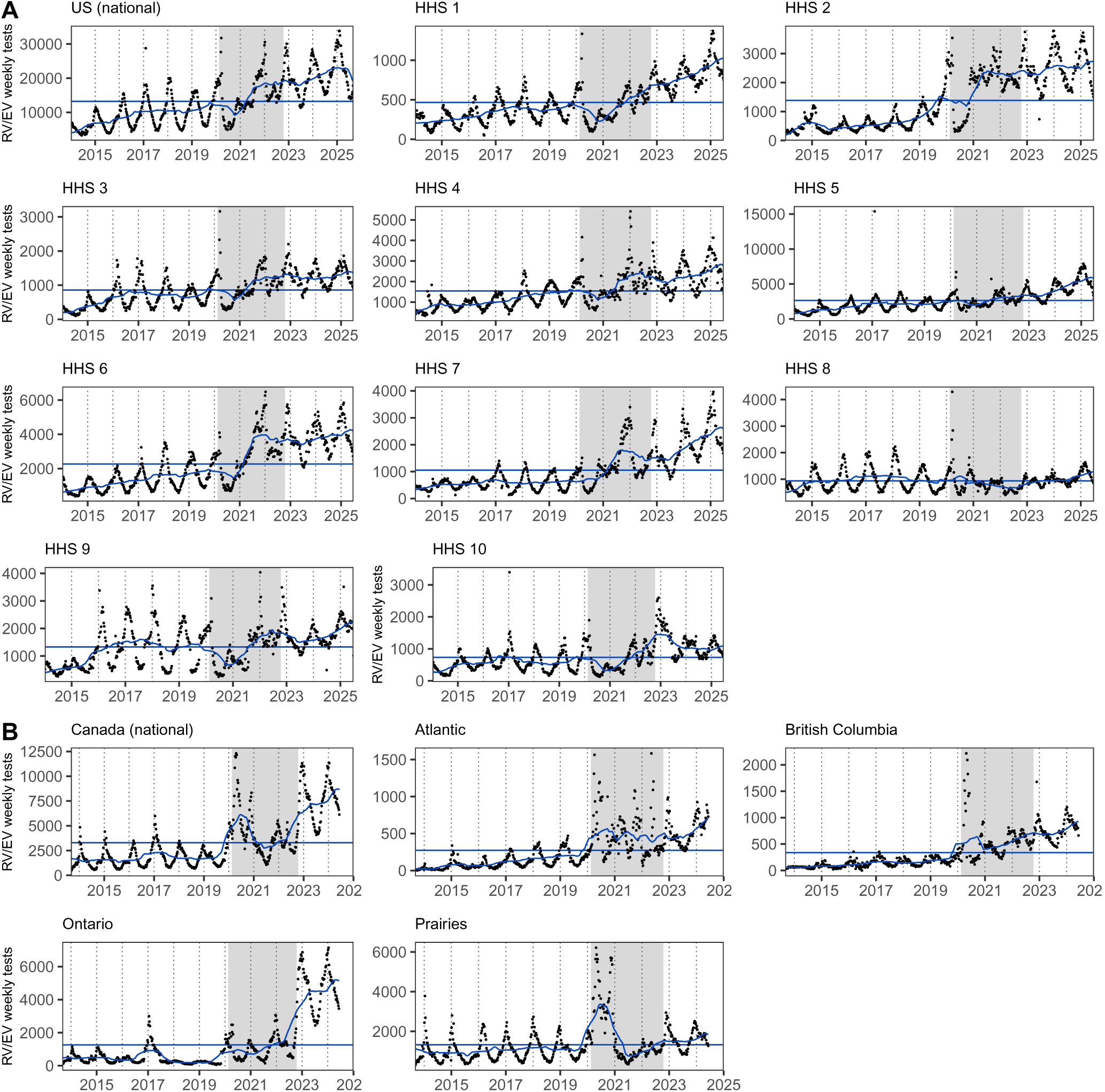
RV testing patterns. Weekly number of RV/EV tests (black points) in (A) the US and (B) Canada. Blue lines correspond to the 1-year moving average and mean number of tests over the studied period. To account for testing behavior, we rescaled incidence data by multiplying raw cases by a weekly testing factor equals to the average number of tests (over the entire studied period) divided by the one-year moving average of the number of tests. Grey backgrounds indicate the pandemic period.

**Figure S5:**
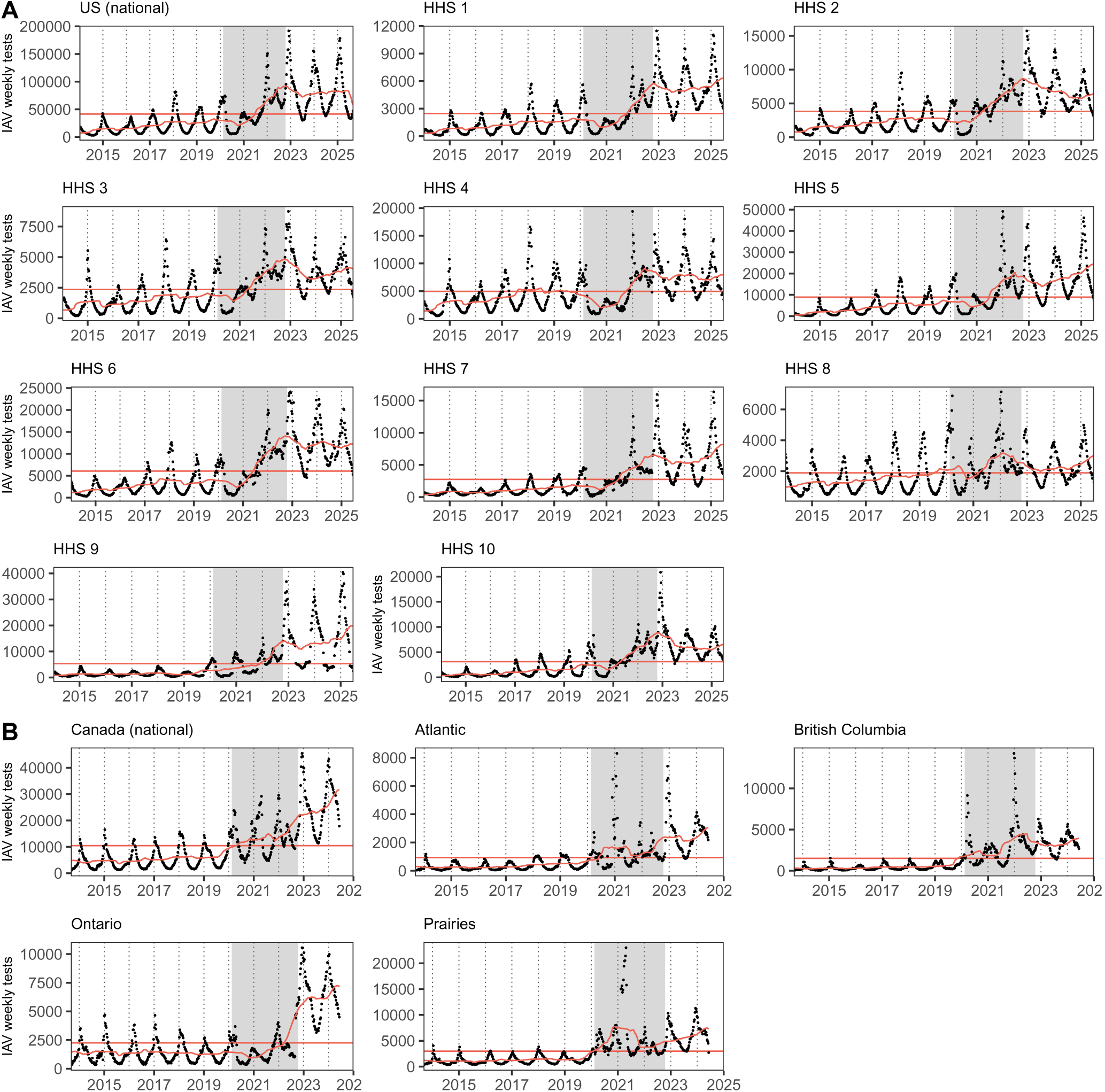
IAV testing patterns. Weekly number of IAV tests (black points) in (A) the US and (B) Canada. Red lines correspond to the one-year moving average and mean number of tests over the studied period. To account for testing behavior, we rescaled incidence data by multiplying raw cases by a weekly testing factor equals to the average number of tests (over the entire studied period) divided by the one-year moving average of the number of tests. Grey backgrounds indicate the pandemic period.

**Figure S 6:**
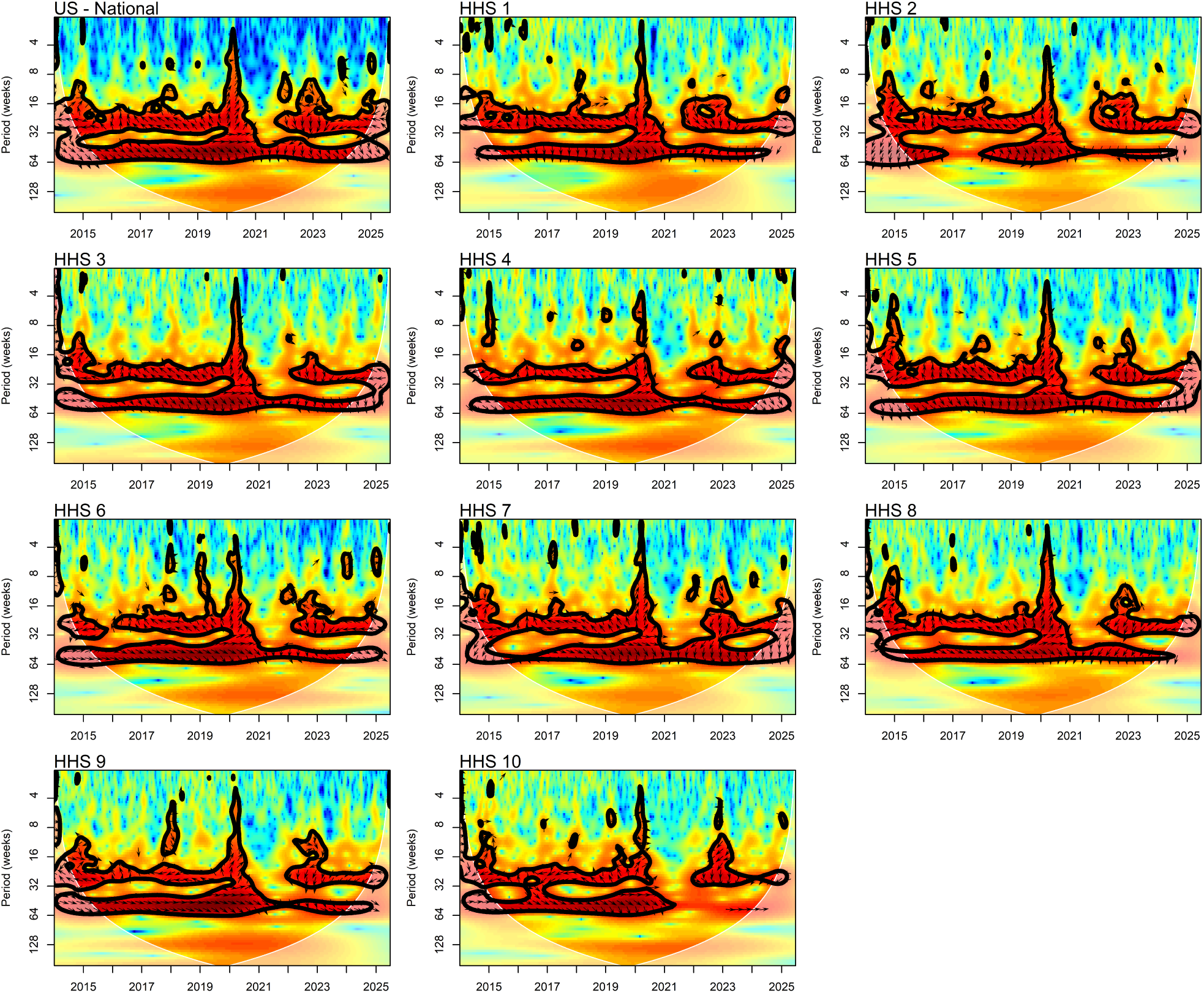
Cross-wavelet transform of IAV and RV detections in the US. We used the function xwt from the R package biwavelet [2] (see details in **SI Appendix §S1**). Colors indicate cross-wavelet power, with significant regions (95% confidence level) outlined by a contour. Arrows pointing right (resp. left) suggest IAV and RV detections are in phase (reps. out-of-phase) and arrows pointing up (resp. down) suggest IAV detections lead (resp. lag) RV detections (with vertical arrows indicating a phase difference of *π*/2 (quarter cycle)).

**Figure S7:**
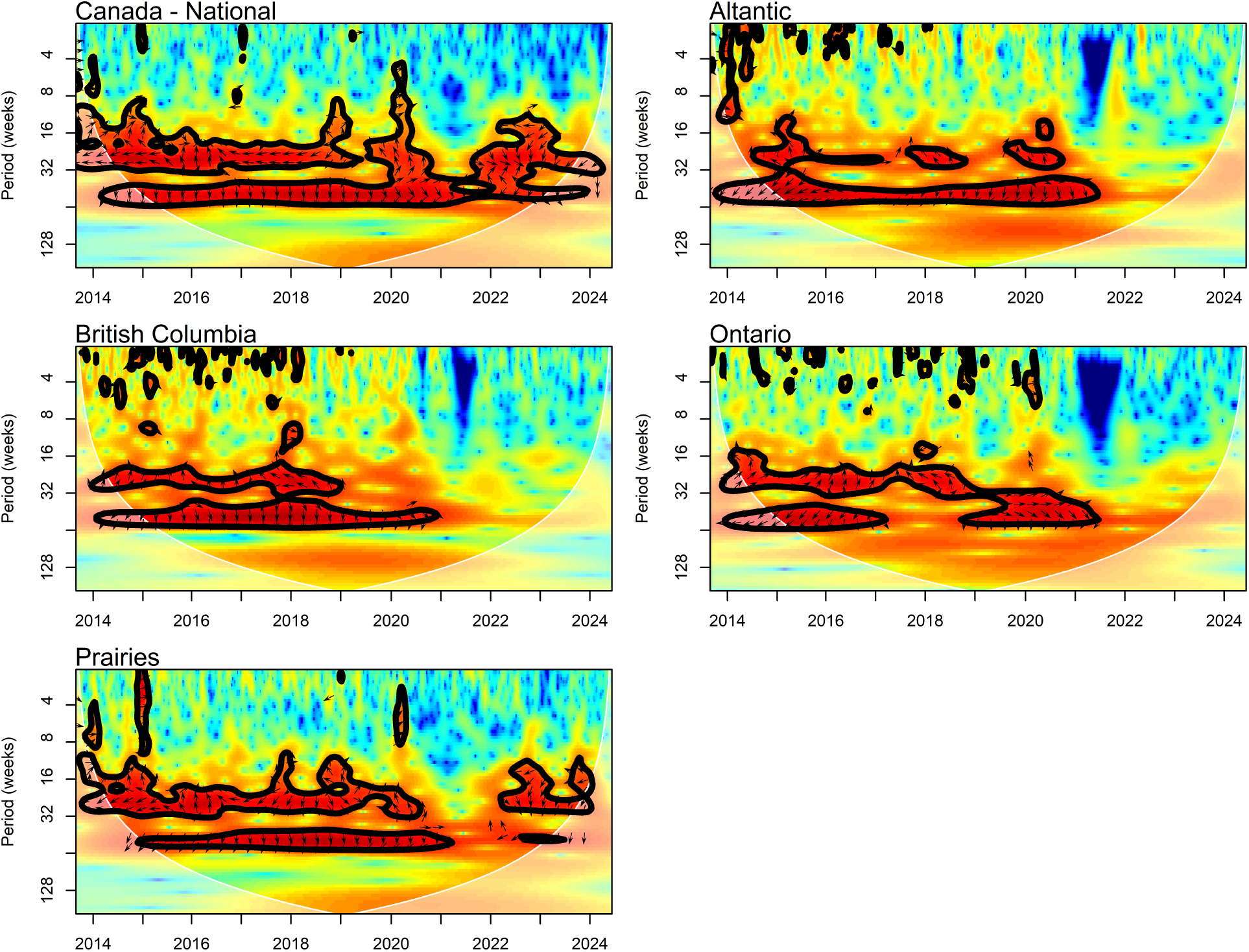
Cross-wavelet transform of IAV and RV detections in Canada. We used the function xwt from the R package biwavelet [2] (see details in **SI Appendix §S1**). Colors indicate cross-wavelet power, with significant regions (95% confidence level) outlined by a contour. Arrows pointing right (resp. left) suggest IAV and RV detections are in phase (reps. out-of-phase) and arrows pointing up (resp. down) suggest IAV detections lead (resp. lag) RV detections (with vertical arrows indicating a phase difference of *π*/2 (quarter cycle)).

**Figure S8:**
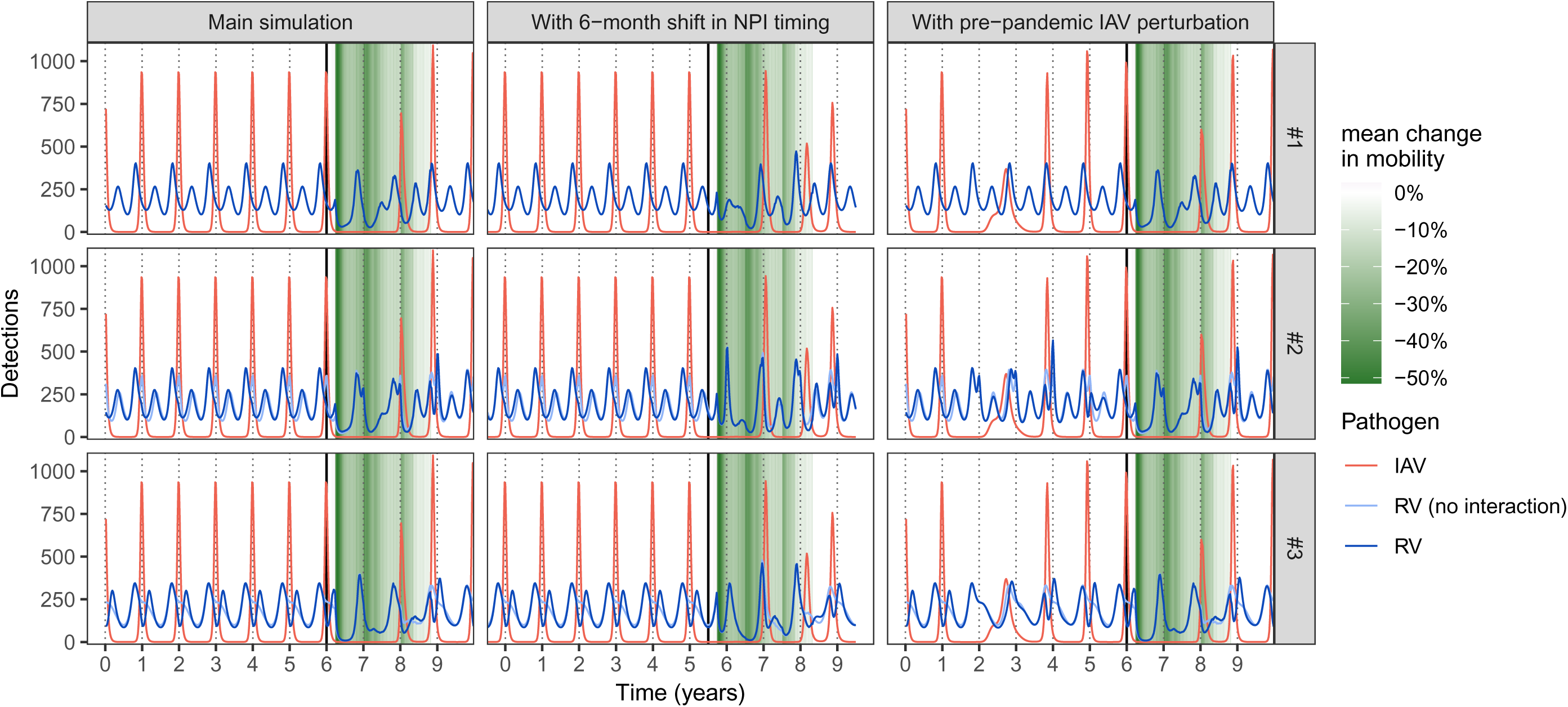
Simulated dynamics. We simulated RV (dark blue lines) and IAV dynamics (red lines) using model (S1) and parameter values listed in **Table S1**. We consider three main scenarios: 1, RV and IAV are independent (*ϕ* = 0); 2, IAV negatively impacts RV (*ϕ* < 0), but we adjust RV seasonal forcing so that its force of infection mimics that from scenario 1 when IAV is at its endemic attractor; 3, simple sinusoidal changes to RV transmission rate, with IAV negatively impacting RV (*ϕ* < 0). See seasonal transmission profiles of RV and IAV in **Fig. 2-AB**. For scenarios 2 and 3, we also plot RV dynamics if there was no interaction due to IAV (light blue lines). In the first column (main simulation), we start from initial conditions on the endemic attractor of both pathogens, then simulate the model for 10 years, corresponding to a 6-year pre-pandemic period (stationary dynamics) followed by a 4-year (post-)pandemic period (vertical solid lines indicate the separation between the two periods). We used Google mobility data from Canada for the pandemic period. In the second column, we shift NPI timing 6-month. In the third column, we introduce a one-off exogenous perturbation perturbation in IAV dynamics (moving 35% of S_IAV_ to R_IAV_) at t = 1.5 year during the pre-pandemic period.

**Figure S9:**
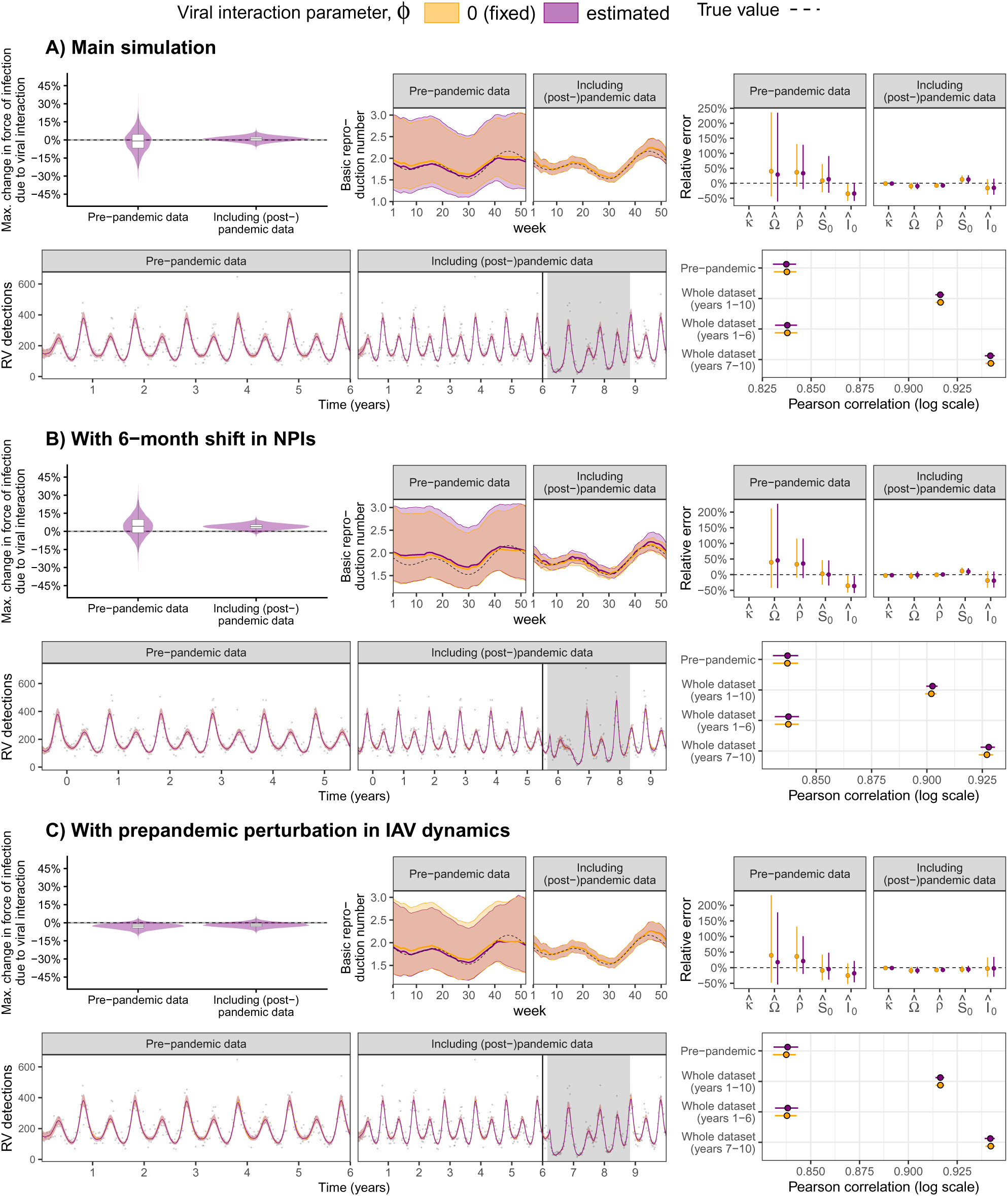
Simulation study estimation results (scenario 1). Here, RV and IAV are independent (**ϕ** = 0). For each panel (A-C), we plot, from top to bottom and from left to right: (i) estimated effect of IAV (posterior distributions); (ii) estimated basic reproduction number of RV (median and 95% CrIs); (iii) relative errors of the remaining estimated parameters (median and 95% CrIs); (iv) simulated data (black points) and fitted values (median and 95% CrIs), where the vertical line marks the transition between preand (post-)pandemic periods and gray backgrounds, the period of implementation of COVID-19 NPIs; and (v) Pearson correlation coefficients (median and 95% CrIs) between simulated and fitted values on the log scale. True values are indicated by dashed lines. We fitted models assuming no viral interaction (**ϕ** = 0, in yellow) or estimating the interaction term **ϕ** (purple). See **Fig. S8** for more details about the simulations.

**Figure S10:**
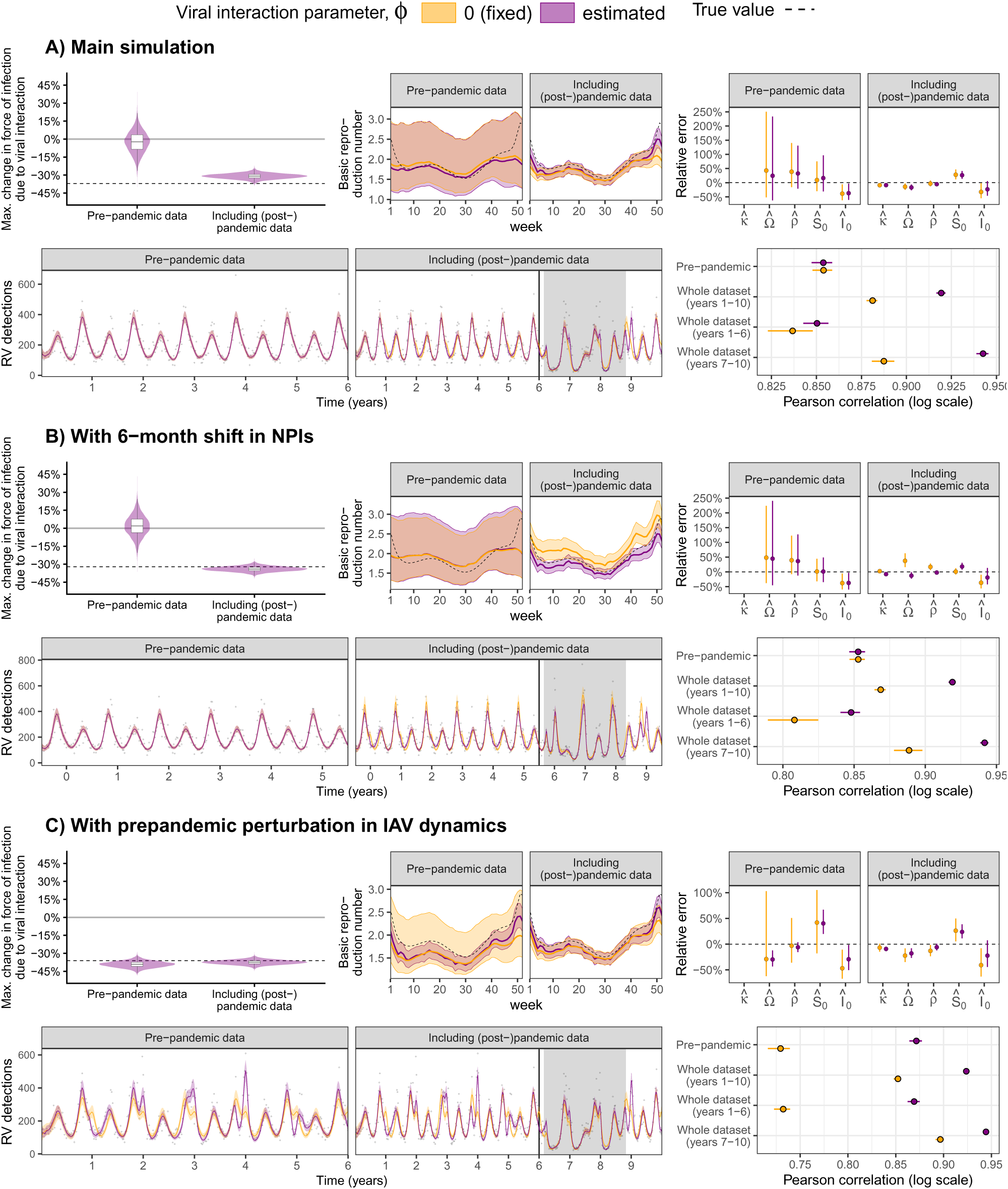
Simulation study estimation results (scenario 2). Here, IAV negatively impacts RV (**ϕ** < 0). For each panel (A-C), we plot, from top to bottom and from left to right: (i) estimated effect of IAV (posterior distributions); (ii) estimated basic reproduction number of RV (median and 95% CrIs); (iii) relative errors of the remaining estimated parameters (median and 95% CrIs); (iv) simulated data (black points) and fitted values (median and 95% CrIs), where the vertical line marks the transition between preand (post-)pandemic periods and gray backgrounds, the period of implementation of COVID-19 NPIs; and (v) Pearson correlation coefficients (median and 95% CrIs) between simulated and fitted values on the log scale. True values are indicated by dashed lines. We fitted models assuming no viral interaction (**ϕ** = 0, in yellow) or estimating the interaction term **ϕ** (purple). See **Fig. S8** for more details about the simulations.

**Figure S11:**
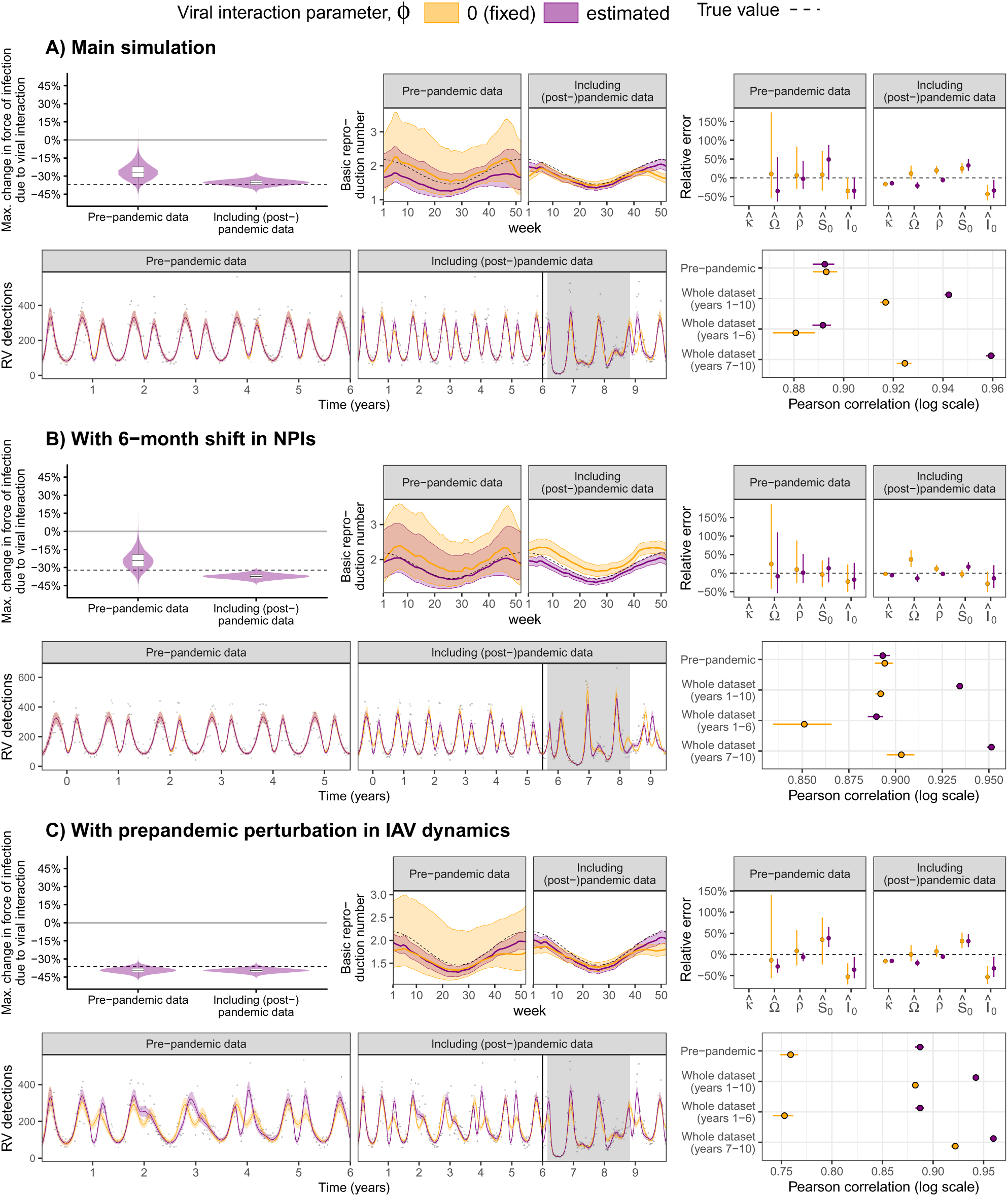
Simulation study estimation results (scenario 3). Here, IAV negatively impacts RV (*ϕ* < 0). For each panel (A-C), we plot, from top to bottom and from left to right: (i) estimated effect of IAV (posterior distributions); (ii) estimated basic reproduction number of RV (median and 95% CrIs); (iii) relative errors of the remaining estimated parameters (median and 95% CrIs); (iv) simulated data (black points) and fitted values (median and 95% CrIs), where the vertical line marks the transition between preand (post-)pandemic periods and gray backgrounds, the period of implementation of COVID-19 NPIs; and (v) Pearson correlation coefficients (median and 95% CrIs) between simulated and fitted values on the log scale. True values are indicated by dashed lines. We fitted models assuming no viral interaction (*ϕ* = 0, in yellow) or estimating the interaction term *ϕ* (purple). See **Fig. S8** for more details about the simulations.

**Figure S12:**
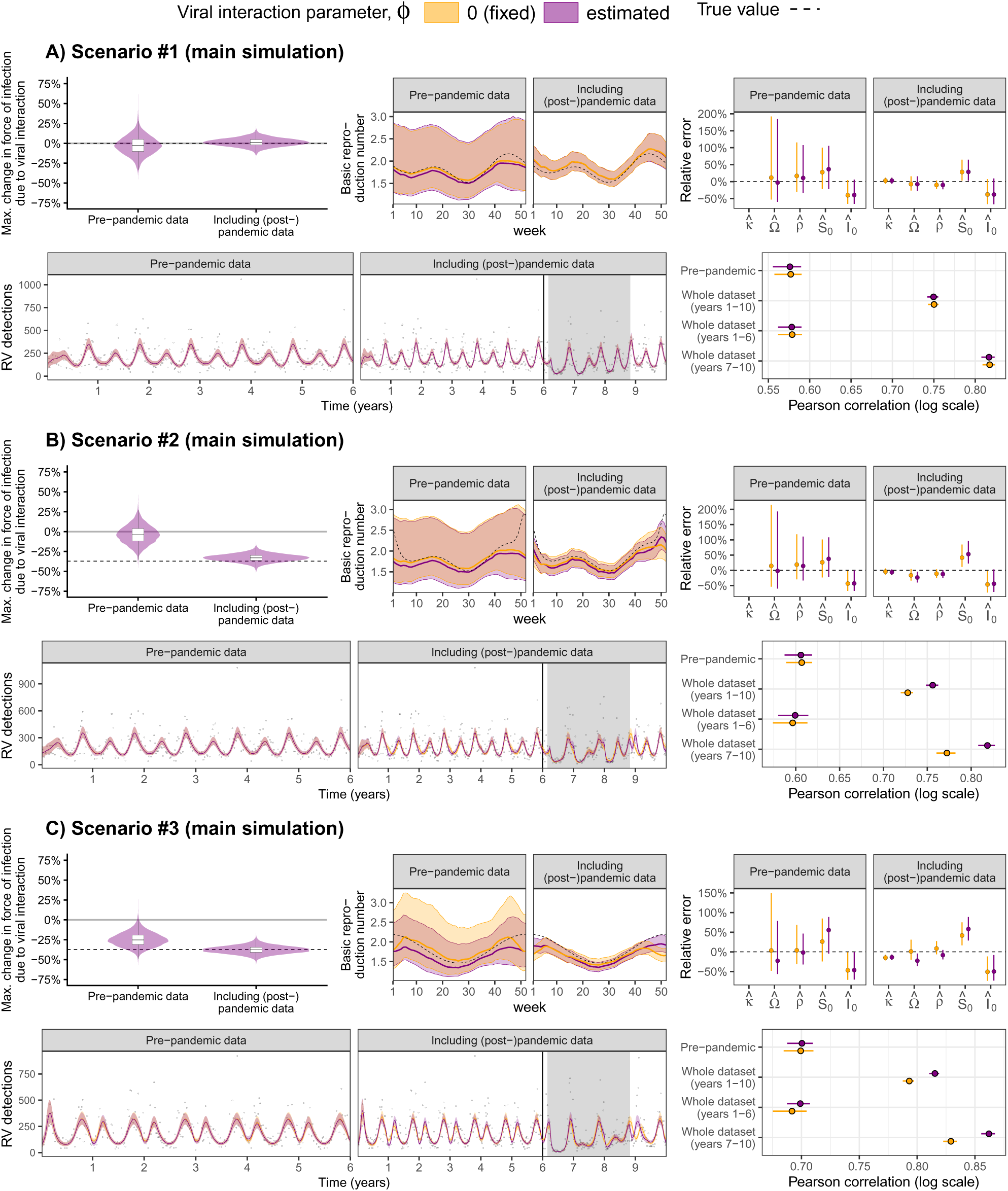
Simulation study estimation results with high observation noise. We assessed the robustness of our results to the observation noise, generating simulated data with σ = 0.5 (instead of σ = 0.25). For each panel (A-C), we plot, from top to bottom and from left to right: (i) estimated effect of IAV (posterior distributions); (ii) estimated basic reproduction number of RV (median and 95% CrIs); (iii) relative errors of the remaining estimated parameters (median and 95% CrIs); (iv) simulated data (black points) and fitted values (median and 95% CrIs), where the vertical line marks the transition between preand (post-)pandemic periods and gray backgrounds, the period of implementation of COVID-19 NPIs; and (v) Pearson correlation coefficients (median and 95% CrIs) between simulated and fitted values on the log scale. True values are indicated by dashed lines. We fitted models assuming no viral interaction (*ϕ* = 0, in yellow) or estimating the interaction term *ϕ* (purple).

**Figure S13:**
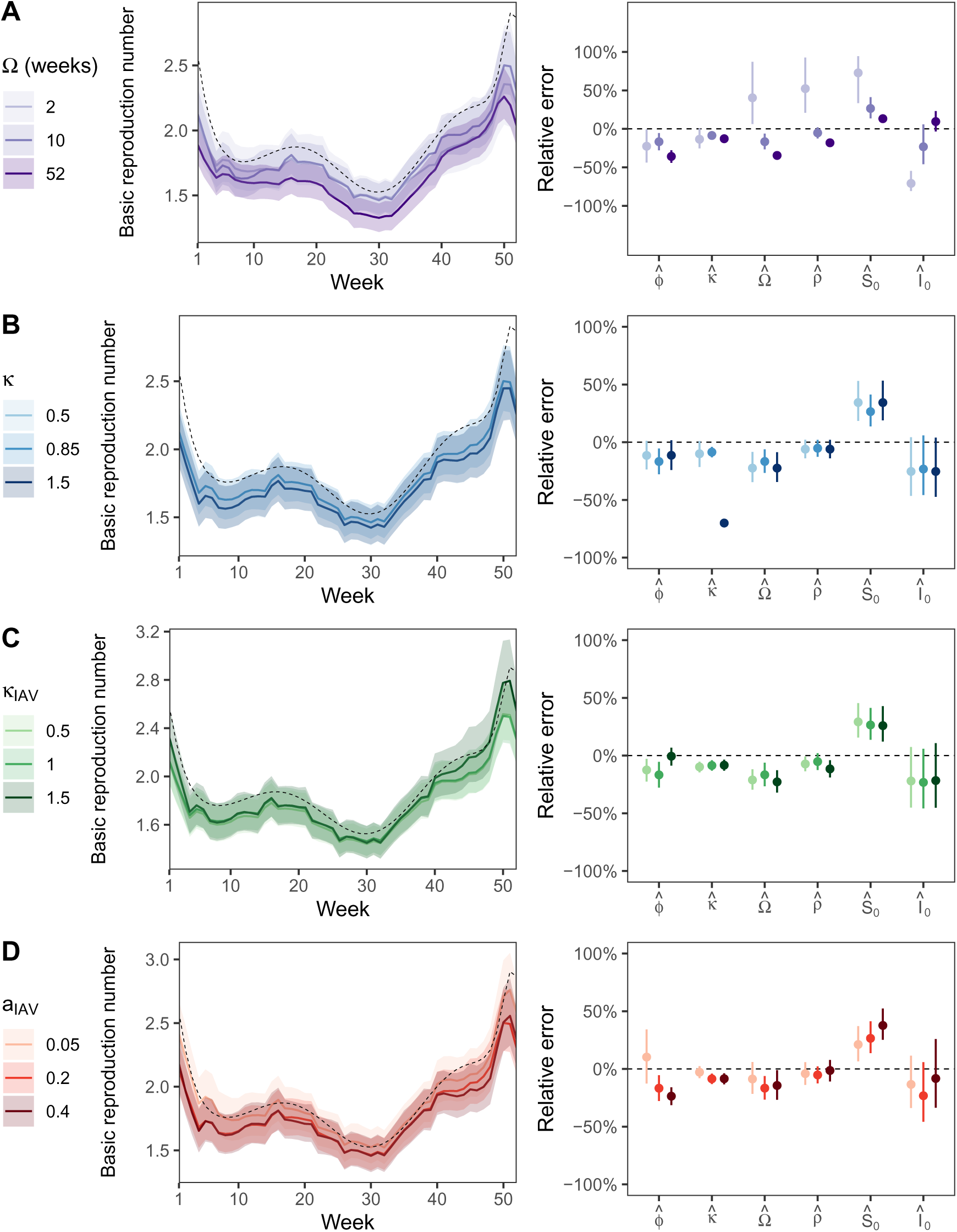
Sensitivity of estimates to chosen parameter values. Built upon the main simulation of scenario 2 (“masked interaction”), we explored the sensitivity of our estimates to some key parameter values of the viruses. We varied (A) the mean duration of immune protection to RV reinfection Ω, (B) the strength of the effect of NPIs on RV transmission κ and (C) on IAV transmission κ_IAV_ and (D) the amplitude of the seasonal forcing of IAV a_IAV_. For each parameter value, we regenerated simulated data and fitted the model with potential viral interaction to the whole dataset (i.e., including the (post-)pandemic period). Parameter estimates remained robust across the explored range of values, except maybe parameters Ω, ρ and S_0_ that were all overestimated when Ω was very short (fixed 2 weeks), and parameters κ that were quite underestimated when its value was high (fixed to 1.5). Solid lines and points denote median values, while envelopes and segments denote 95% CrIs; true values are indicated by dashed lines.

**Figure S 14:**
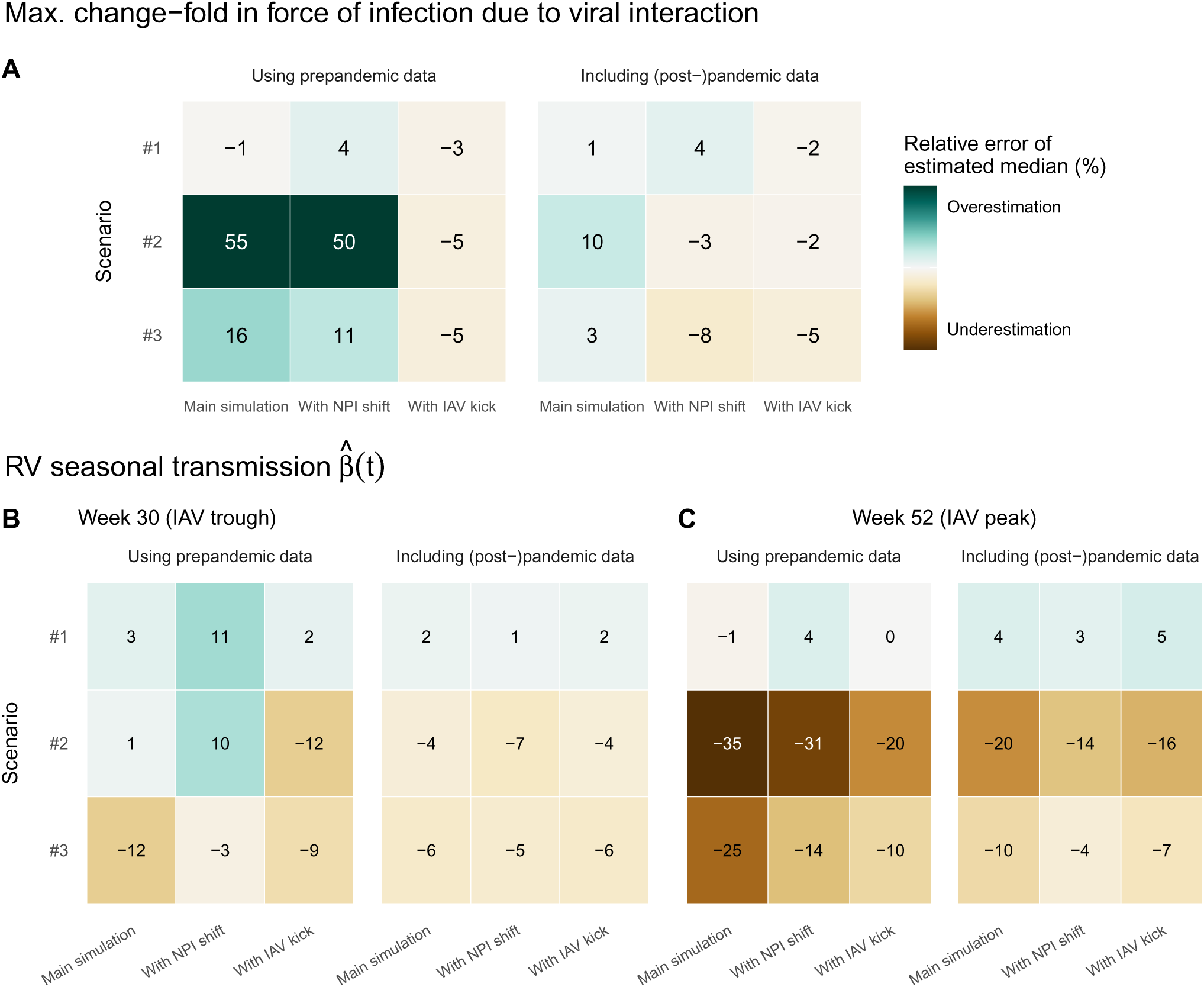
Relative errors in estimates of viral interaction and seasonal forcing using simulated data. We compared the relative errors in the estimated median of (A) the maximum change-fold in RV force of infection due to viral interaction of IAV 1 + *ϕ̂* and (B) RV seasonal transmission *β̂*(t) at the 30^th^ week (when IAV circulation is minimal) and 52^th^ week (when IAV circulation is maximal). For each case, we present results using pre-pandemic data and including (post-)pandemic data. In each heatmap, rows represent the three scenarios and columns represent the corresponding main simulation and two variations: one where the timing of NPI occurs 6-month earlier (*With NPI shift*) and one where we introduce a one-off exogenous perturbation in IAV dynamics, moving 35% of the susceptibles to the recovered compartment during the pre-pandemic period (*With IAV kick*).

**Figure S15:**
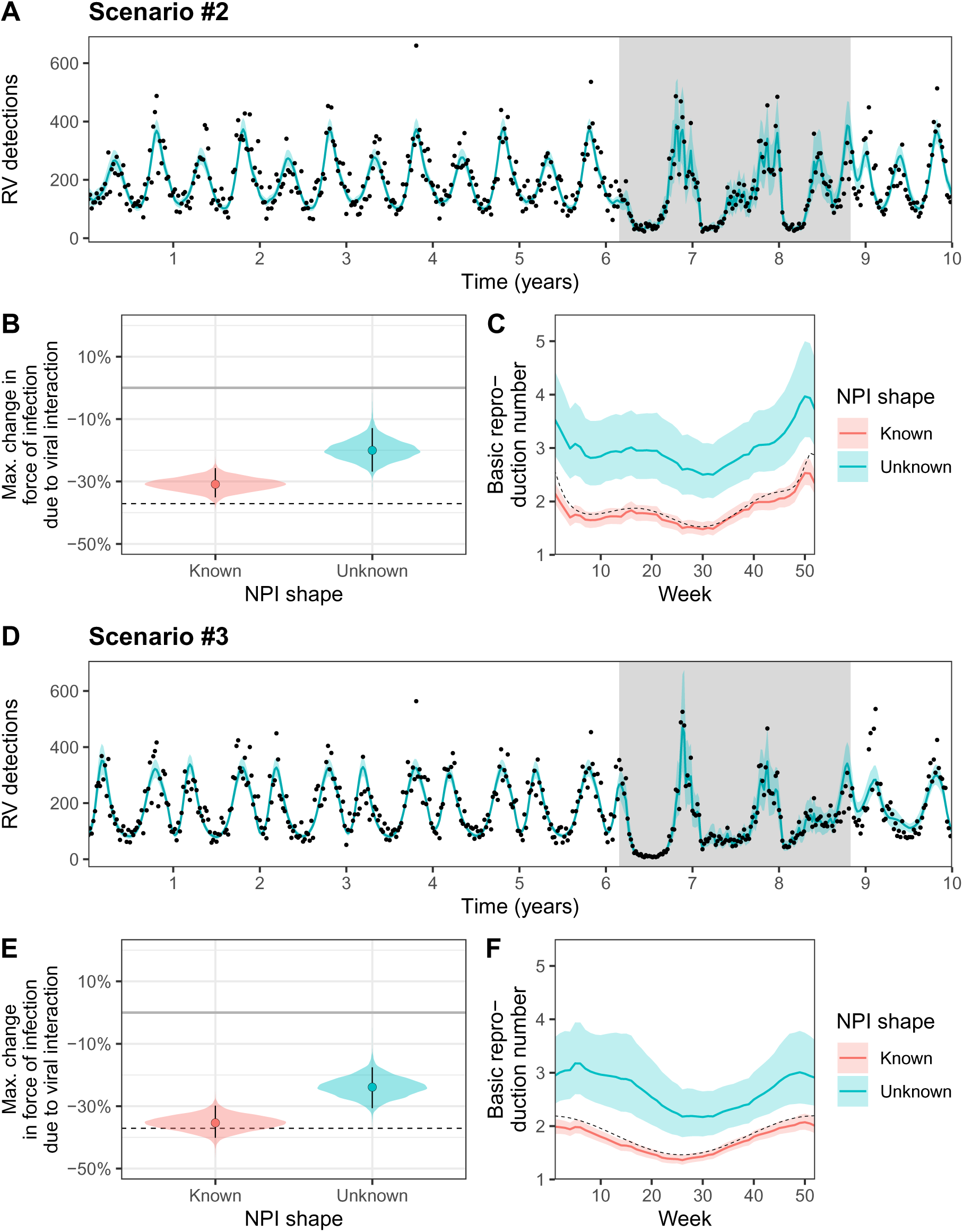
Unknown shape of NPI perturbations can hamper the inference of viral interaction and seasonal forcing. Here, we use simulated data from the main simulations of scenarios 2 (A-C) and 3 (D-F); in both scenarios IAV negatively impacts RV (*ϕ* = 0). We contrast results obtained when constraining the shape of NPIs (in red, using Google mobility data) or not (in blue). In the latter, we estimated the NPI effect of each week of the pandemic period (gray background in A and C) using a uniform prior between 0 and 1. (A,D) Median (lines) and 95% CrIs (envelopes) of the fitted values when the shape of NPIs is unknown and estimated. (B,E) Posterior distributions, median (point) and 95% CrIs (segments) of the maximum interaction effect due to IAV and (C,F) Median (lines) and 95% CrIs (envelopes) of RV basic reproduction (proportional to seasonal forcing). True values are indicated by dashed lines.

**Figure S 16:**
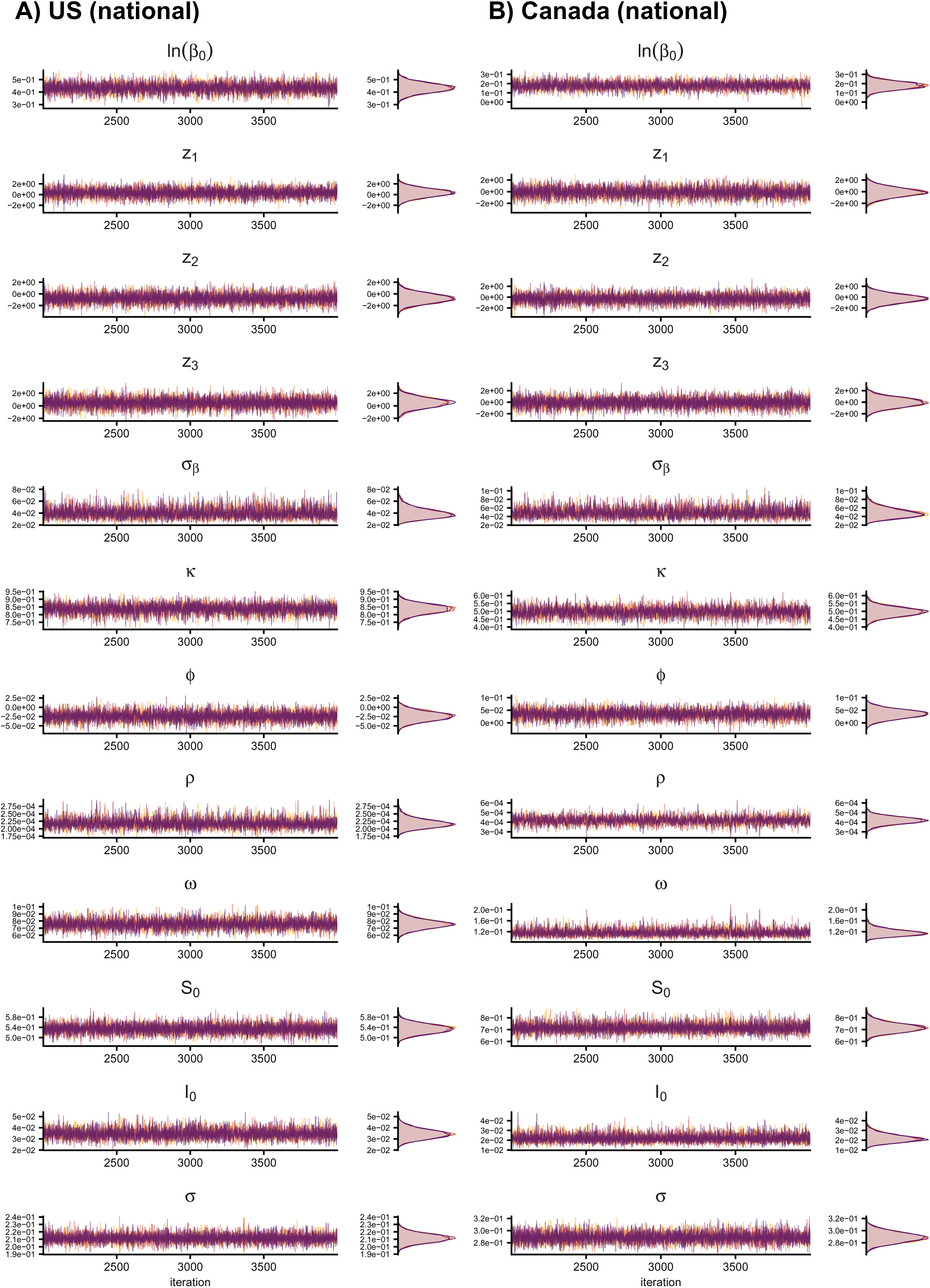
Examples of MCMC chains and posterior distributions. Examples from the model fitted to national-level time series from (A) US and (B) Canada. The first 2,000 iterations (burn-in periods) are not represented.

**Figure S 17:**
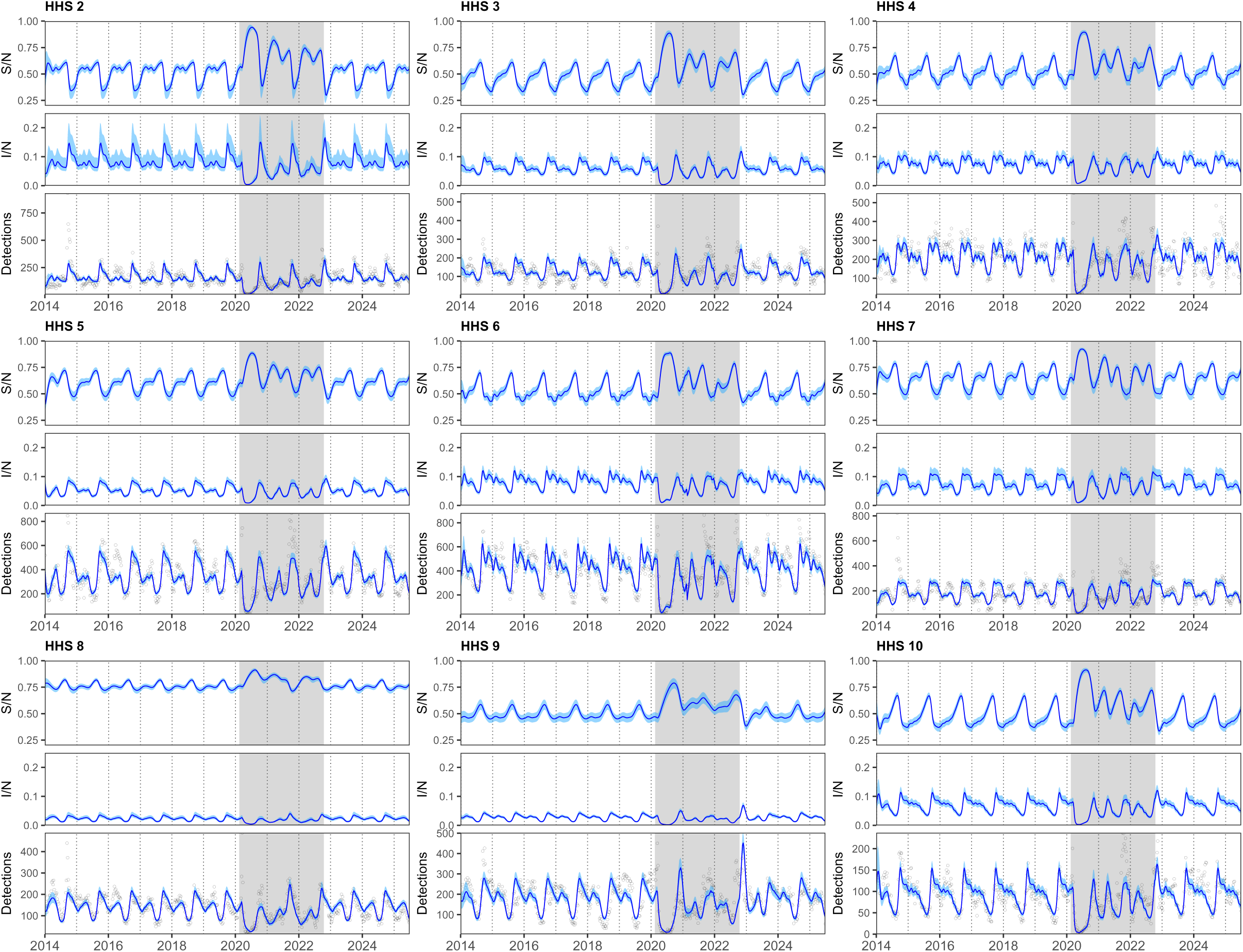
Fitted model results in the US at the regional level. For HHS regions 2-10, we independently fitted model (3)-(4) with seasonal transmission (8) (see **Materials and Methods §4.2-4.3**) to rescaled RV/EV detections (black points); we only show results obtained with the model with no viral interaction (*ϕ* = 0). We plot posterior median values (lines) and 95% CrIs (envelopes) for (top) the proportion susceptible S/N, (middle) the proportion infected I/N and (bottom) the number of detections. Grey backgrounds indicate the pandemic period.

**Figure S18:**
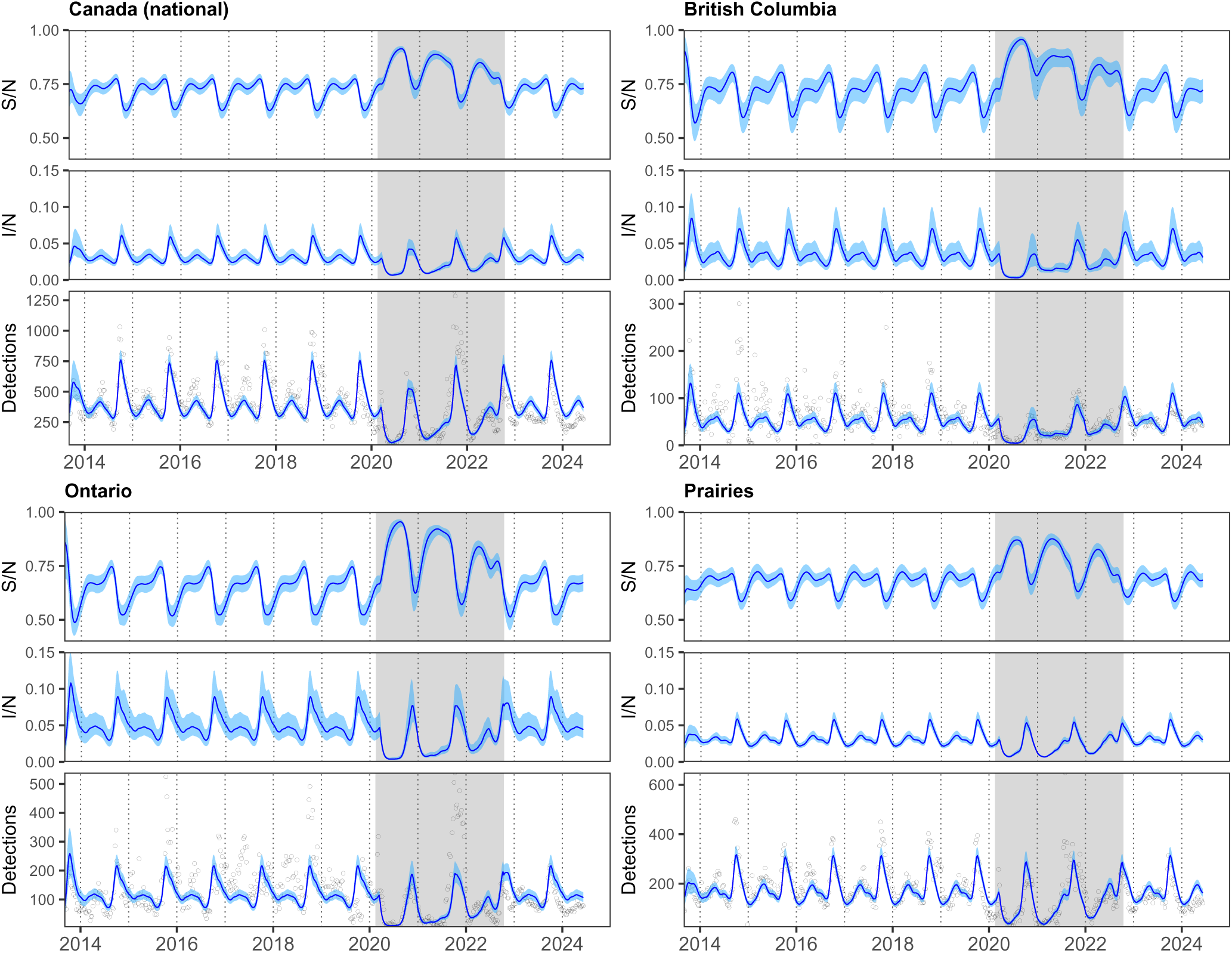
Fitted model results in Canada at the national and provincial/regional level. For each location, we independently fitted model (3)-(4) with seasonal transmission (8) (see **Materials and Methods §4.2-4.3**) to rescaled RV/EV detections (black points); we only show results obtained with the model with no viral interaction (*ϕ* = 0). We plot posterior median values (lines) and 95% CrIs (envelopes) for (top) the proportion susceptible S/N, (middle) the proportion infected I/N and (bottom) the number of detections. Grey backgrounds indicate the pandemic period.

**Figure S19:**
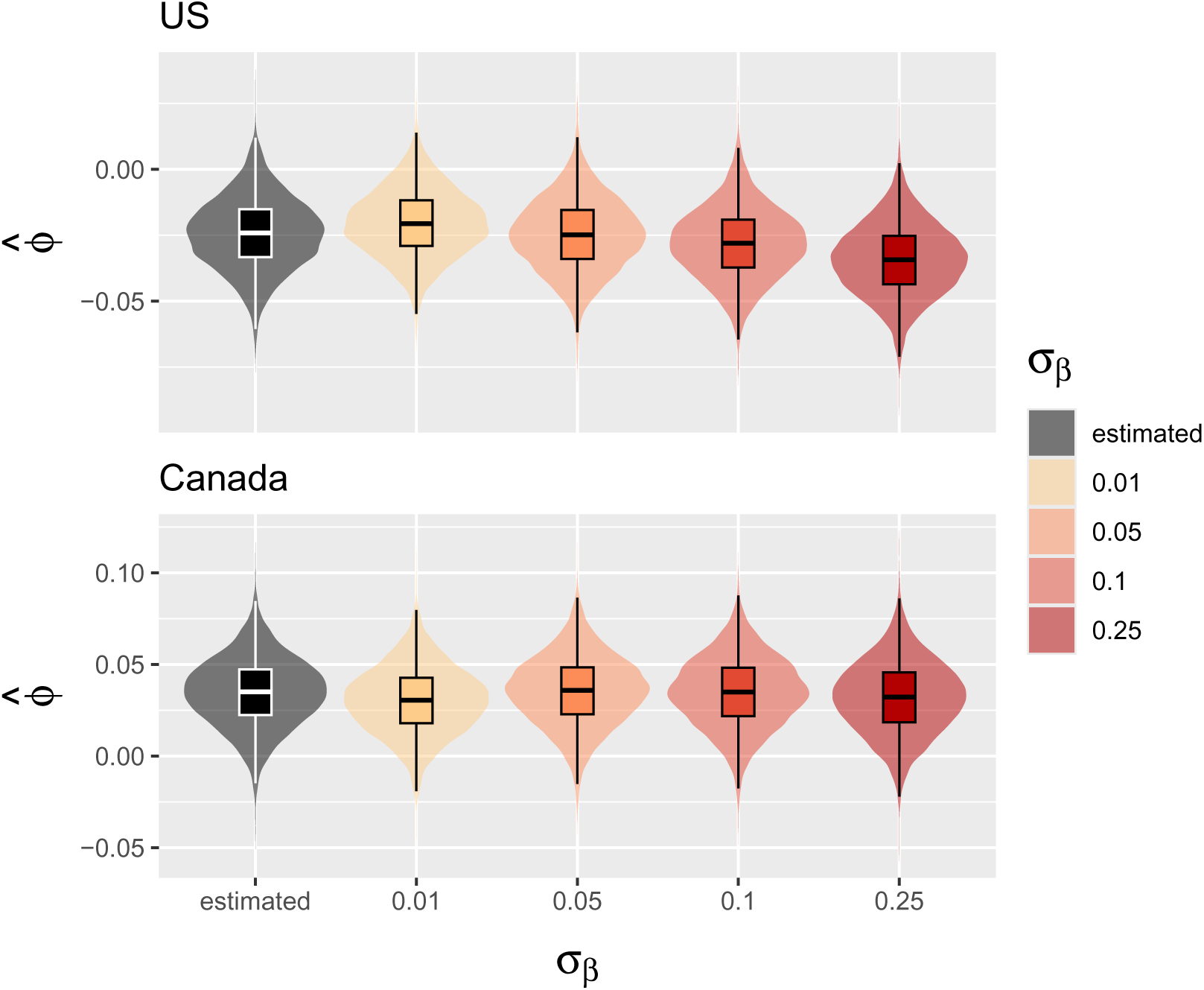
Sensitivity analysis to *σ_β_*. We assessed the robustness of the inferred posterior distribution of the viral interaction parameter *ϕ* to the prior constraint on seasonal transmission variability. To do so, we refitted the model to national-level data while fixing *σ*_*β*_ – the scale parameter governing the magnitude of weekly increments in the random walk for the seasonal transmission rate – at increasing values from 0.01 to 0.25.

**Figure S20:**
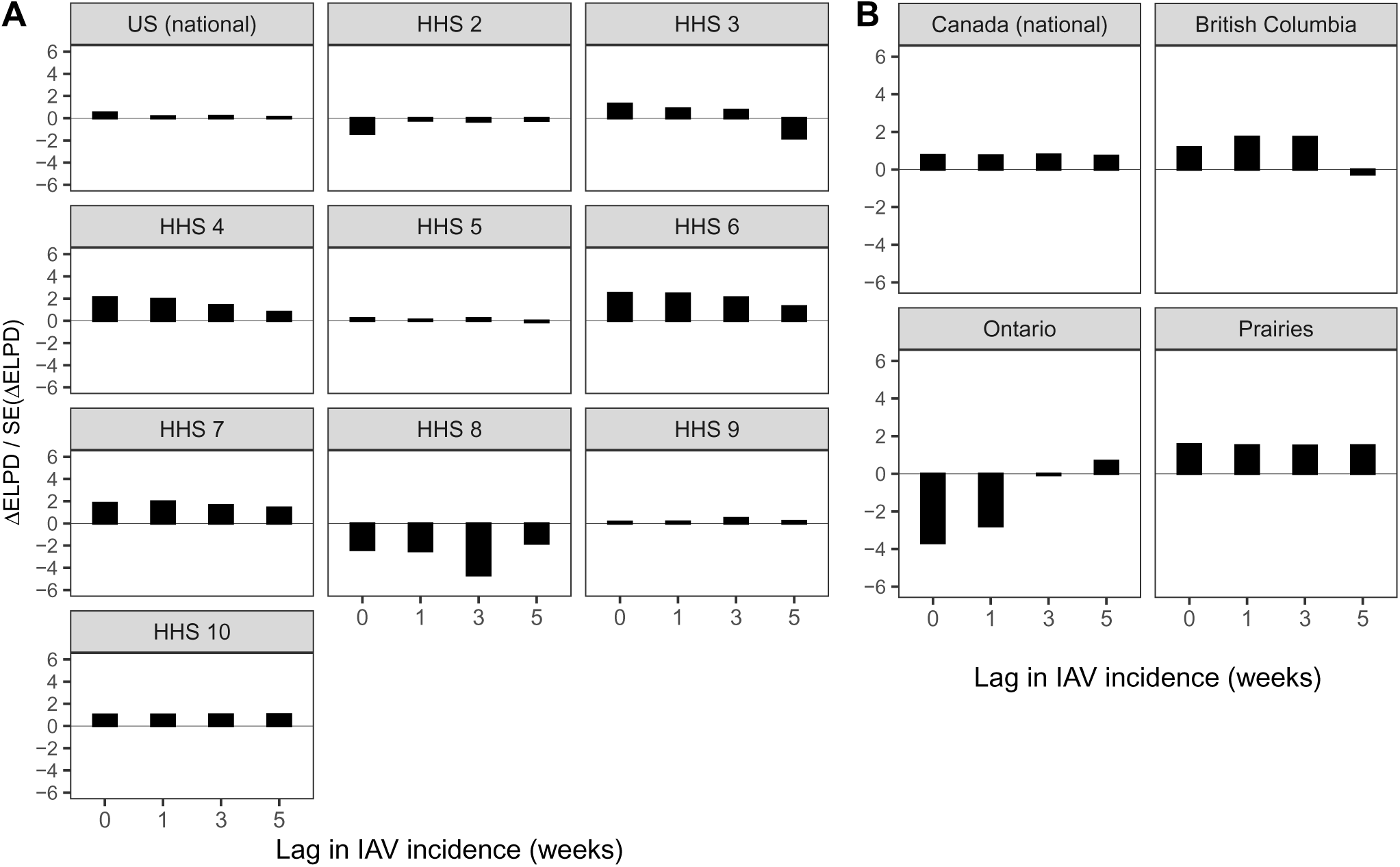
Model comparisons. We computed approximate leave-one-out cross-validation using Pareto smoothed importance sampling, as implemented in the R package loo [5] (we evaluated exact log-likelihood for the very few observations with Pareto k-values > 0.7). Fitted models allowing for potential viral interaction – using current IAV incidence (lag=0) or lagged sum of IAV incidence of 1, 3 or 5 weeks – were compared against a null model with no interaction (*ϕ* = 0). Comparisons are based on differences in expected log-predictive density ΔELPD relative to the null model, normalized by their standard error SE(ΔELPD). Absolute values below 2–4 are generally considered insufficient to provide strong evidence favoring the model with higher ELPD, in which case the more parsimonious model is preferred.

**Figure S 21:**
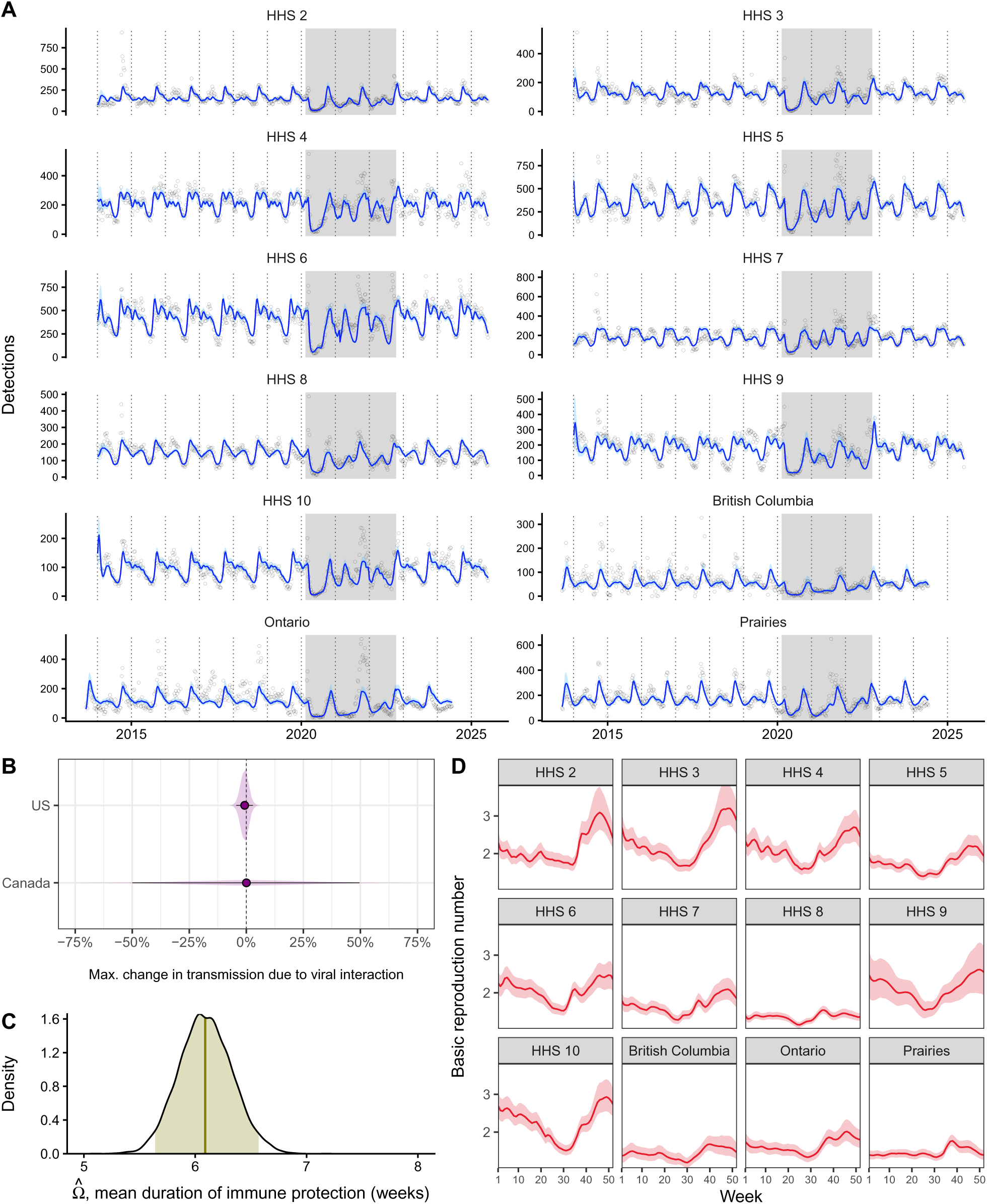
Joint model fit across all locations. Using IAV current incidence, we also fitted the model simultaneously to all regions/provinces, constraining the rate of immune waning *ω* to be shared across all locations and the viral interaction parameter *ϕ* across locations within a given country (US vs. Canada). See more details in **supporting information §S3**. (A) Model fit to RV/EV detections (black points). (B) Posterior distributions (with median values (points) and 95% CrIs (segments)) of the maximum change in RV transmission due to IAV interaction for US and Canadians locations. (C) Posterior distribution of the mean duration of immune protection Ω (same for all locations). (D) Estimated profiles of RV basic reproduction number _0_. Lines denote median values and envelopes denote 95% CrIs. Grey backgrounds indicate the pandemic period.

**Figure S22:**
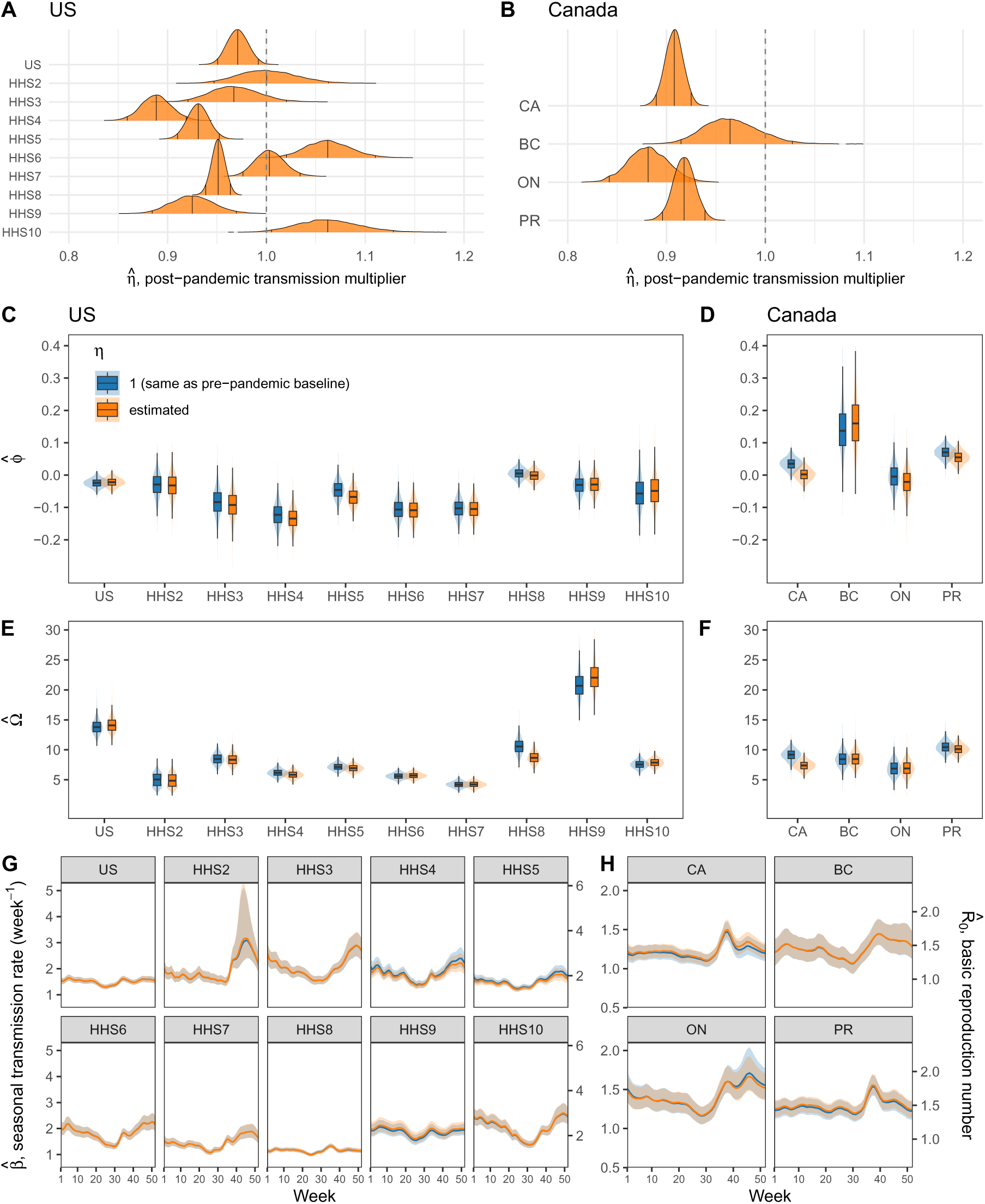
Sensitivity to post-pandemic behavioral changes. Google COVID-19 mobility data were no longer reported after October 15, 2022. In the main analysis, we assumed *c* = 0 (i.e., contacts are the same as in the pre-pandemic period) after this date. However, as NPIs were relaxed, new behavioral norms may have stabilized at non-baseline levels. We assessed the robustness of our results to this assumption by estimating a (constant) post-pandemic transmission multiplier, *η*, such that the seasonal transmission rate becomes *η β*(*t*) after October 15, 2022 (in the main analysis, *η* = 1), with prior *η* ∼ Log-*N*(0, 0.4). Estimated values were close to 1 (i.e., similar to pre-pandemic baseline) and had little impact on the estimates of the other parameters. (A-B) Posterior distributions of *η*; vertical black lines represent 2.5%, 50% and 97.5% quantiles, respectively. (C-H) Comparison of the posterior distributions of (C,D) viral interaction parameter *ϕ*, (E,F) mean duration of immune protection Ω (expressed in weeks) and (G,H) of median (lines) and 95% CrIs (envelopes) of seasonal transmission profiles. CA=Canada (national), BC=British Columbia, ON=Ontario, PR=Prairies.

**Figure S23:**
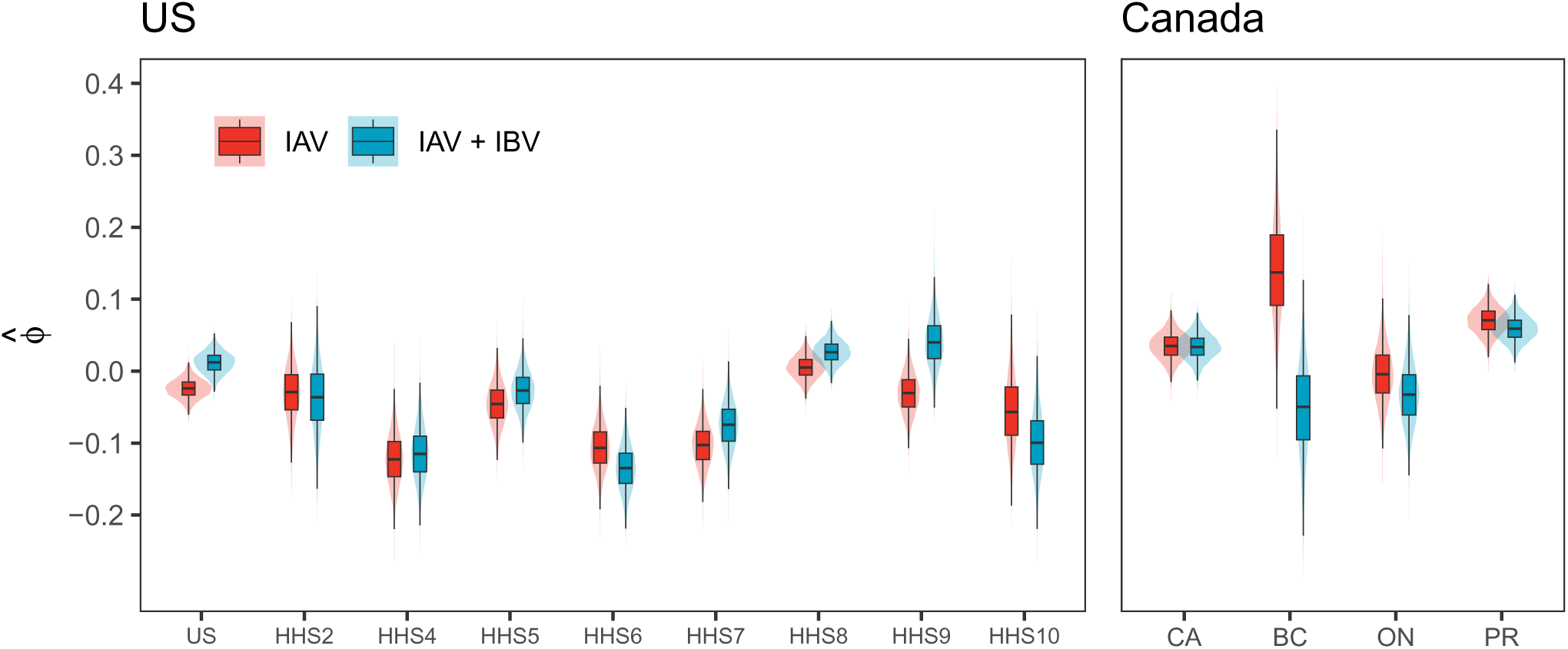
The effect of including IBV on viral interaction estimates in the US and Canada. Posterior distributions of the viral interaction parameter *ϕ* estimated using IAV current incidence (same as in the main text, in red) and combined IV current incidence (IAV+IBV, in blue). CA=Canada (national), BC=British Columbia, ON=Ontario, PR=Prairies.

**Figure S24:**
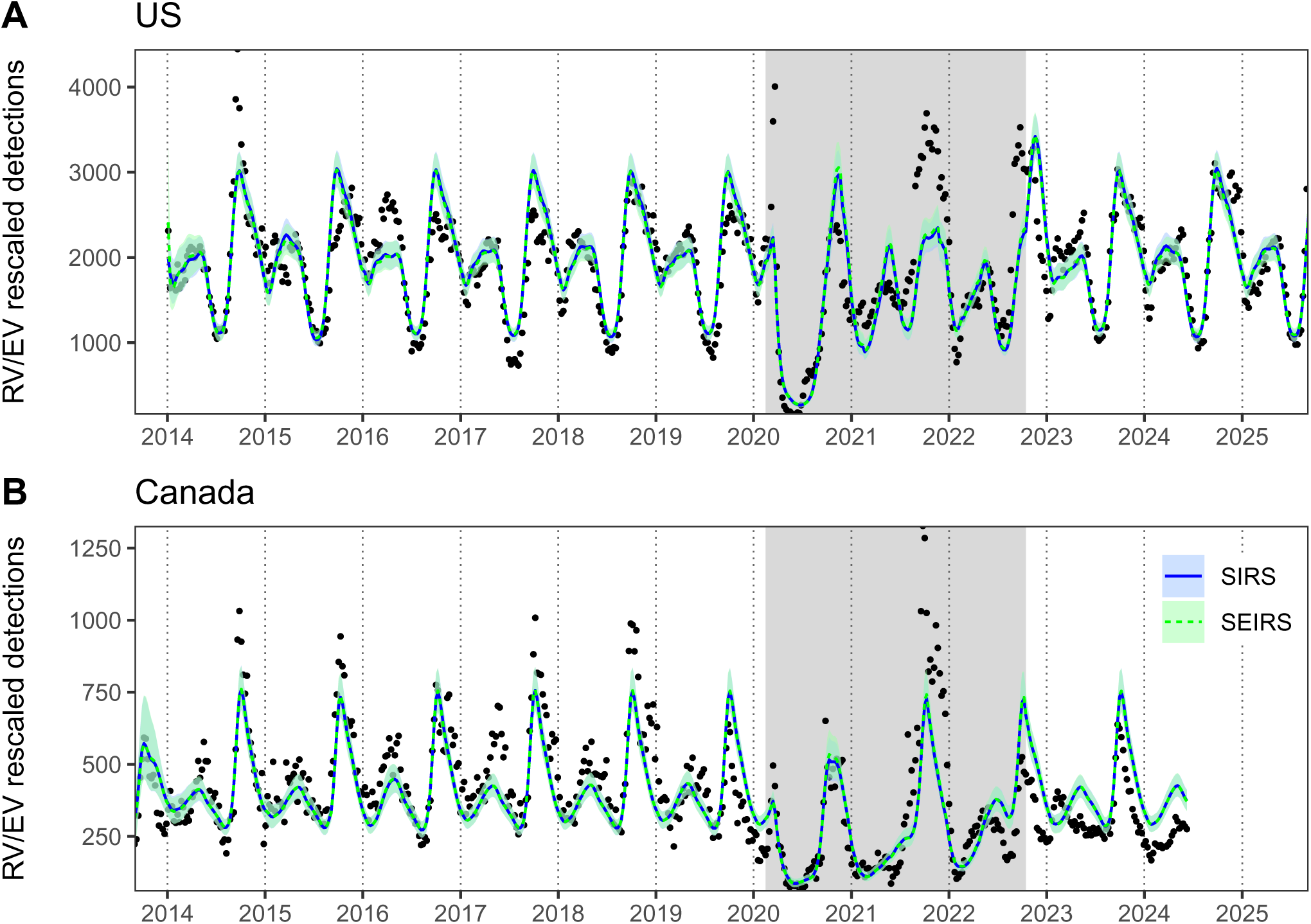
Comparison of SIRS and SEIRS model fits at the national level. Built upon the SIRS model, the SEIRS model includes an exposed (i.e., infected but not yet infectious) stage E. Because the incubation period (i.e., mean sojourn time in E) is shorter than a week, we used a smaller time step of Δt = 0.25 week to fit the discretized SEIRS model. We fixed the transition rate from E to I to 5 week^−1^ (≈2.45 days for incubation) and the recovery rate to 0.975 week^−1^ (≈8 days for recovery) [6]. We fitted both models to rescaled national RV/EV detections (points) in (A) the US and (B) Canada. Lines represent median values of posterior distributions and envelopes represent 95% CrIs; grey backgrounds indicate the pandemic period. For both models, the estimated viral interference parameter *ϕ̂* was not significantly different from 0.

**Figure S25:**
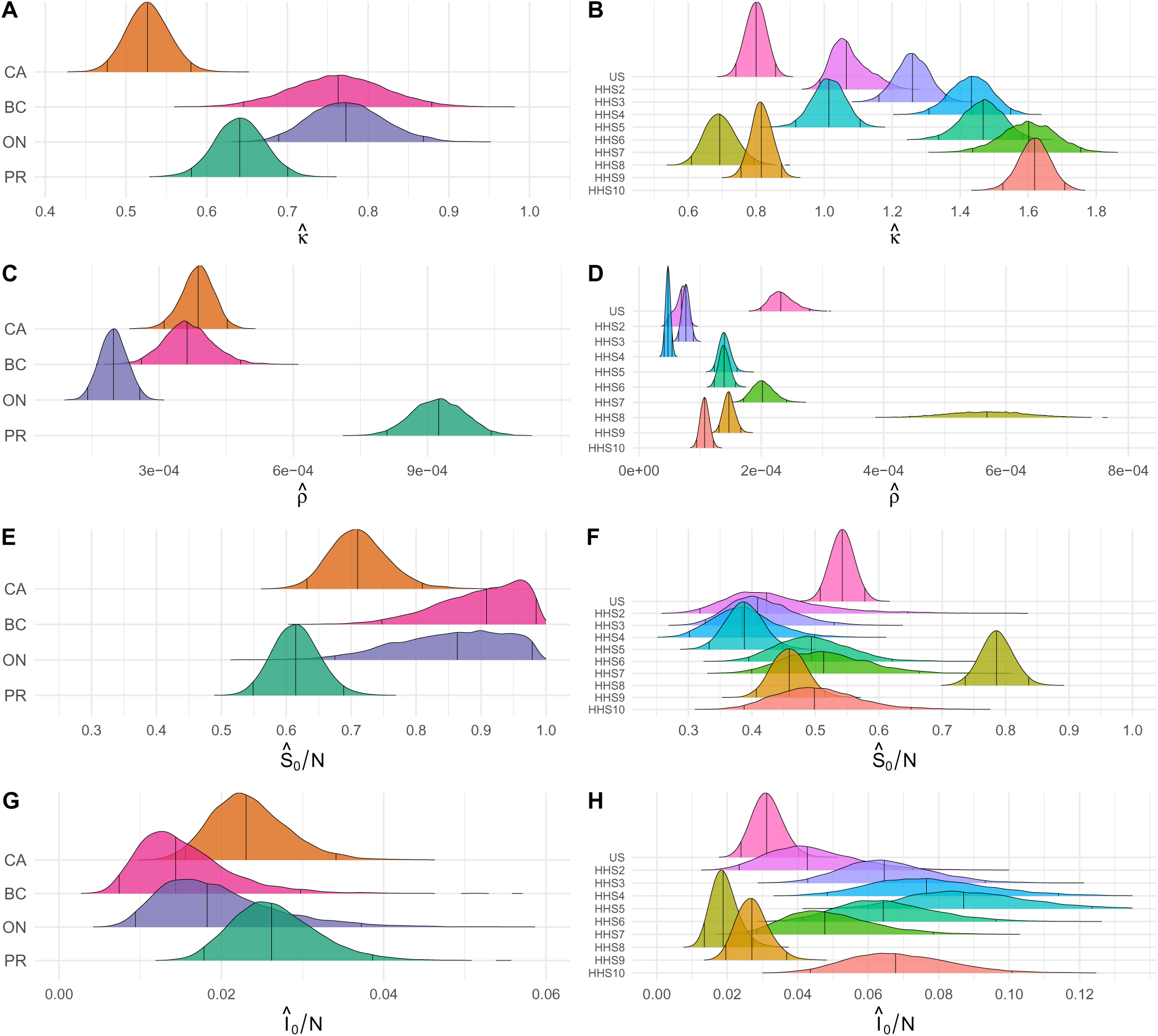
Posterior density distributions of the remaining estimated parameters. We show posterior distributions obtained with the model with no viral interaction (*ϕ* = 0) using time series from (first column) Canada and (second column) the US. Vertical black lines represent 2.5%, 50% (median) and 97.5% quantiles, respectively. See **Fig. 5** in the main text for estimates of RV weekly transmission rates and mean duration of immune protection, and see **Table 1** for notations. CA=Canada (national), BC=British Columbia, ON=Ontario, PR=Prairies.

**Figure S26:**
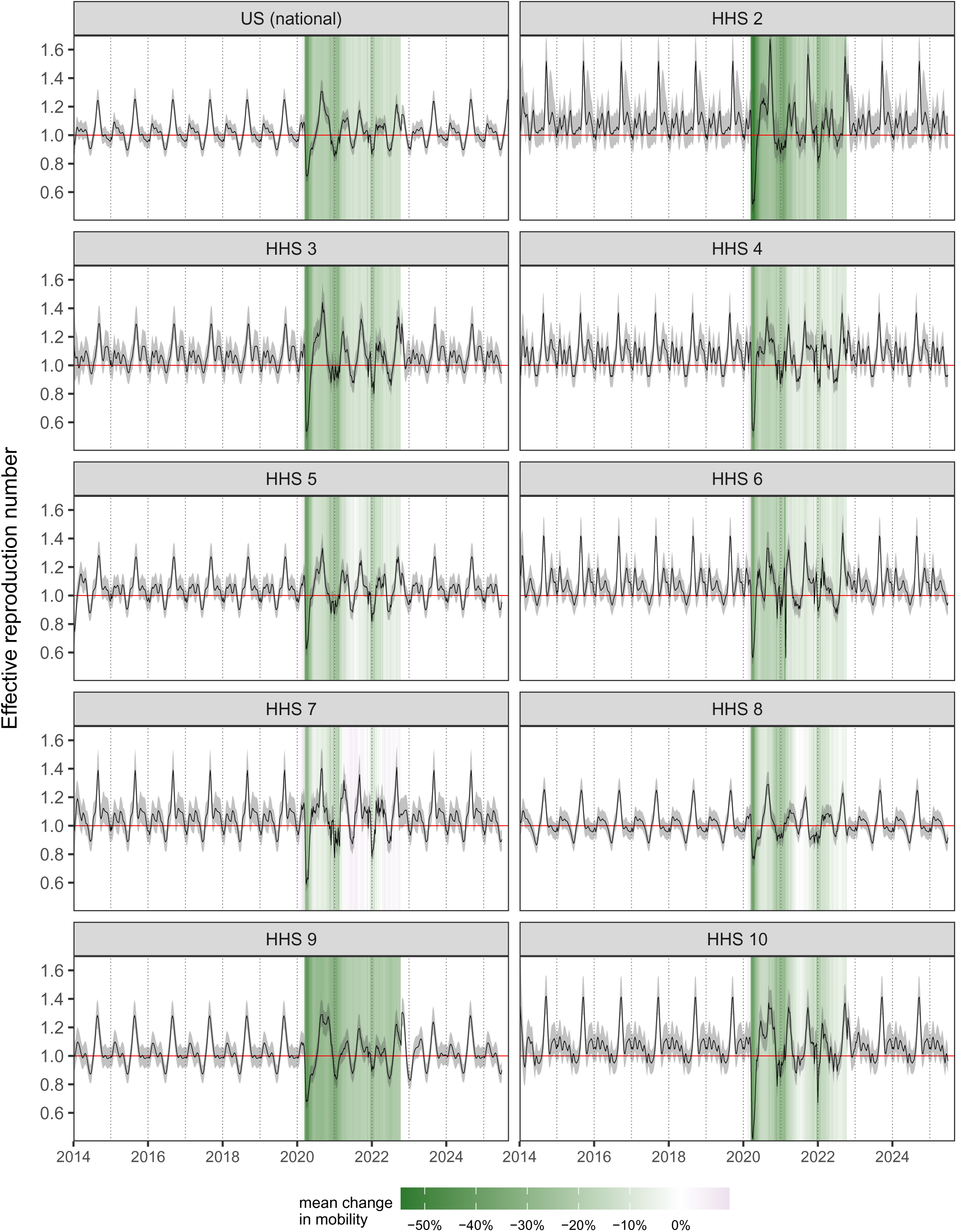
Dynamics of the estimated effective reproduction number in the US. We only show results obtained with the model with no viral interaction (*ϕ* = 0). Median (black lines) and 95% CrIs (shaded envelopes) of RV/EV effective reproduction number were computed from parameter posterior distributions. The epidemic grows as soon as > 1 (horizontal red line). Mean change in mobility (colored background) during the COVID-19 pandemic were computed from Google COVID-19 Community Mobility Reports.

**Figure S27:**
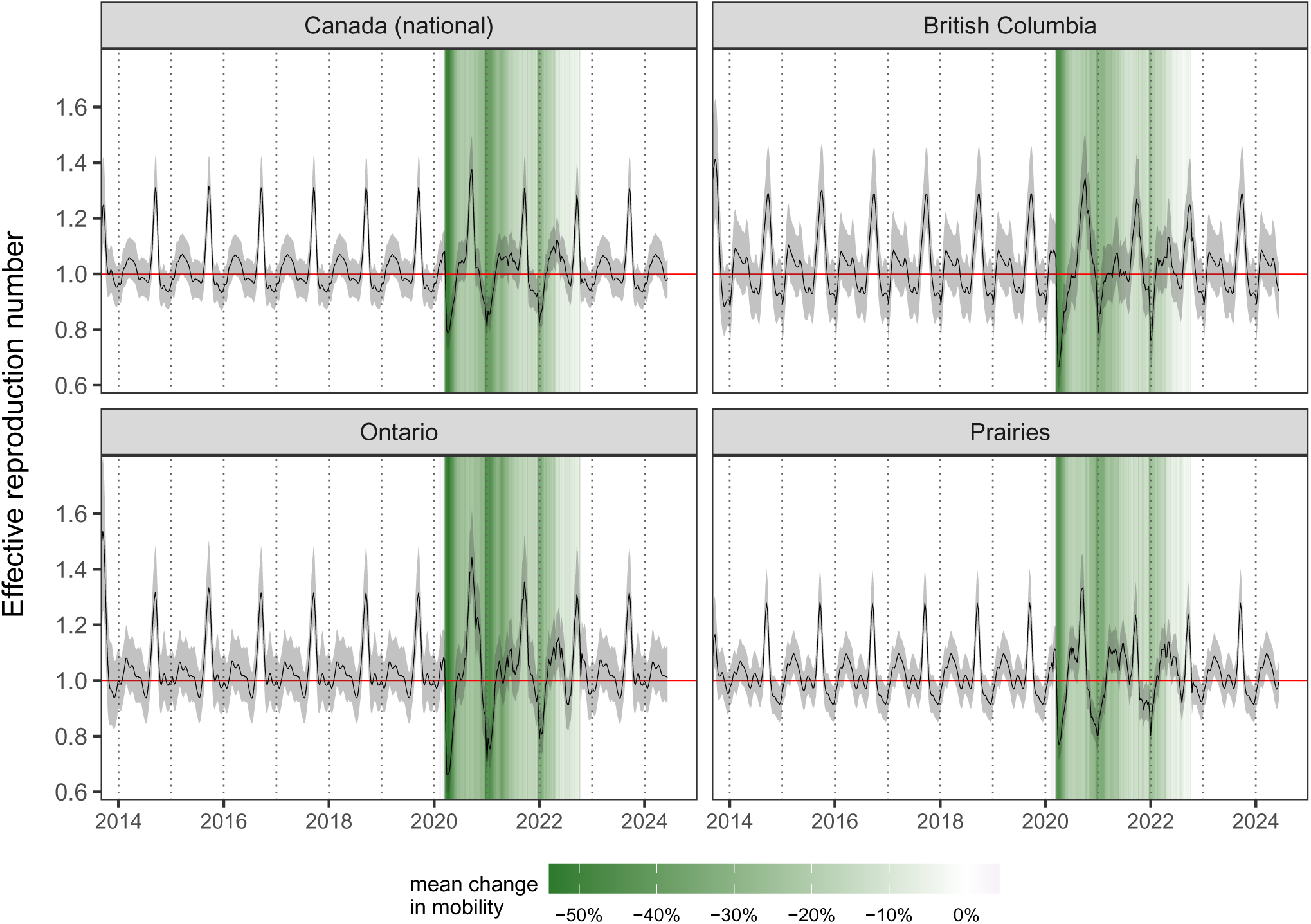
Dynamics of the estimated effective reproduction number in Canada. We only show results obtained with the model with no viral interaction (*ϕ* = 0). Median (black lines) and 95% CrIs (shaded envelopes) of RV/EV effective reproduction number were computed from parameter posterior distributions. The epidemic grows as soon as > 1 (horizontal red line). Mean change in mobility (colored background) during the COVID-19 pandemic were computed from Google COVID-19 Community Mobility Reports.

### S5 Supplementary tables

**Table S1:**
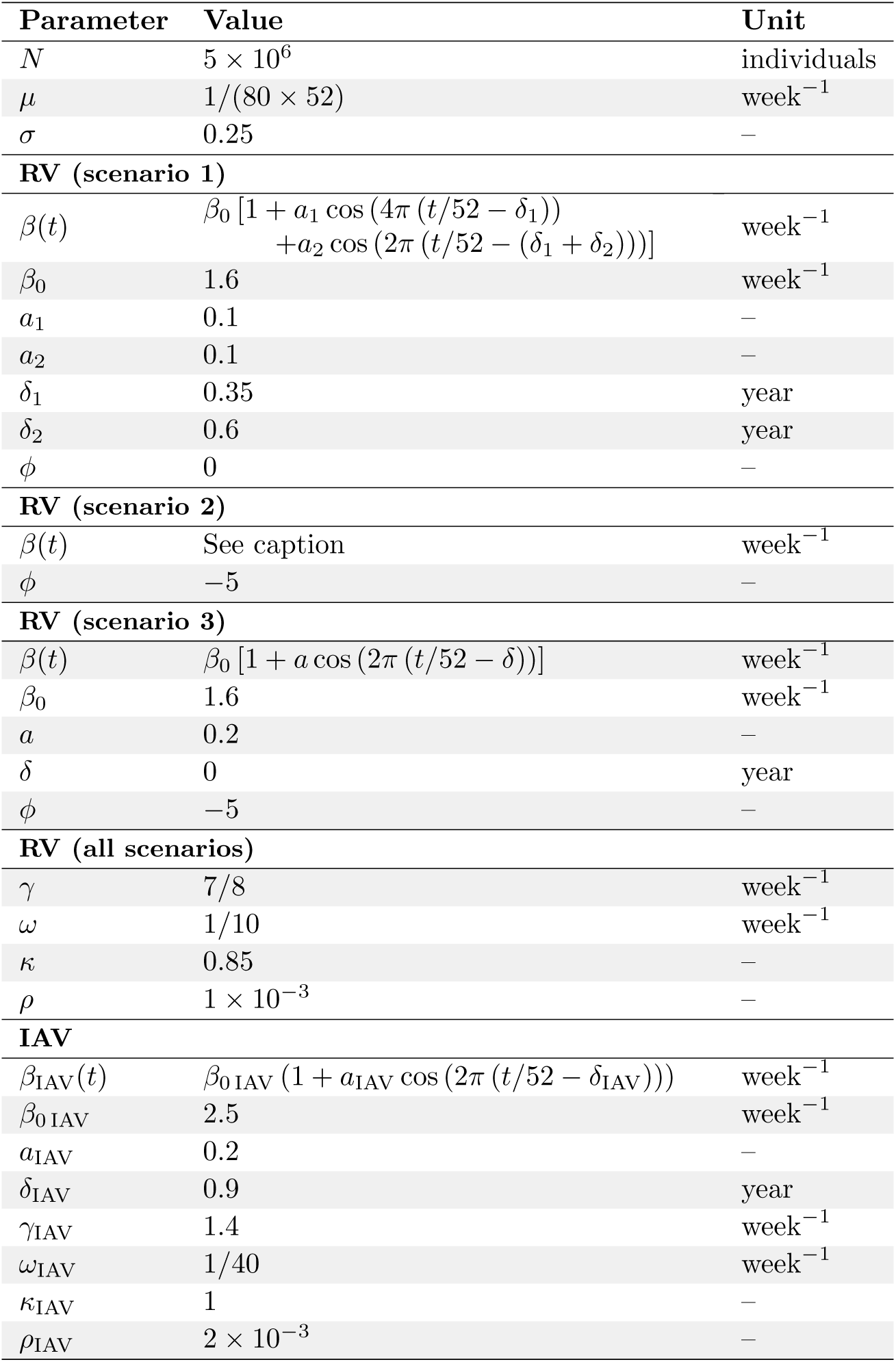
Parameters for the simulation study. Virus parameters are based on existing literature and/or to reproduce the observed epidemiological characteristics of RV and IAV. With these parameter values, IAV exhibits a single annual peak during winter, whereas RV shows two seasonal peaks in fall and spring. The basic reproduction number (*R*_0_ = *β*/(*γ* + *µ*)) ranges between 1.53 and 2.17 for RV, and between 1.33 and 2.0 for IAV, which is consistent with [6–8]. The mean duration of infectiousness is set to 1/*γ*_IAV_ =5 days for IAV [6, 9], and 1/*γ* =8 days for RV [6]. The mean duration of IAV immunity is set to 1/*ω*_IAV_ =40 weeks, reflecting temporary protection over approximately only one influenza season due to antigenic drift. For the transmission rate of RV in scenario 2, we substitute 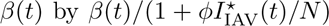 using values from scenario 1 (except for *ϕ*) and where * denotes the endemic attractor, so that the force of infection of RV mimics the one in 1 in the absence of perturbation.

**Table S2:**
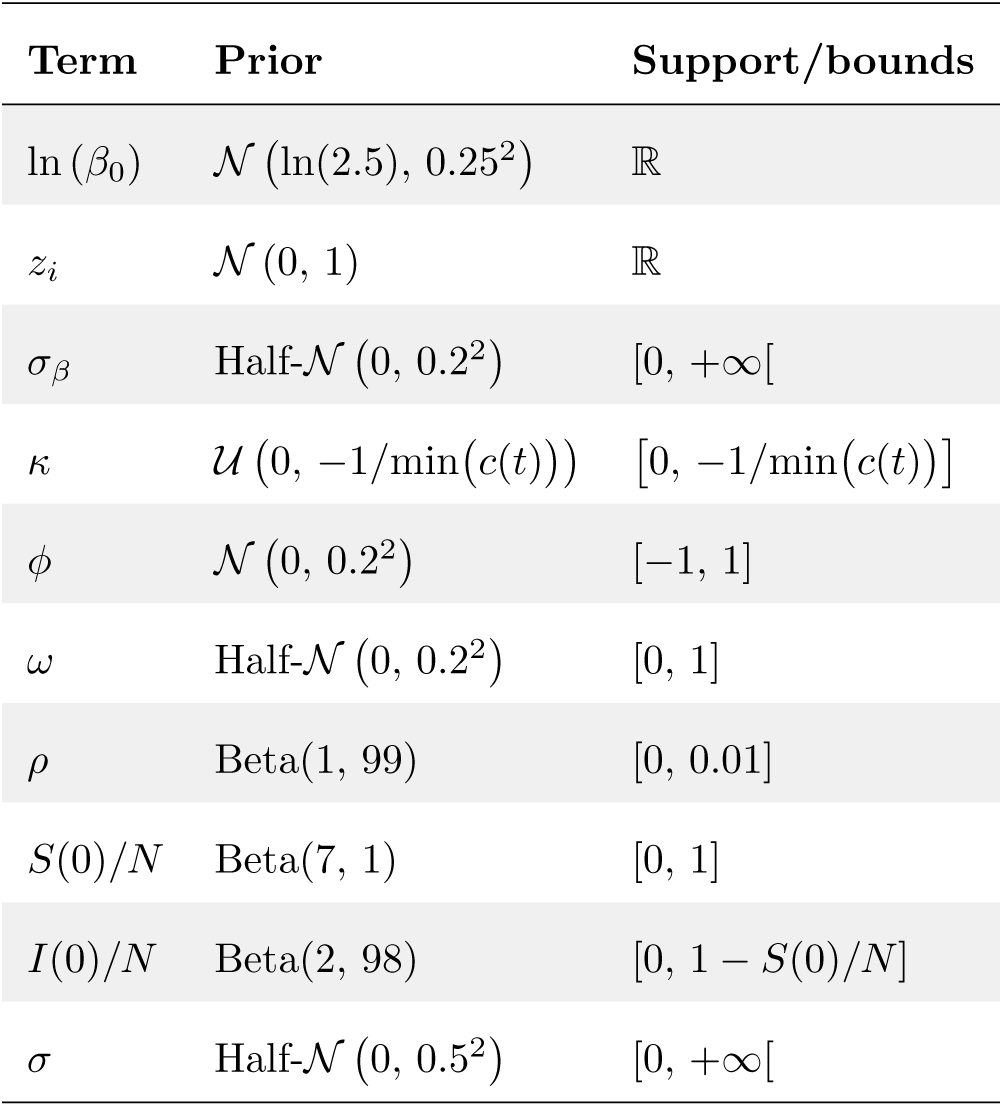
Priors. (Half-)*N µ*, *σ*^2^ refers to the (half-)normal distribution with mean *µ* and variance *σ*^2^, and *U* (a, b) refers to the uniform distribution between a and b.

**Table S3:**
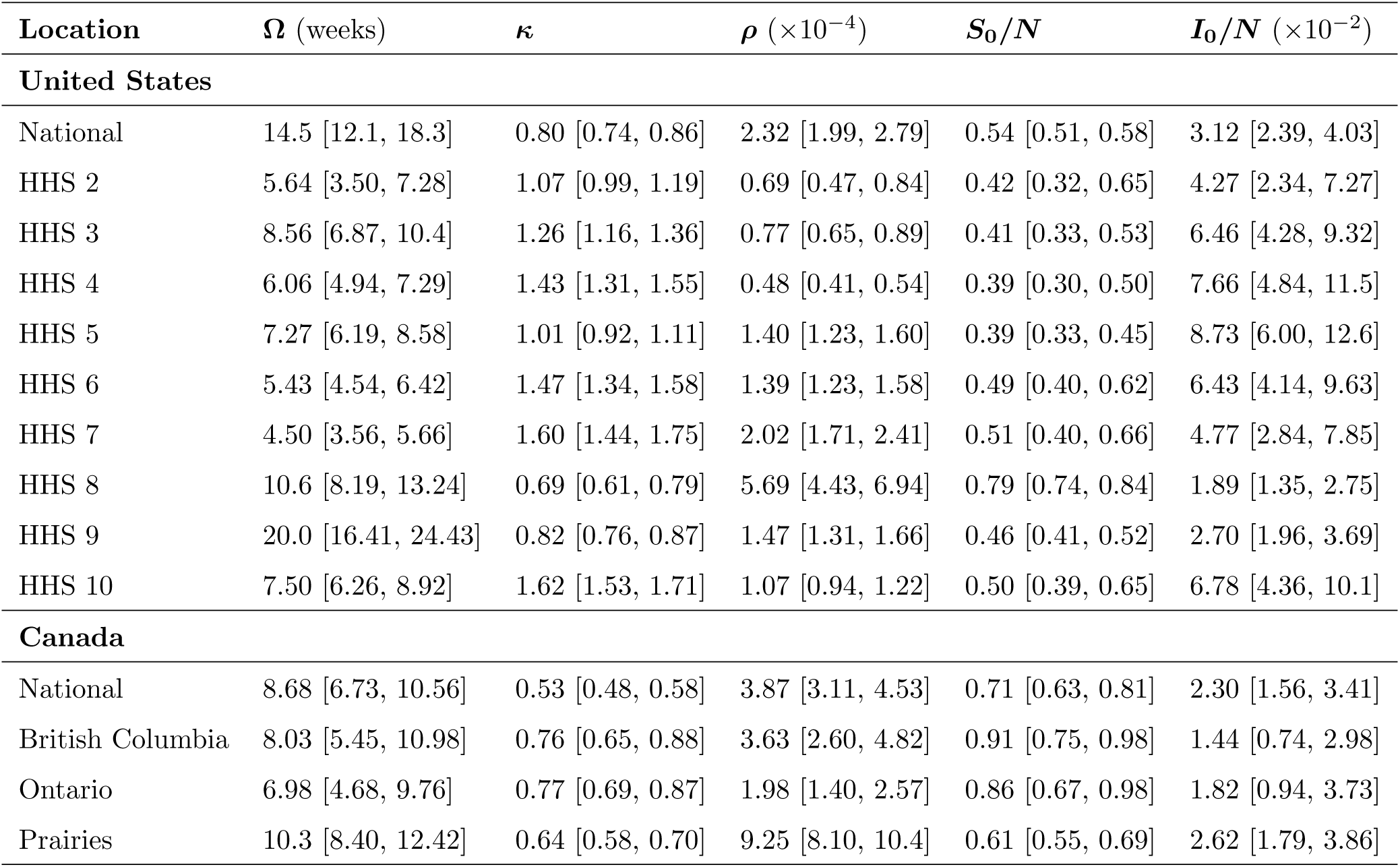
Parameter estimates. Median and 95% CrIs of posterior distributions obtained with the model without viral interaction (see distributions in **Fig. S25**). We plot estimations of seasonal transmission profiles *β*(t) and mean durations of immune protection Ω in **Fig. 5**.

