## Supplementary text, figures and tables for "Leveraging perturbations to infer the population dynamics of human rhinovirus and interaction of influenza A virus"

July 29, 2026

**Fig. S1** Colored map of the studied regions.
**Fig. S2** RV and IAV time series (US).
**Fig. S3** RV and IAV time series (Canada).
**Fig. S4** RV testing patterns.
**Fig. S5** IAV testing patterns.
**Fig. S6** Cross-wavelet transform of IAV and RV detections in the US.
**Fig. S7** Cross-wavelet transform of IAV and RV detections in Canada.
**Fig. S8** Simulated dynamics.
**Fig. S9** Simulation study estimation results (scenario 1).
**Fig. S10** Simulation study estimation results (scenario 2).
**Fig. S11** Simulation study estimation results (scenario 3).
**Fig. S12** Simulation study estimation results with high observation noise.
**Fig. S13** Sensitivity of estimates to chosen parameter values.
**Fig. S14** Relative errors in estimates of viral interaction and seasonal forcing using simulated data.
**Fig. S15** Unknown shape of NPI perturbations can hamper the inference of viral interaction and seasonal forcing.
**Fig. S16** Examples of MCMC chains and posterior distributions.
**Fig. S17** Fitted model results in the US at the regional level.
**Fig. S18** Fitted model results in Canada at the national and provincial/regional level.
**Fig. S19** Sensitivity analysis to $\sigma_\beta$.
**Fig. S20** Model comparisons.
**Fig. S21** Joint model fit across all locations.
**Fig. S22** Sensitivity to post-pandemic behavioral changes.
**Fig. S23** The effect of including IBV on viral interaction estimates in the US and Canada.
**Fig. S24** Comparison of SIRS and SEIRS model fits at the national level.
**Fig. S25** Posterior density distributions of the remaining estimated parameters.
**Fig. S26** Dynamics of the estimated effective reproduction number in the US.
**Fig. S27** Dynamics of the estimated effective reproduction number in Canada.

**S5. Supplementary tables** ........ **32**

**Table S1** Parameters for the simulation study.
**Table S2** Priors.
**Table S3** Parameter estimates.

**Supplementary references** ........ **35**

### S1 Cross-wavelet transform analysis

We performed cross-wavelet transform analysis to examine the relationships in time-frequency space between rescaled RV/EV case detections ($C$) and rescaled IAV case detections ($C_{\mathrm{IAV}}$).

Let ($x_t$, $t = 1, \ldots, N$) be a time series. Its continuous wavelet transform is given by:

$$W_t^X(s) = \sqrt{\frac{\delta t}{s}} \sum_{\tau=1}^{N} x_\tau \psi_0 \left[ (\tau - t) \frac{\delta t}{s} \right],$$

with $s$, the wavelet scale, $\delta t$, the time step, and $\psi_0$, the mother wavelet (Morlet wavelet by default) [1]. The cross-wavelet transform of two time series $x_t$ and $y_t$ is then given by:

$$W_t^{XY}(s) = W_t^X(s)\, W_t^{Y*}(s),$$

where * denotes complex conjugation. Cross-wavelet power is defined as the corresponding modulus, $\left|W_t^{XY}(s)\right|$, and represents the local strength of common power between the two time series in time-frequency space, while the complex argument yields their local relative phase [1].

In practice, we use the function `xwt` from the R package `biwavelet` [2]. Time series were first square-root transformed to stabilize variance and standardized to ensure comparability between time series prior to cross-wavelet analysis, that is:

$$x_t = \frac{\sqrt{C_{IAV}} - \overline{\sqrt{C_{IAV}}}}{\mathrm{SD}\left(\sqrt{C_{IAV}}\right)}, \quad \text{and} \quad y_t = \frac{\sqrt{C} - \overline{\sqrt{C}}}{\mathrm{SD}\left(\sqrt{C}\right)},$$

where the overline denotes the corresponding arithmetic mean and SD, the standard deviation. The cross-wavelet power was standardized to facilitate comparison across locations. Specifically, the standardized cross-wavelet power was calculated as $\log_2\left(\left|W_t^{XY}(s)\right|\right)/(\sigma_x \sigma_y)$, where $\sigma_x$ and $\sigma_y$ are the standard deviations of the standardized input time series $x_t$ and $y_t$, respectively. Results were used to confirm and complement the initial visualization and description of the time series (i.e., the asynchronous dynamics of IAV and RV/EV, see **Fig. 1**), further motivating the subsequent analyses.

#### S2 Two-pathogen transmission model

We used the seasonally-forced SIRS model (1)-(2) to simulate RV epidemiological dynamics, using IAV incidence as an external input. In order to generate synthetic data in the simulation study, we extended this model to explicitly simulate IAV dynamics (denoted by the subscript IAV) as follows:

$$
\begin{aligned}
\dot{S}(t) &= \mu N + \omega R(t) - \left[(1+\kappa\, c(t))\left(1+\phi\,\frac{I_{\text{IAV}}(t)}{N}\right)\beta(t)\frac{I\,(t)}{N} + \mu\right] S(t) \\
\dot{I}(t) &= (1+\kappa\, c(t))\left(1+\phi\,\frac{I_{\text{IAV}}(t)}{N}\right)\beta(t)\frac{I\,(t)}{N} S(t) - (\gamma+\mu)\, I(t) \\
\dot{R}(t) &= \gamma I(t) - (\omega+\mu)\, R(t) \\
\\
\dot{S}_{\text{IAV}}(t) &= \mu N + \omega_{\text{IAV}} R_{\text{IAV}}(t) - \left[(1+\kappa_{\text{IAV}}\, c(t))\,\beta_{\text{IAV}}(t)\frac{I_{\text{IAV}}\,(t)}{N} + \mu\right] S_{\text{IAV}}(t) \\
\dot{I}_{\text{IAV}}(t) &= \beta_{\text{IAV}}(t)\frac{I_{\text{IAV}}(t)}{N} S_{\text{IAV}}(t) - (\gamma_{\text{IAV}}+\mu)\, I_{\text{IAV}}(t) \\
\dot{R}_{\text{IAV}}(t) &= \gamma_{\text{IAV}} I_{\text{IAV}}(t) - (\omega_{\text{IAV}}+\mu)\, R_{\text{IAV}}(t)
\end{aligned}
\tag{S1}
$$

where IAV transmission rate $\beta_{\text{IAV}}(t)$ is modeled as a standard sinusoidal function:

$$
\beta_{\text{IAV}}(t) = \beta_{0\,\text{IAV}}\left(1 + a_{\text{IAV}}\cos\left(2\pi\left(\frac{t}{52} - \delta_{\text{IAV}}\right)\right)\right).
$$

We listed parameter values in **Table S1**.

#### S3 Joint model fit across all locations

In the main text, we fitted the model either independently to each region/province or to data aggregated at the national level. We additionally fitted the model simultaneously to all regions/provinces, allowing to constrain selected parameters across locations. Specifically, we assumed that the rate of immune waning $\omega$ was identical across all locations and that the viral interaction parameter $\phi$ was identical across locations within a given country (US vs. Canada). In our framework, the latter implicitly assumes that reporting is sufficiently similar across locations within a country, such that relative differences in IAV incidence reflect true differences in viral circulation between locations rather than spatial variation in reporting effort. In practice, both spatial heterogeneities in IAV circulation and reporting effort are likely to exist, but we consider here this assumption to be more plausible at the country scale than across different countries. Within each country (US vs. Canada), IAV incidence was thus scaled between 0 and 1 using the global maximum of IAV incidence across all seasons and corresponding locations. The model structure is otherwise similar to that presented in the main text, with the exception of the prior of the location-specific initial conditions, $(S_{0,i},\ I_{0,i},\ R_{0,i})$, for which we specified a Dirichlet$(35, 1, 4)$ distribution.

#### S4 Supplementary figures

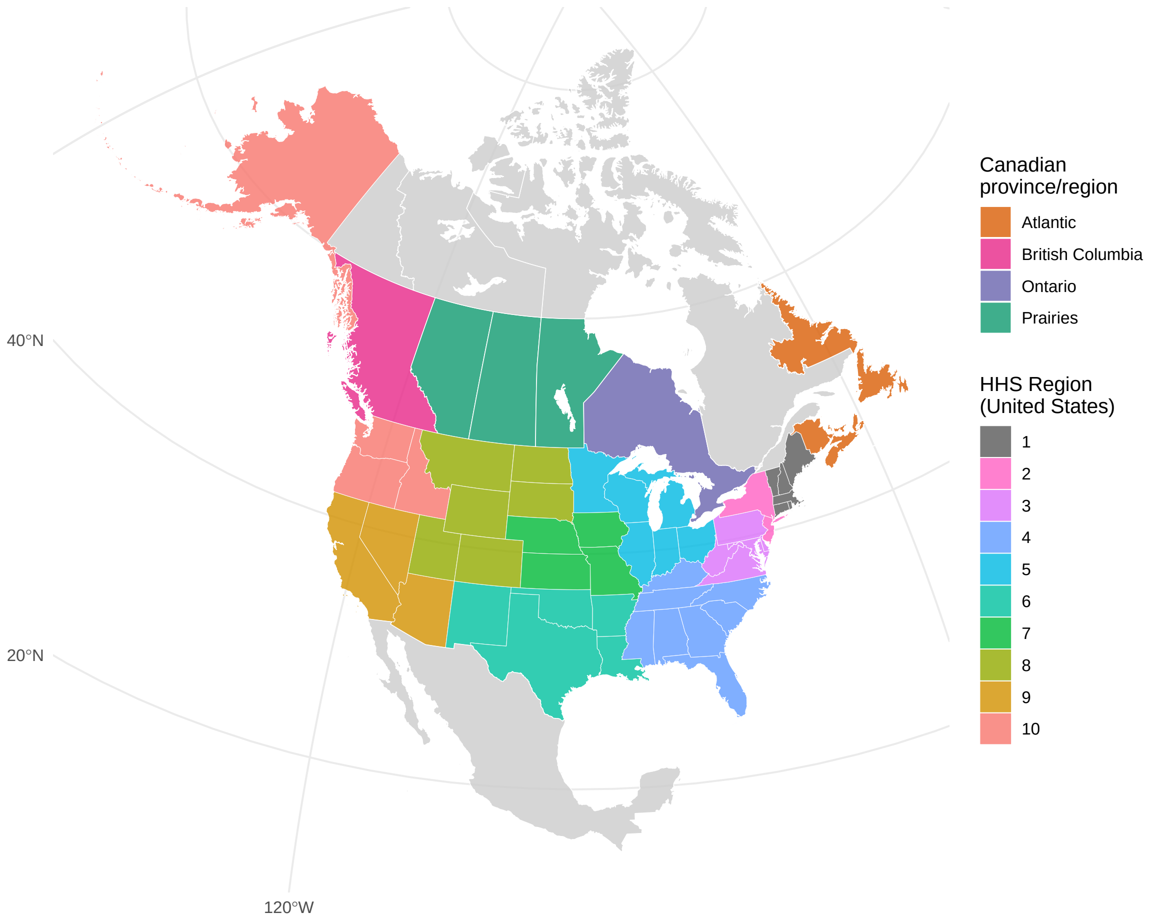

Figure S1: **Colored map of the studied regions.** We focused here on the US and Canada. At the subnational level, we considered 4 provinces/regions for Canada – Atlantic, British Columbia, Ontario and Prairies – as well as the 10 Health and Human Services (HHS) regions for the US. Not displayed: HHS region 2 also includes Puerto Rico and the Virgin Islands; HHS region 9 also includes Hawaii, American Samoa, the Commonwealth of the Northern Mariana Islands, the Federated States of Micronesia, Guam, the Marshall Islands and the Republic of Palau. Base layer of the map were obtained from Natural Earth (https://www.naturalearthdata.com/) via the `rnaturalearth` R package [3].

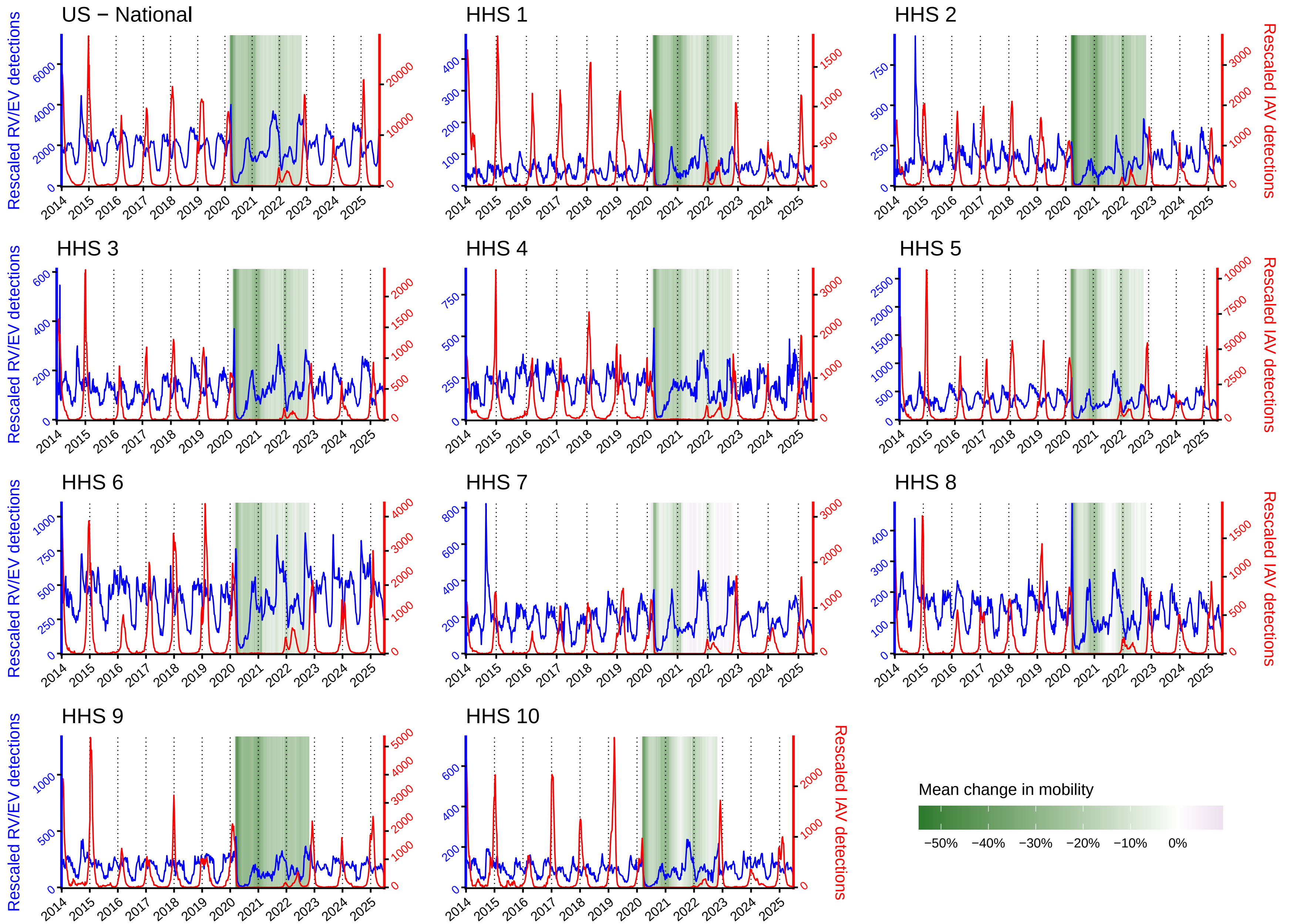

Figure S2: **RV and IAV time series (US).** RV/EV (blue) and IAV (red) detections in the US (national and 10 HHS regions). Data were rescaled to account for changes in testing patterns (see **Fig. S4** and **S5**). We plot time series from the Centers for Disease Control and Prevention (CDC), as reported through the NREVSS. Mean change in mobility (colored background) during the COVID-19 pandemic were computed from [4].

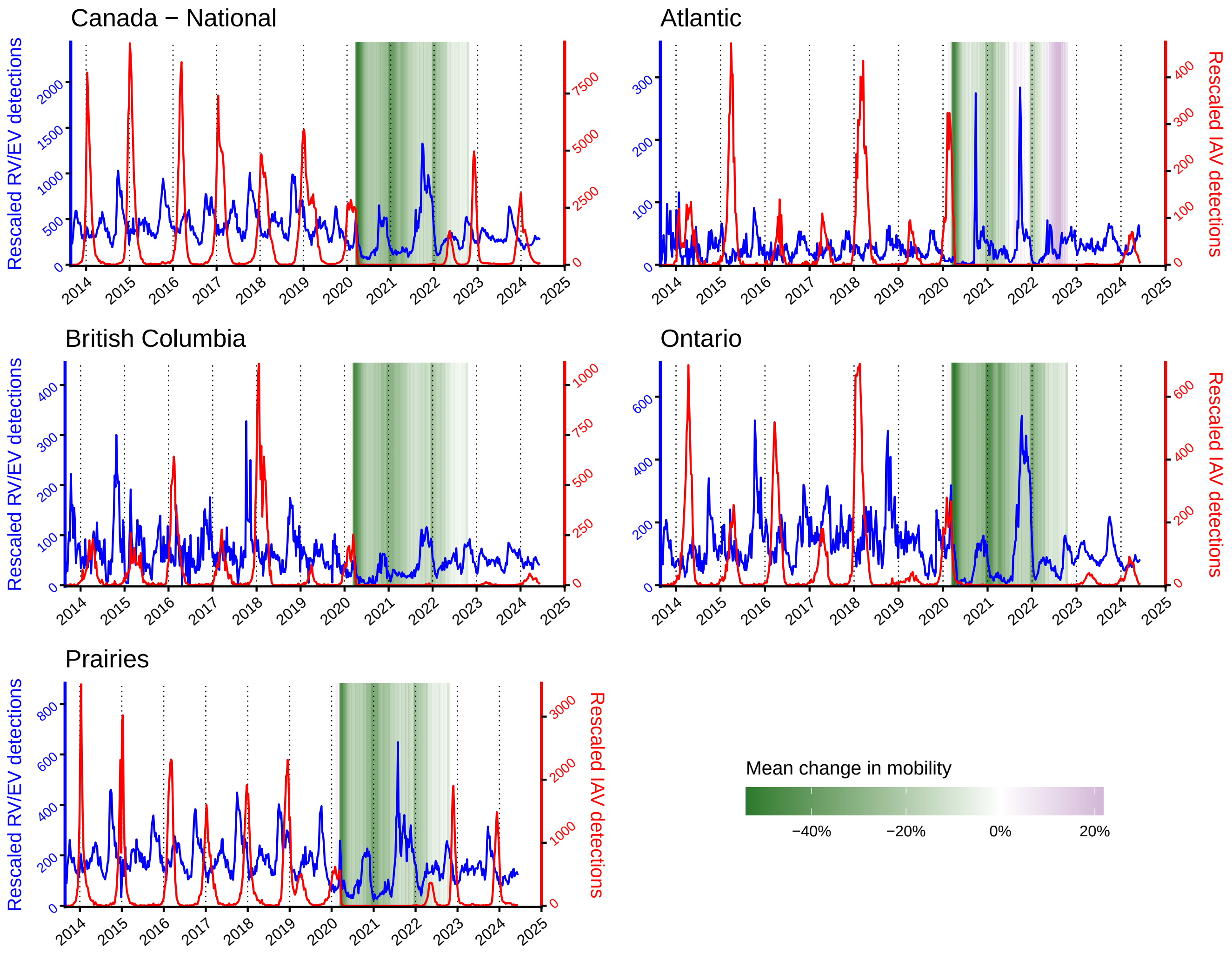

Figure S3: **RV and IAV time series (Canada).** RV/EV (blue) and IAV (red) detections in Canada (national and 4 provinces). Data were rescaled to account for changes in testing patterns (see **Fig. S4** and **S5**). Time series were reconstructed from publicly available reports by the RVDSS. Mean change in mobility (colored background) during the COVID-19 pandemic were computed from [4].

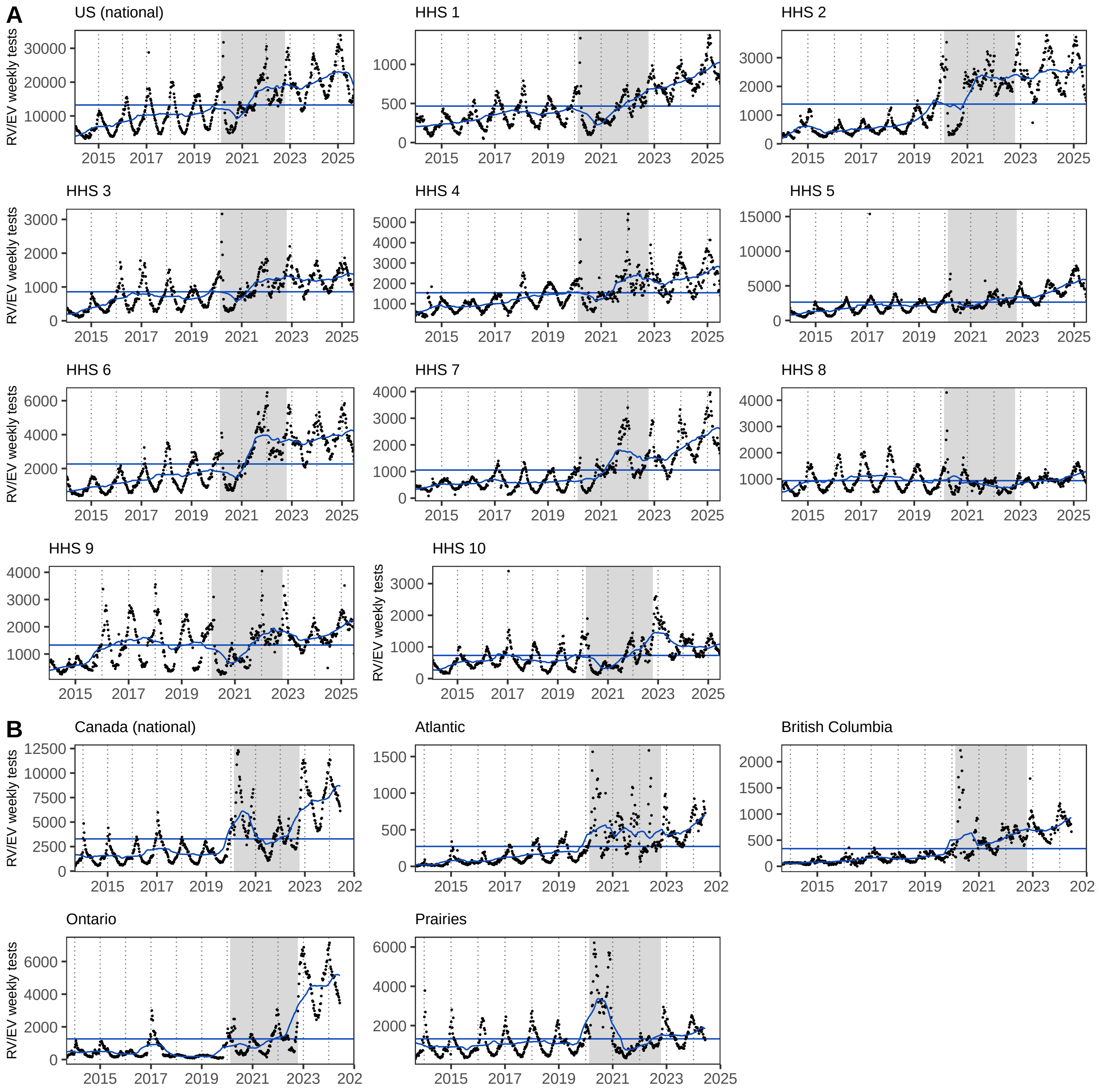

Figure S4: **RV testing patterns**. Weekly number of RV/EV tests (black points) in (A) the US and (B) Canada. Blue lines correspond to the 1-year moving average and mean number of tests over the studied period. To account for testing behavior, we rescaled incidence data by multiplying raw cases by a weekly testing factor equals to the average number of tests (over the entire studied period) divided by the one-year moving average of the number of tests. Grey backgrounds indicate the pandemic period.

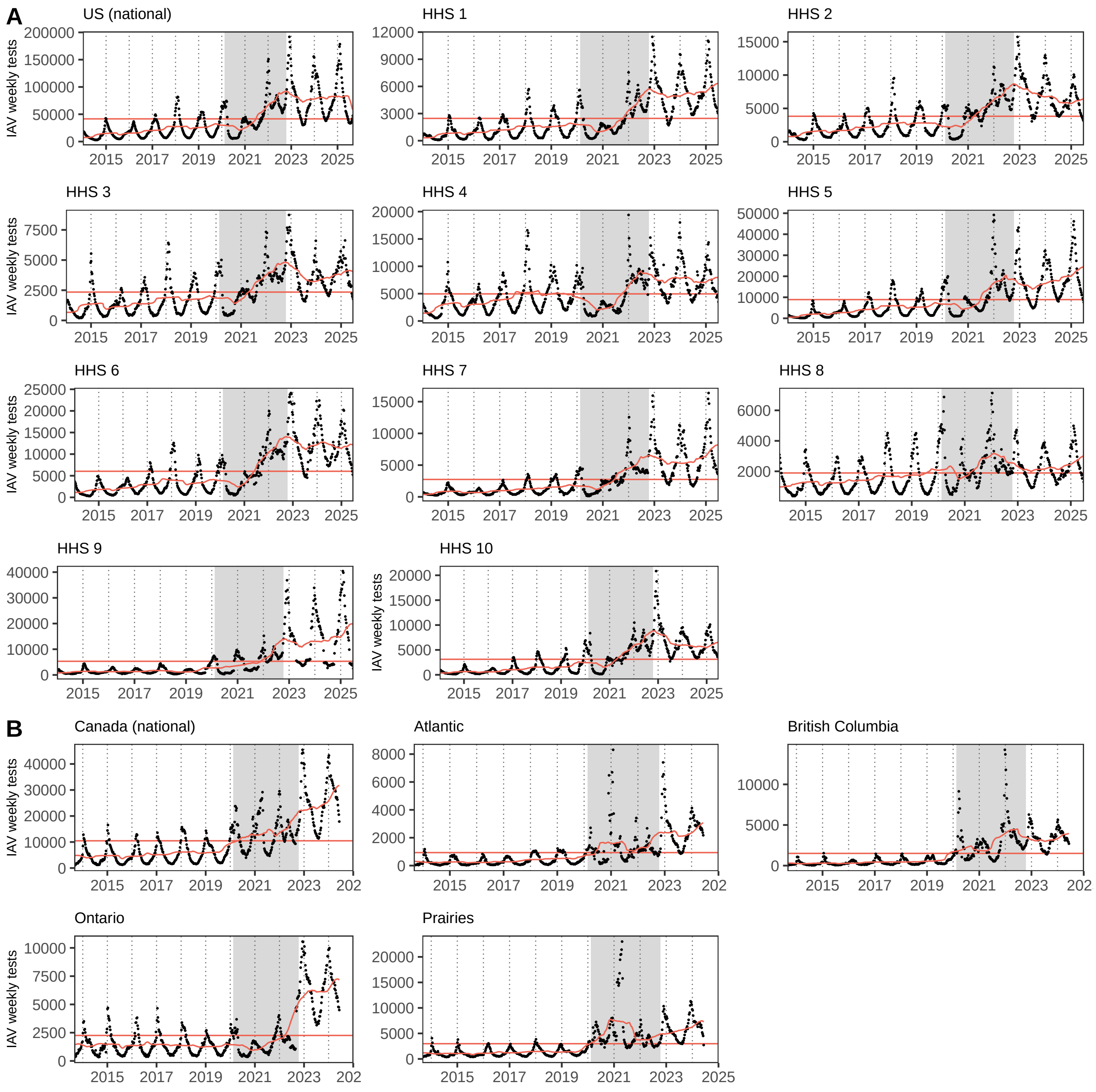

Figure S5: **IAV testing patterns**. Weekly number of IAV tests (black points) in (A) the US and (B) Canada. Red lines correspond to the one-year moving average and mean number of tests over the studied period. To account for testing behavior, we rescaled incidence data by multiplying raw cases by a weekly testing factor equals to the average number of tests (over the entire studied period) divided by the one-year moving average of the number of tests. Grey backgrounds indicate the pandemic period.

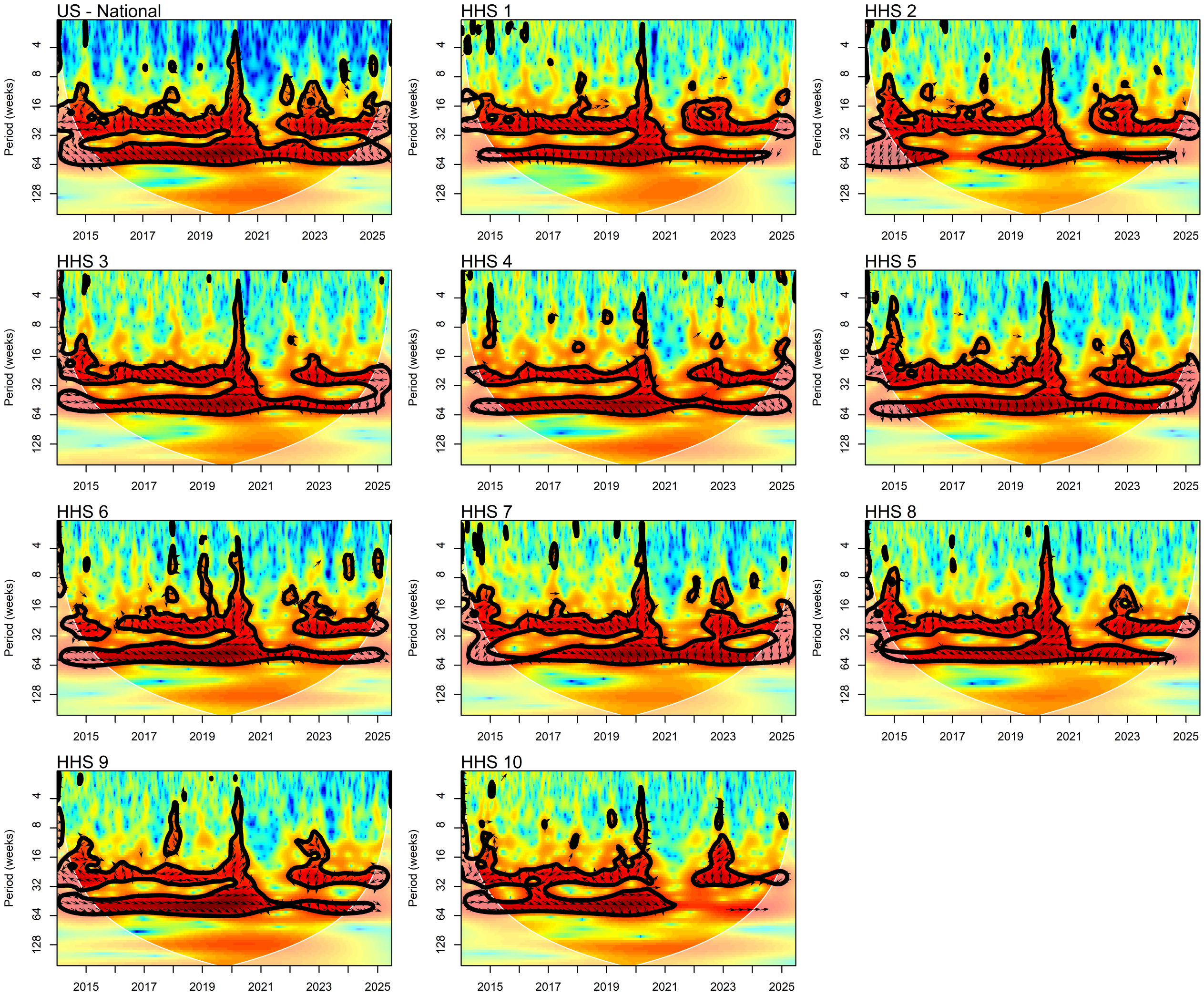

Figure S6: **Cross-wavelet transform of IAV and RV detections in the US.** We used the function `xwt` from the R package `biwavelet` [2] (see details in **SI Appendix §S1**). Colors indicate cross-wavelet power, with significant regions (95% confidence level) outlined by a contour. Arrows pointing right (resp. left) suggest IAV and RV detections are in phase (reps. out-of-phase) and arrows pointing up (resp. down) suggest IAV detections lead (resp. lag) RV detections (with vertical arrows indicating a phase difference of $\pi/2$ (quarter cycle)).

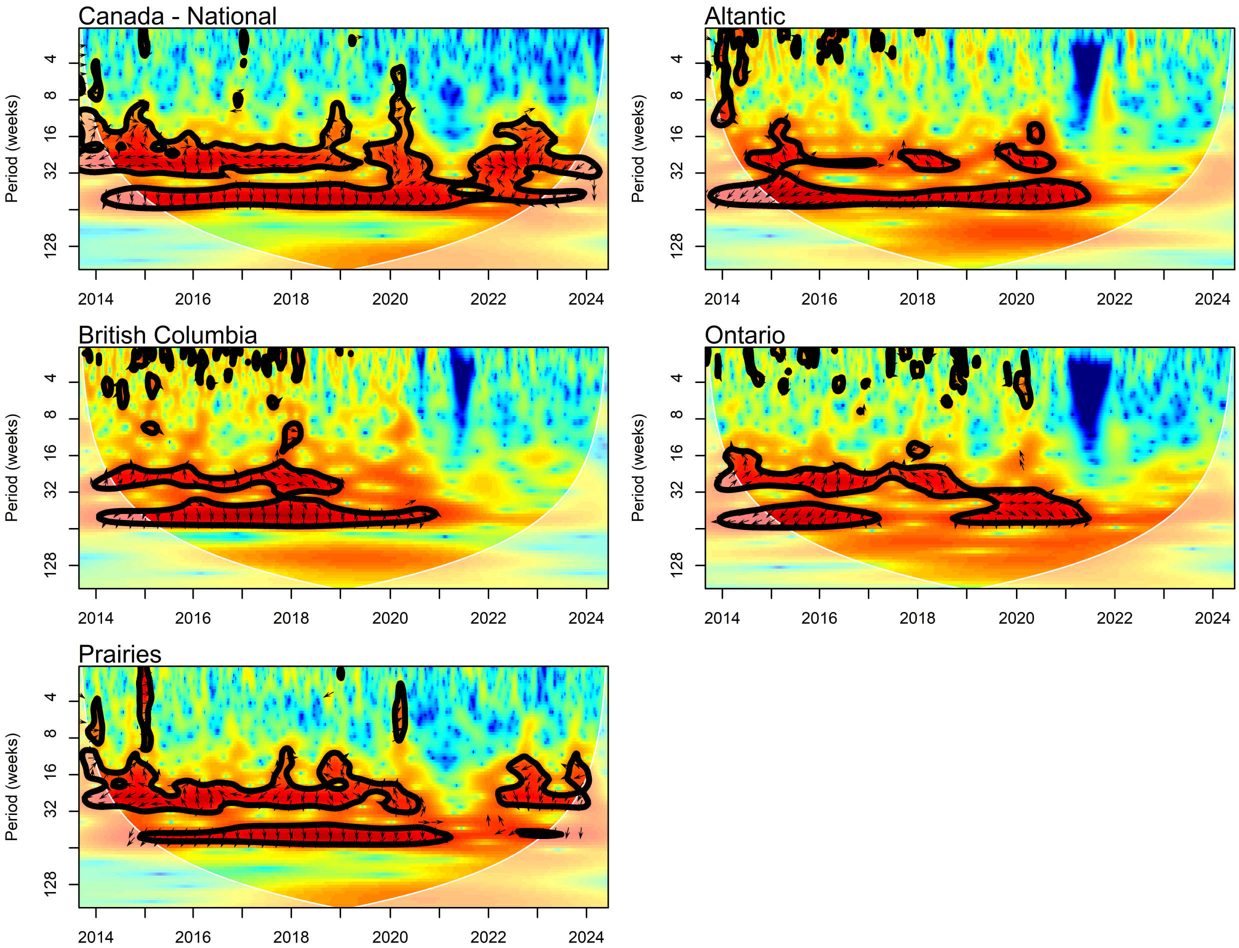

Figure S7: **Cross-wavelet transform of IAV and RV detections in Canada.** We used the function `xwt` from the R package `biwavelet` [2] (see details in **SI Appendix §S1**). Colors indicate cross-wavelet power, with significant regions (95% confidence level) outlined by a contour. Arrows pointing right (resp. left) suggest IAV and RV detections are in phase (reps. out-of-phase) and arrows pointing up (resp. down) suggest IAV detections lead (resp. lag) RV detections (with vertical arrows indicating a phase difference of $\pi/2$ (quarter cycle)).

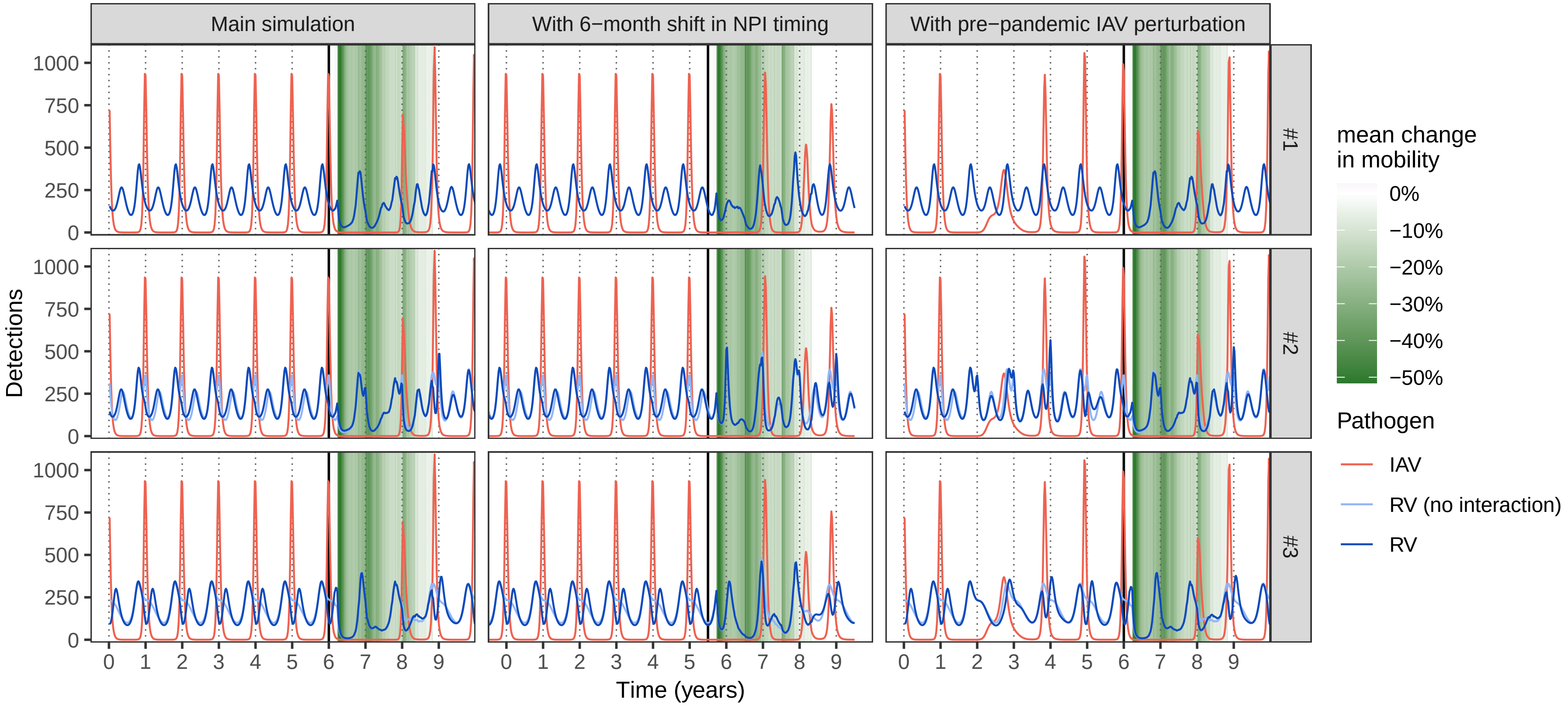

Figure S8: **Simulated dynamics.** We simulated RV (dark blue lines) and IAV dynamics (red lines) using model (S1) and parameter values listed in **Table S1**. We consider three main scenarios: 1, RV and IAV are independent ($\phi = 0$); 2, IAV negatively impacts RV ($\phi < 0$), but we adjust RV seasonal forcing so that its force of infection mimics that from scenario 1 when IAV is at its endemic attractor; 3, simple sinusoidal changes to RV transmission rate, with IAV negatively impacting RV ($\phi < 0$). See seasonal transmission profiles of RV and IAV in **Fig. 2-AB**. For scenarios 2 and 3, we also plot RV dynamics if there was no interaction due to IAV (light blue lines). In the first column (main simulation), we start from initial conditions on the endemic attractor of both pathogens, then simulate the model for 10 years, corresponding to a 6-year pre-pandemic period (stationary dynamics) followed by a 4-year (post-)pandemic period (vertical solid lines indicate the separation between the two periods). We used Google mobility data from Canada for the pandemic period. In the second column, we shift NPI timing 6-month. In the third column, we introduce a one-off exogenous perturbation perturbation in IAV dynamics (moving 35% of $S_{\text{IAV}}$ to $R_{\text{IAV}}$) at $t = 1.5$ year during the pre-pandemic period.

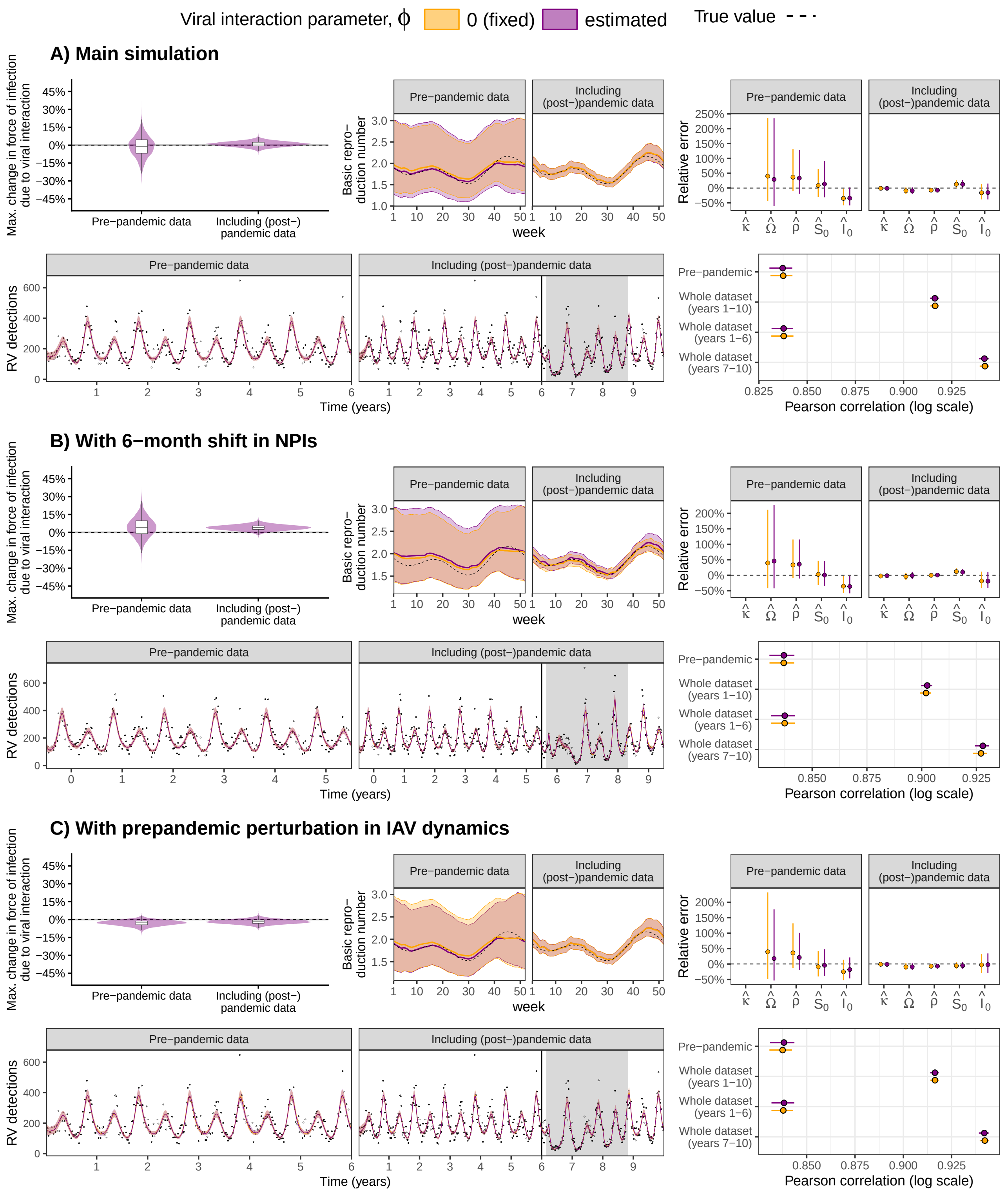

Figure S9: **Simulation study estimation results (scenario 1).** Here, RV and IAV are independent ($\phi = 0$). For each panel (A-C), we plot, from top to bottom and from left to right: (i) estimated effect of IAV (posterior distributions); (ii) estimated basic reproduction number of RV (median and 95% CrIs); (iii) relative errors of the remaining estimated parameters (median and 95% CrIs); (iv) simulated data (black points) and fitted values (median and 95% CrIs), where the vertical line marks the transition between pre- and (post-)pandemic periods and gray backgrounds, the period of implementation of COVID-19 NPIs; and (v) Pearson correlation coefficients (median and 95% CrIs) between simulated and fitted values on the log scale. True values are indicated by dashed lines. We fitted models assuming no viral interaction ($\phi = 0$, in yellow) or estimating the interaction term $\phi$ (purple). See **Fig. S8** for more details about the simulations.

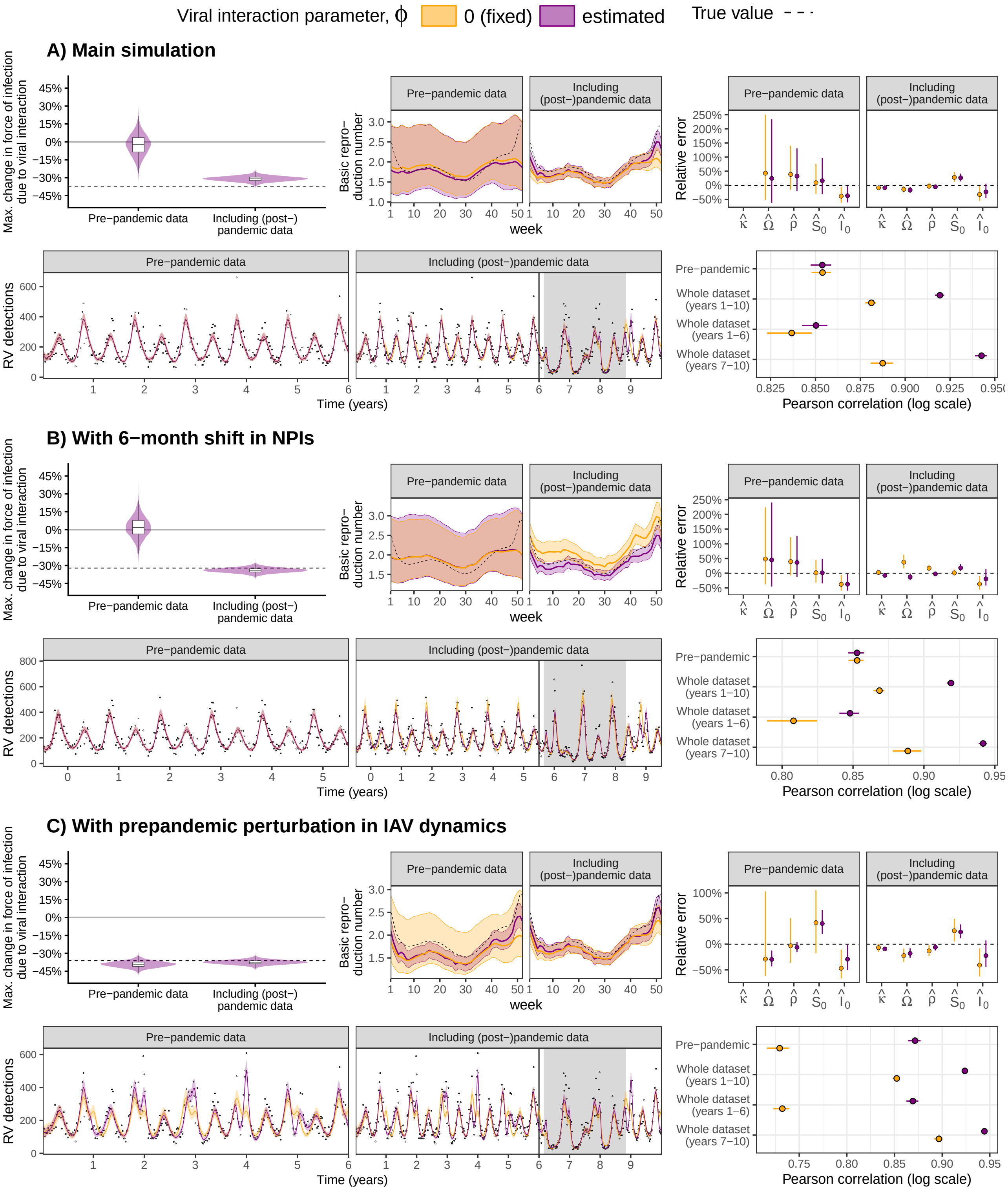

Figure S10: **Simulation study estimation results (scenario 2).** Here, IAV negatively impacts RV ($\phi < 0$). For each panel (A-C), we plot, from top to bottom and from left to right: (i) estimated effect of IAV (posterior distributions); (ii) estimated basic reproduction number of RV (median and 95% CrIs); (iii) relative errors of the remaining estimated parameters (median and 95% CrIs); (iv) simulated data (black points) and fitted values (median and 95% CrIs), where the vertical line marks the transition between pre- and (post-)pandemic periods and gray backgrounds, the period of implementation of COVID-19 NPIs; and (v) Pearson correlation coefficients (median and 95% CrIs) between simulated and fitted values on the log scale. True values are indicated by dashed lines. We fitted models assuming no viral interaction ($\phi = 0$, in yellow) or estimating the interaction term $\phi$ (purple). See **Fig. S8** for more details about the simulations.

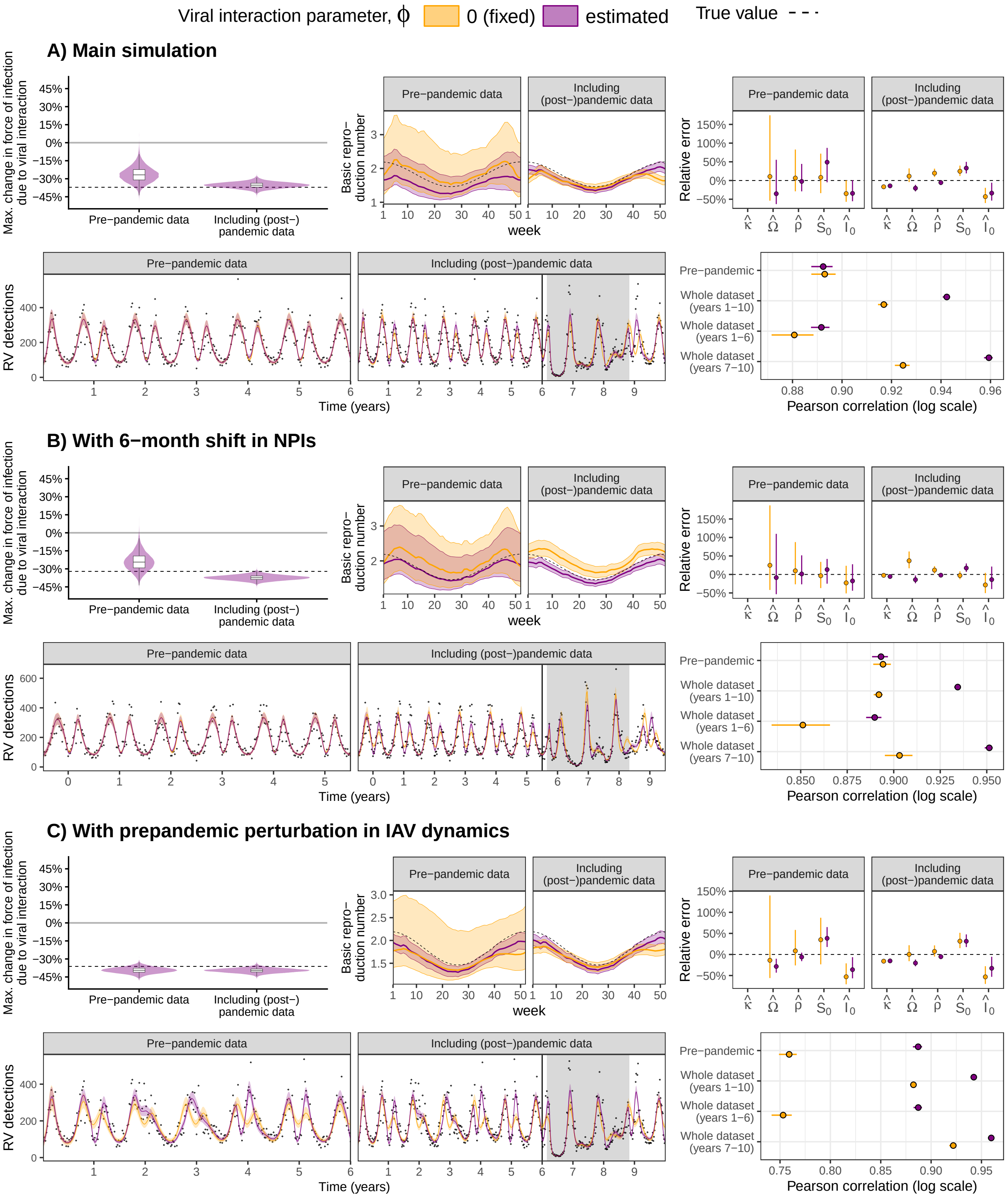

Figure S11: **Simulation study estimation results (scenario 3).** Here, IAV negatively impacts RV ($\phi < 0$). For each panel (A-C), we plot, from top to bottom and from left to right: (i) estimated effect of IAV (posterior distributions); (ii) estimated basic reproduction number of RV (median and 95% CrIs); (iii) relative errors of the remaining estimated parameters (median and 95% CrIs); (iv) simulated data (black points) and fitted values (median and 95% CrIs), where the vertical line marks the transition between pre- and (post-)pandemic periods and gray backgrounds, the period of implementation of COVID-19 NPIs; and (v) Pearson correlation coefficients (median and 95% CrIs) between simulated and fitted values on the log scale. True values are indicated by dashed lines. We fitted models assuming no viral interaction ($\phi = 0$, in yellow) or estimating the interaction term $\phi$ (purple). See **Fig. S8** for more details about the simulations.

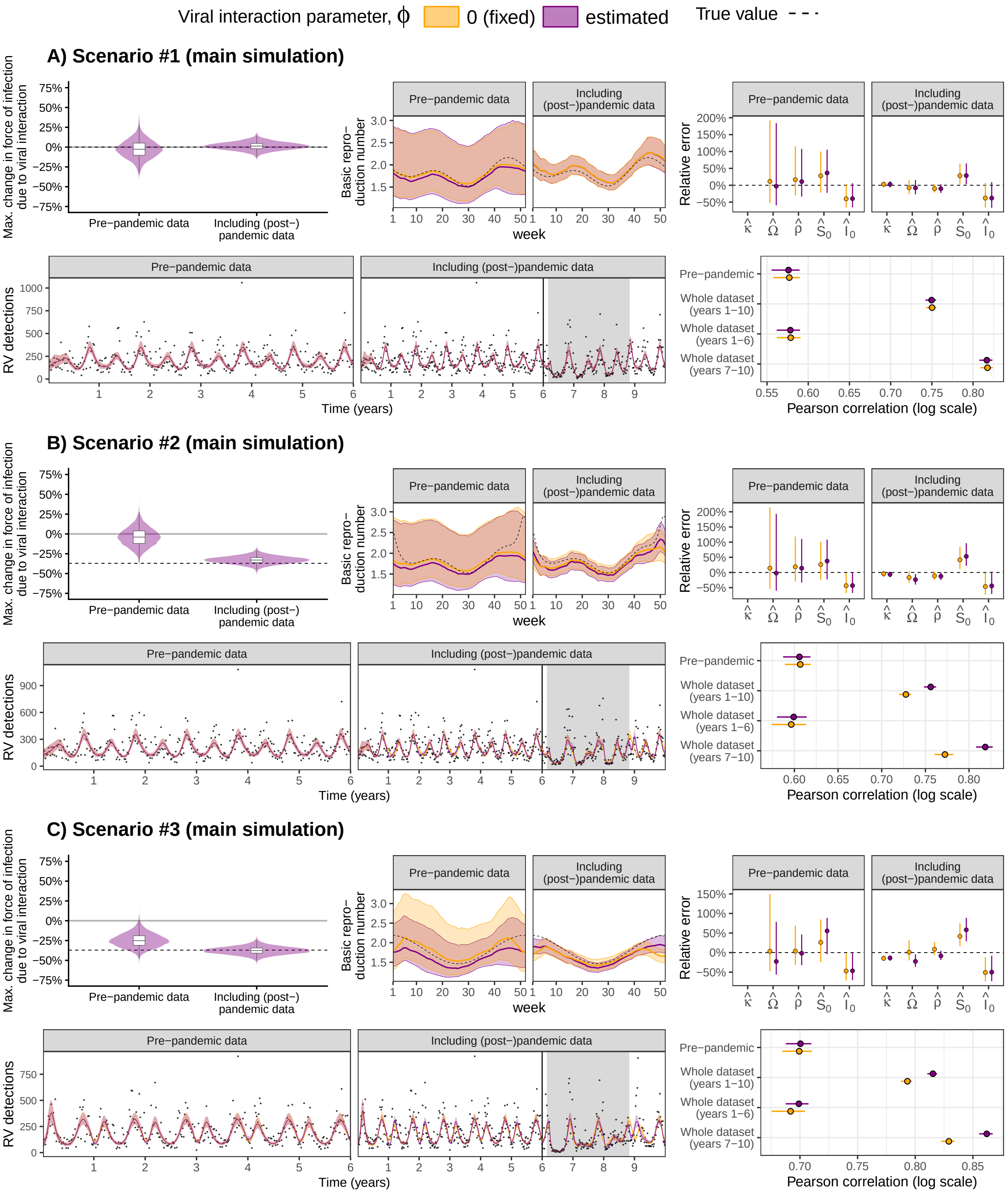

Figure S12: **Simulation study estimation results with high observation noise.** We assessed the robustness of our results to the observation noise, generating simulated data with $\sigma = 0.5$ (instead of $\sigma = 0.25$). For each panel (A-C), we plot, from top to bottom and from left to right: (i) estimated effect of IAV (posterior distributions); (ii) estimated basic reproduction number of RV (median and 95% CrIs); (iii) relative errors of the remaining estimated parameters (median and 95% CrIs); (iv) simulated data (black points) and fitted values (median and 95% CrIs), where the vertical line marks the transition between pre- and (post-)pandemic periods and gray backgrounds, the period of implementation of COVID-19 NPIs; and (v) Pearson correlation coefficients (median and 95% CrIs) between simulated and fitted values on the log scale. True values are indicated by dashed lines. We fitted models assuming no viral interaction ($\phi = 0$, in yellow) or estimating the interaction term $\phi$ (purple).

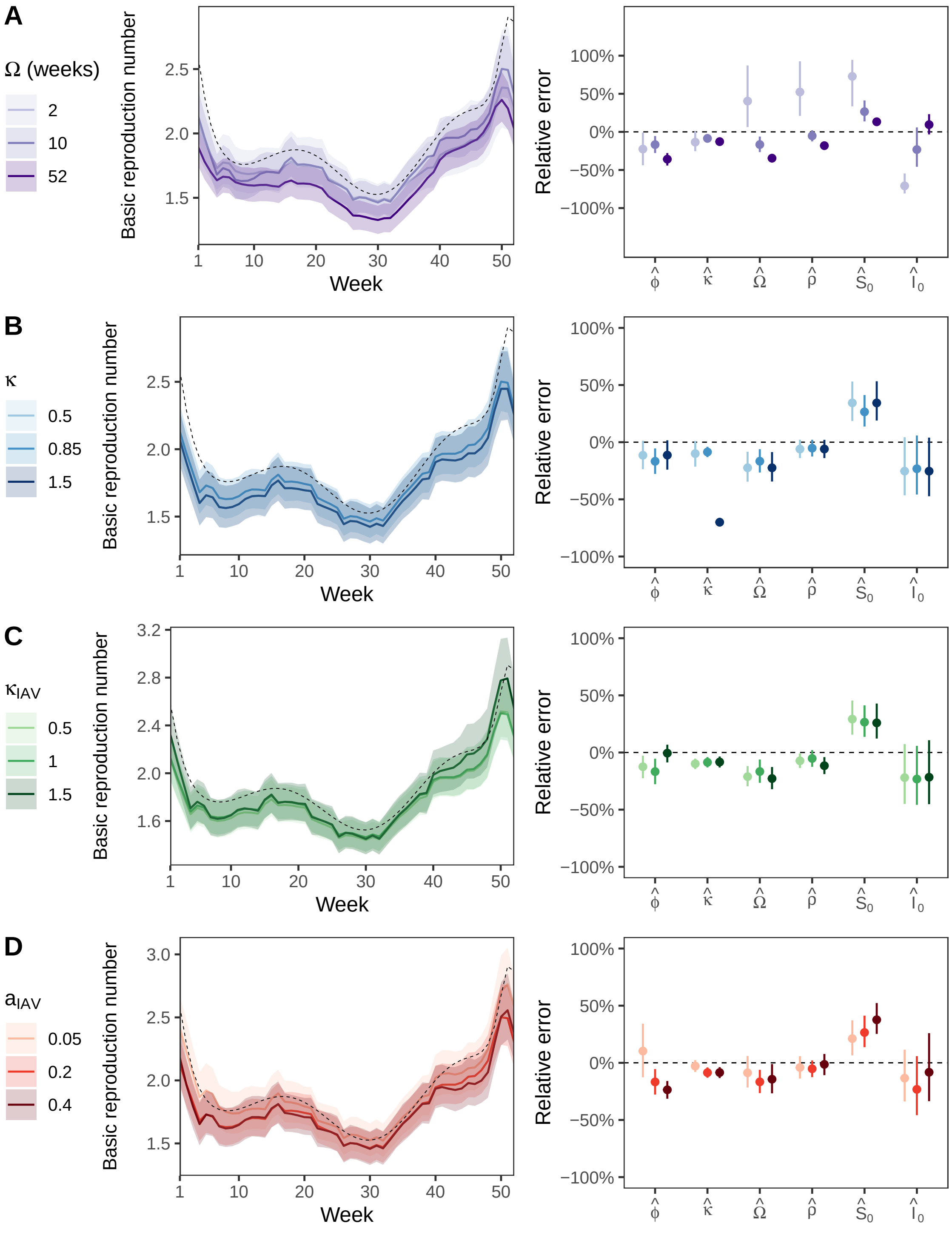

Figure S13: **Sensitivity of estimates to chosen parameter values.** Built upon the main simulation of scenario 2 ("masked interaction"), we explored the sensitivity of our estimates to some key parameter values of the viruses. We varied (A) the mean duration of immune protection to RV reinfection $\Omega$, (B) the strength of the effect of NPIs on RV transmission $\kappa$ and (C) on IAV transmission $\kappa_{\text{IAV}}$ and (D) the amplitude of the seasonal forcing of IAV $a_{\text{IAV}}$. For each parameter value, we regenerated simulated data and fitted the model with potential viral interaction to the whole dataset (i.e., including the (post-)pandemic period). Parameter estimates remained robust across the explored range of values, except maybe parameters $\Omega$, $\rho$ and $S_0$ that were all overestimated when $\Omega$ was very short (fixed 2 weeks), and parameters $\kappa$ that were quite underestimated when its value was high (fixed to 1.5). Solid lines and points denote median values, while envelopes and segments denote 95% CrIs; true values are indicated by dashed lines.

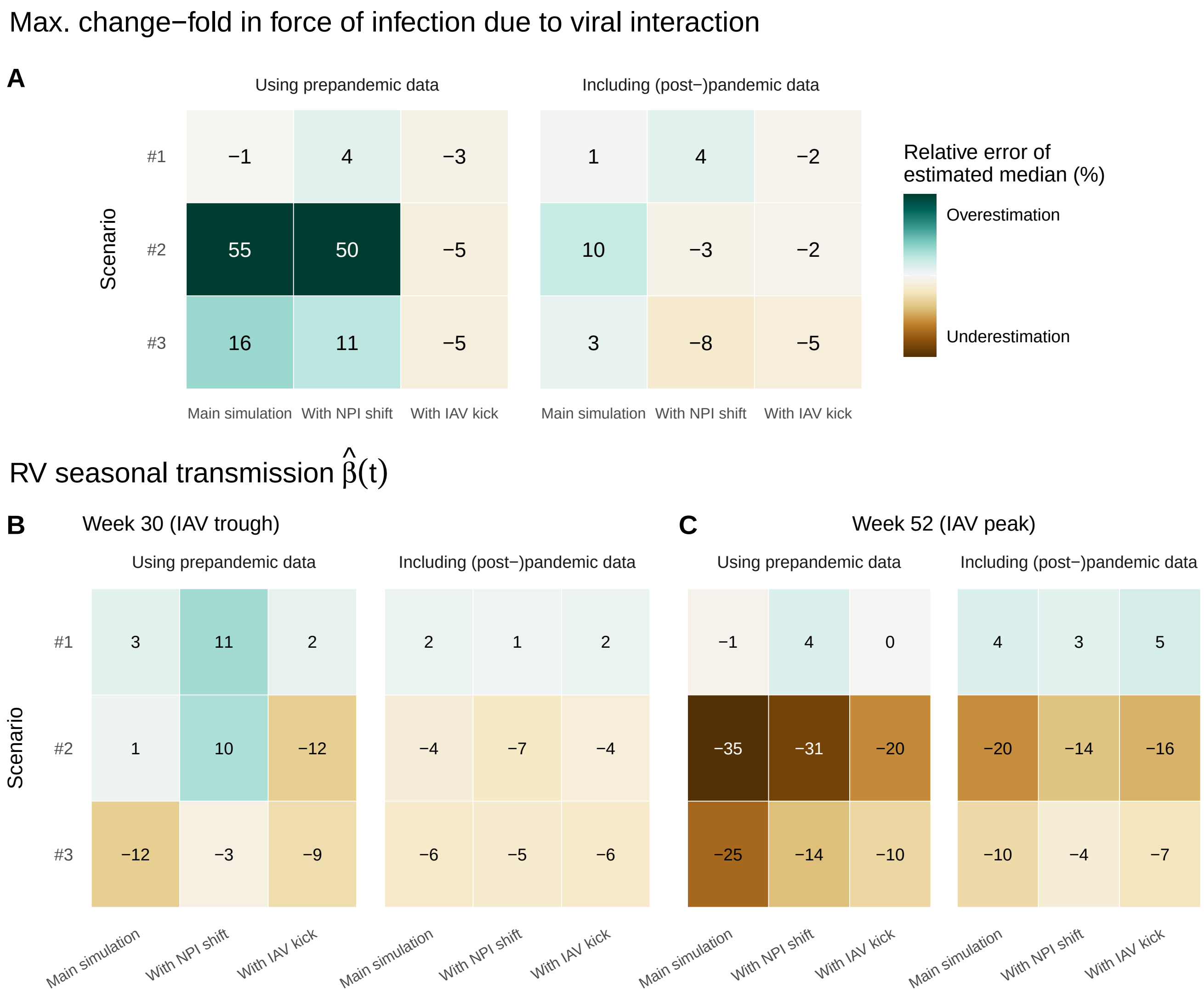

Figure S14: **Relative errors in estimates of viral interaction and seasonal forcing using simulated data.** We compared the relative errors in the estimated median of (A) the maximum change-fold in RV force of infection due to viral interaction of IAV $\left(1+\hat{\phi}\right)$ and (B) RV seasonal transmission $\hat{\beta}(t)$ at the $30^{\text{th}}$ week (when IAV circulation is minimal) and $52^{\text{th}}$ week (when IAV circulation is maximal). For each case, we present results using pre-pandemic data and including (post-)pandemic data. In each heatmap, rows represent the three scenarios and columns represent the corresponding main simulation and two variations: one where the timing of NPI occurs 6-month earlier (*With NPI shift*) and one where we introduce a one-off exogenous perturbation in IAV dynamics, moving 35% of the susceptibles to the recovered compartment during the pre-pandemic period (*With IAV kick*).

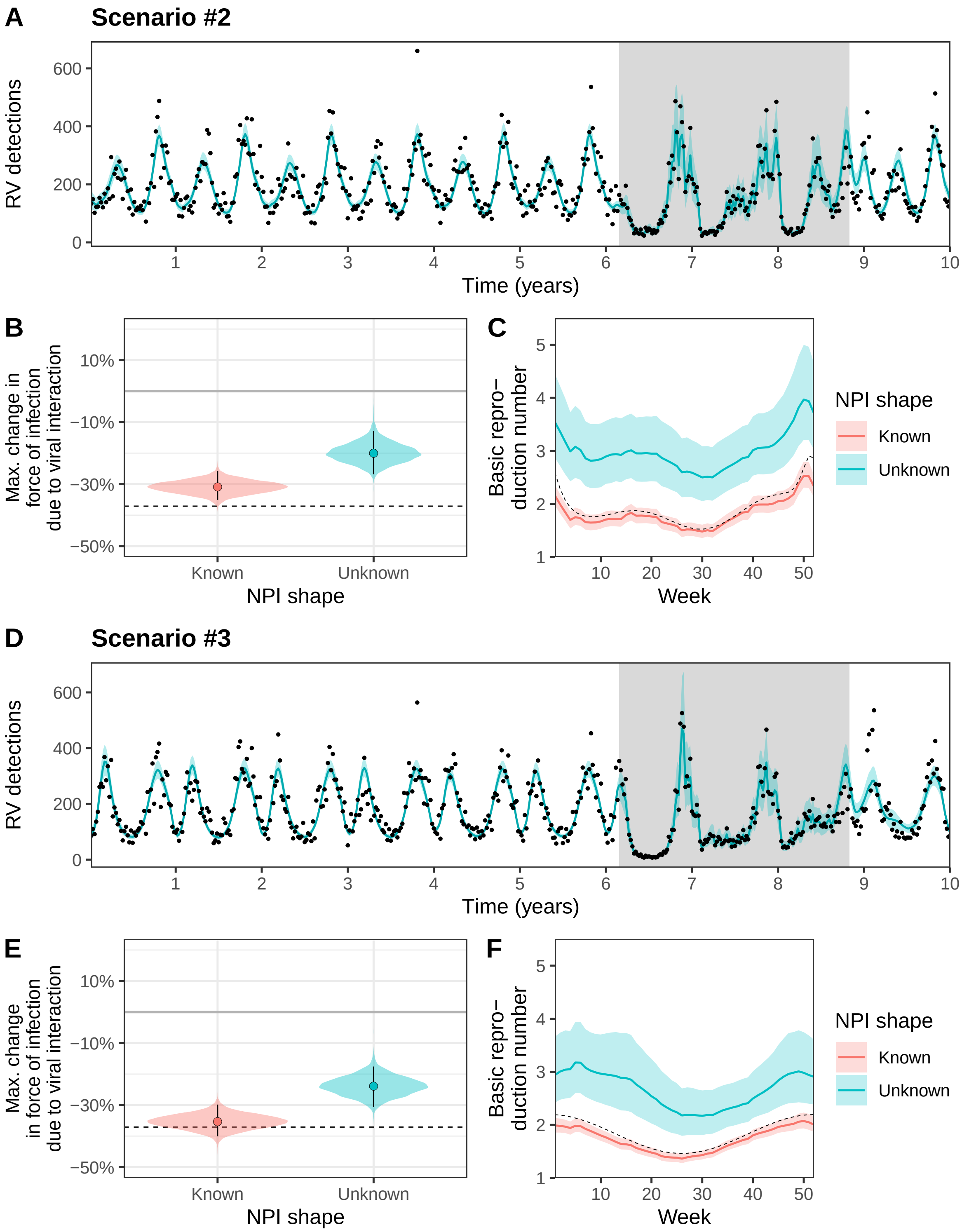

Figure S15: **Unknown shape of NPI perturbations can hamper the inference of viral interaction and seasonal forcing.** Here, we use simulated data from the main simulations of scenarios 2 (A-C) and 3 (D-F); in both scenarios IAV negatively impacts RV ($\phi = 0$). We contrast results obtained when constraining the shape of NPIs (in red, using Google mobility data) or not (in blue). In the latter, we estimated the NPI effect of each week of the pandemic period (gray background in A and C) using a uniform prior between 0 and 1. (A,D) Median (lines) and 95% CrIs (envelopes) of the fitted values when the shape of NPIs is unknown and estimated. (B,E) Posterior distributions, median (point) and 95% CrIs (segments) of the maximum interaction effect due to IAV and (C,F) Median (lines) and 95% CrIs (envelopes) of RV basic reproduction (proportional to seasonal forcing). True values are indicated by dashed lines.

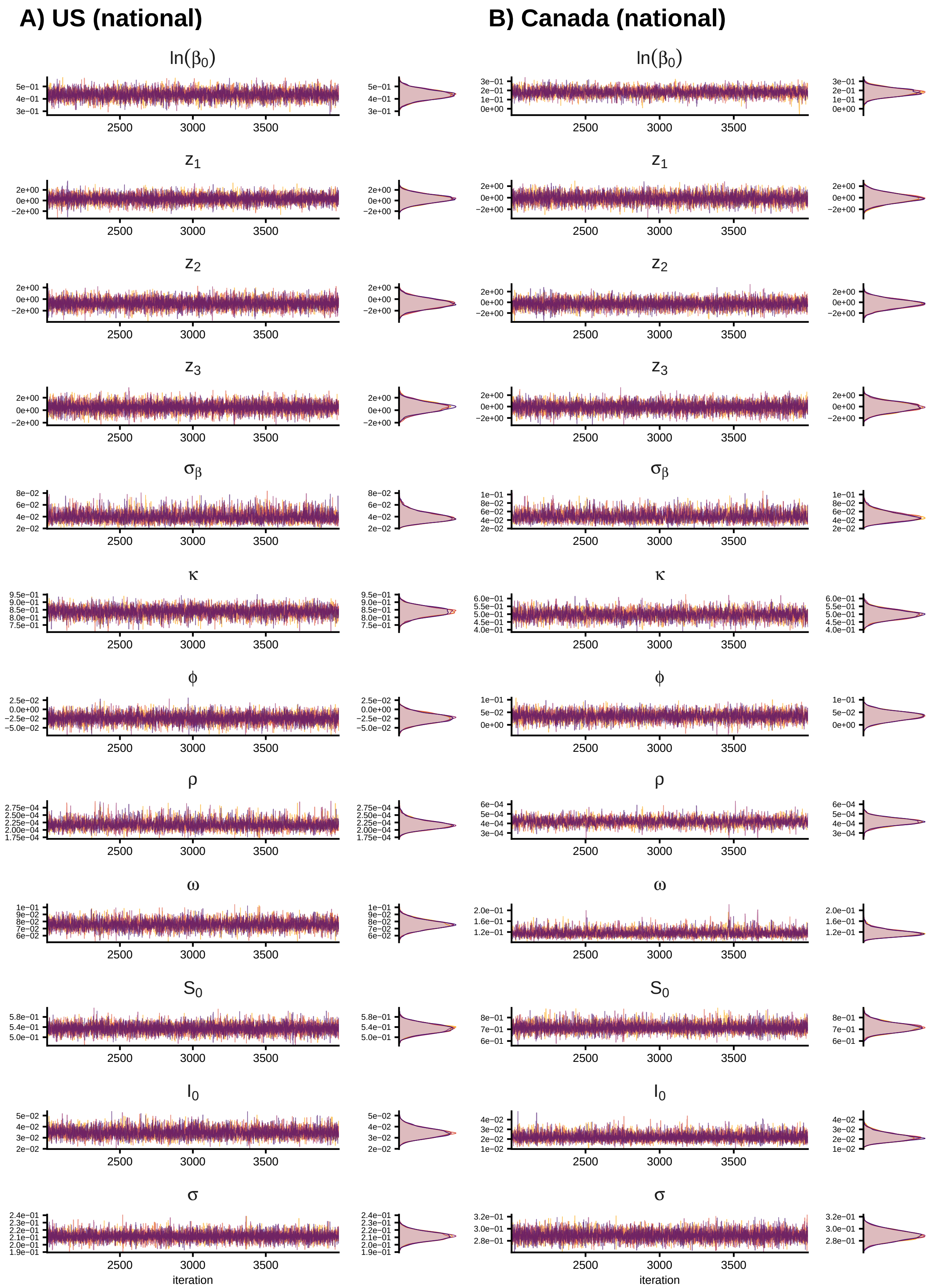

Figure S16: **Examples of MCMC chains and posterior distributions.** Examples from the model fitted to national-level time series from (A) US and (B) Canada. The first 2,000 iterations (burn-in periods) are not represented.

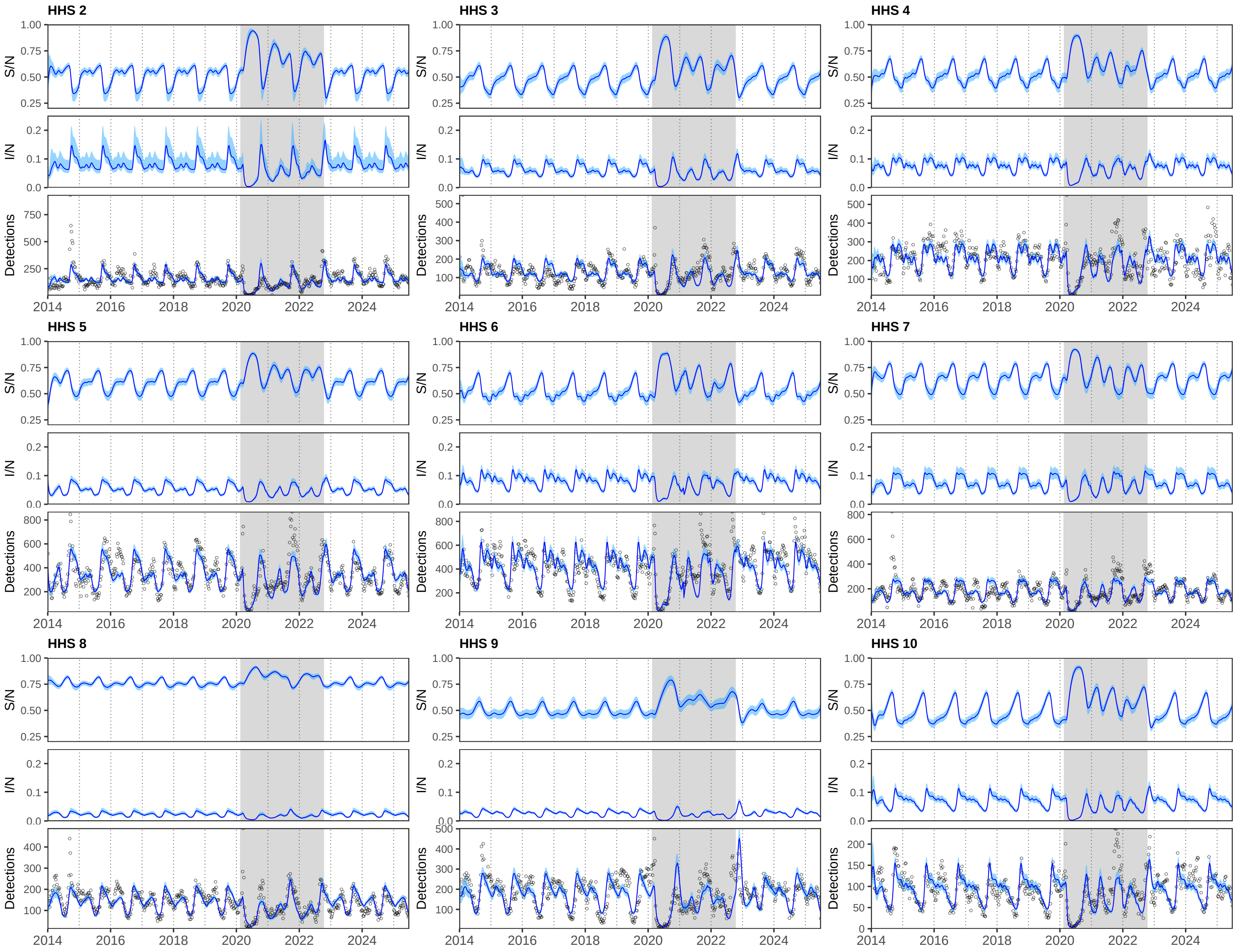

Figure S17: **Fitted model results in the US at the regional level.** For HHS regions 2-10, we independently fitted model (3)-(4) with seasonal transmission (8) (see **Materials and Methods §4.2-4.3**) to rescaled RV/EV detections (black points); we only show results obtained with the model with no viral interaction ($\phi = 0$). We plot posterior median values (lines) and 95% CrIs (envelopes) for (top) the proportion susceptible $S/N$, (middle) the proportion infected $I/N$ and (bottom) the number of detections. Grey backgrounds indicate the pandemic period.

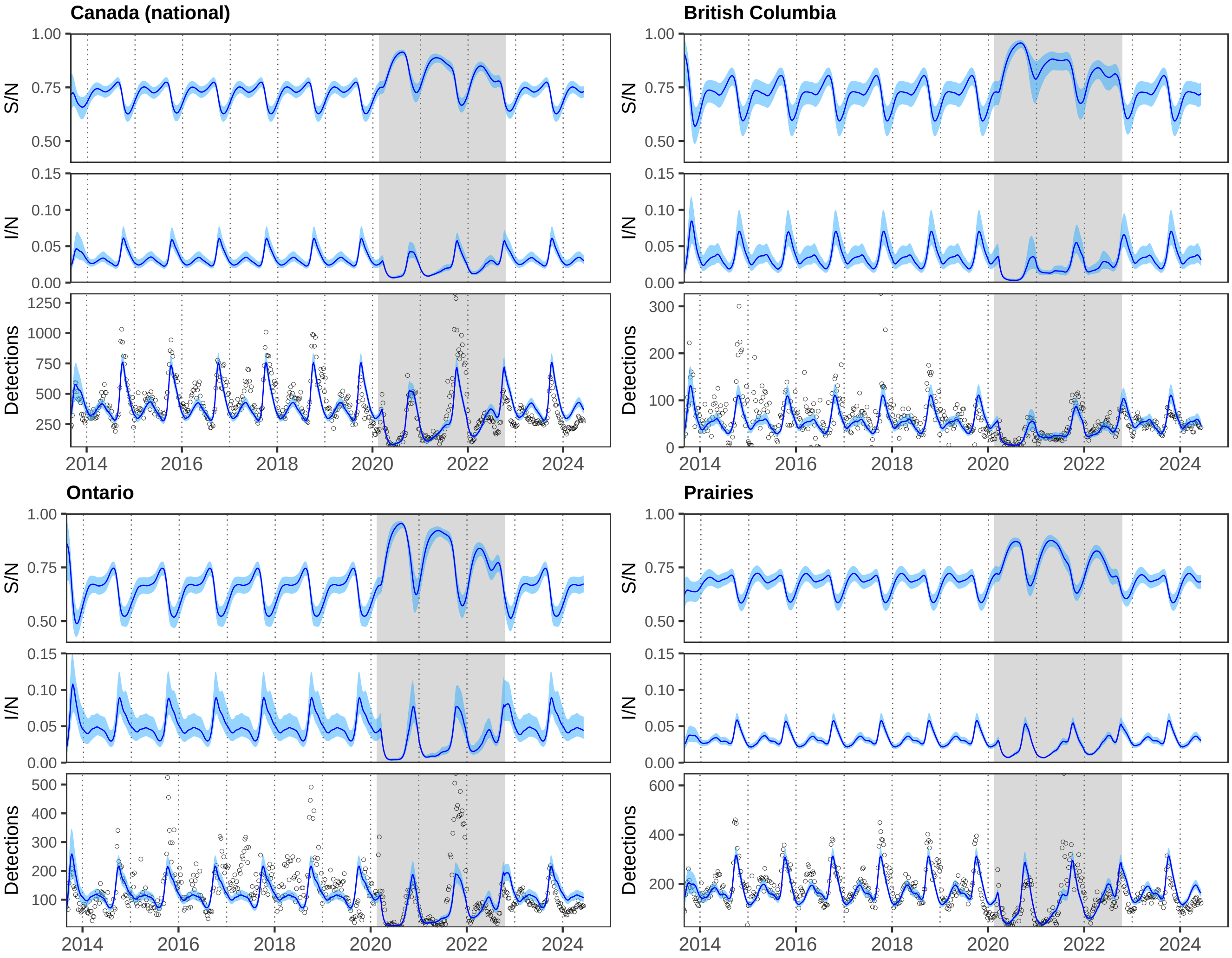

Figure S18: **Fitted model results in Canada at the national and provincial/regional level.** For each location, we independently fitted model (3)-(4) with seasonal transmission (8) (see **Materials and Methods §4.2-4.3**) to rescaled RV/EV detections (black points); we only show results obtained with the model with no viral interaction ($\phi = 0$). We plot posterior median values (lines) and 95% CrIs (envelopes) for (top) the proportion susceptible $S/N$, (middle) the proportion infected $I/N$ and (bottom) the number of detections. Grey backgrounds indicate the pandemic period.

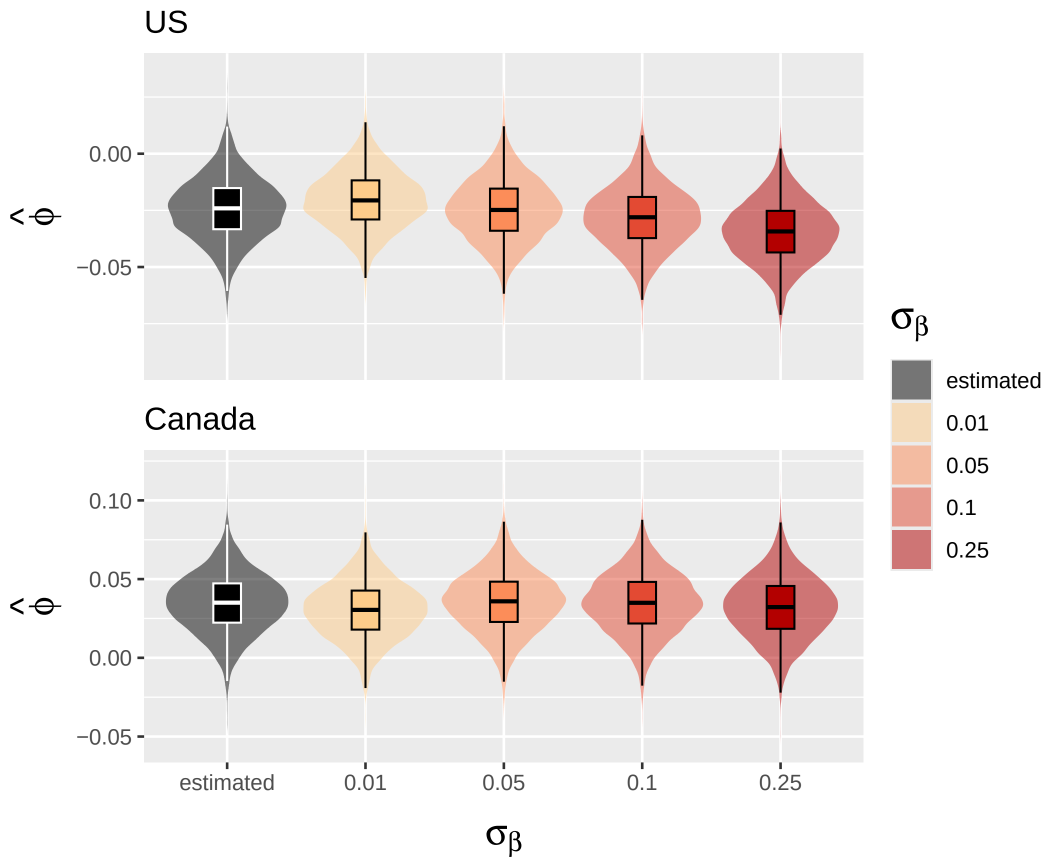

Figure S19: **Sensitivity analysis to $\sigma_\beta$.** We assessed the robustness of the inferred posterior distribution of the viral interaction parameter $\phi$ to the prior constraint on seasonal transmission variability. To do so, we refitted the model to national-level data while fixing $\sigma_\beta$ – the scale parameter governing the magnitude of weekly increments in the random walk for the seasonal transmission rate – at increasing values from 0.01 to 0.25.

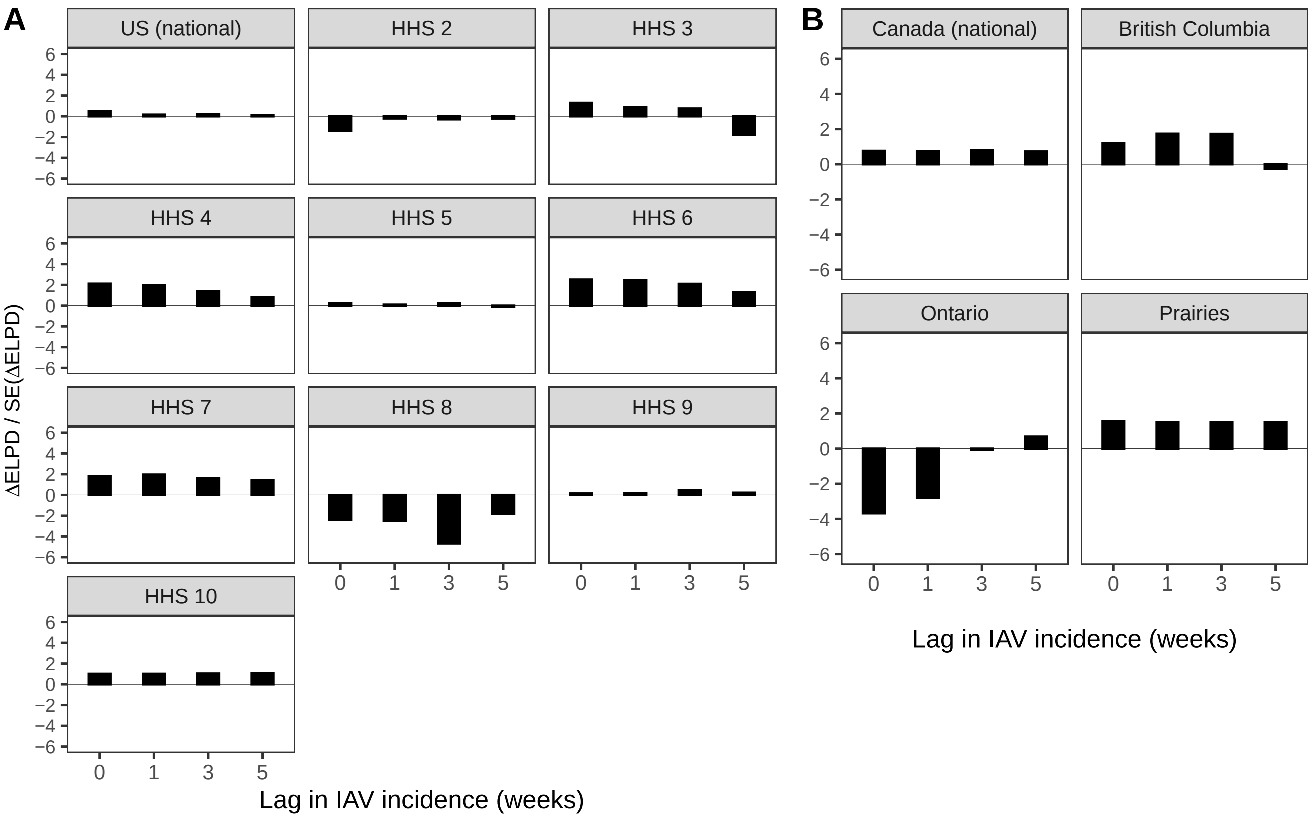

Figure S20: **Model comparisons.** We computed approximate leave-one-out cross-validation using Pareto smoothed importance sampling, as implemented in the R package `loo` [5] (we evaluated exact log-likelihood for the very few observations with Pareto k-values $> 0.7$). Fitted models allowing for potential viral interaction – using current IAV incidence (lag=0) or lagged sum of IAV incidence of 1, 3 or 5 weeks – were compared against a null model with no interaction ($\phi = 0$). Comparisons are based on differences in expected log-predictive density ΔELPD relative to the null model, normalized by their standard error SE(ΔELPD). Absolute values below 2–4 are generally considered insufficient to provide strong evidence favoring the model with higher ELPD, in which case the more parsimonious model is preferred.

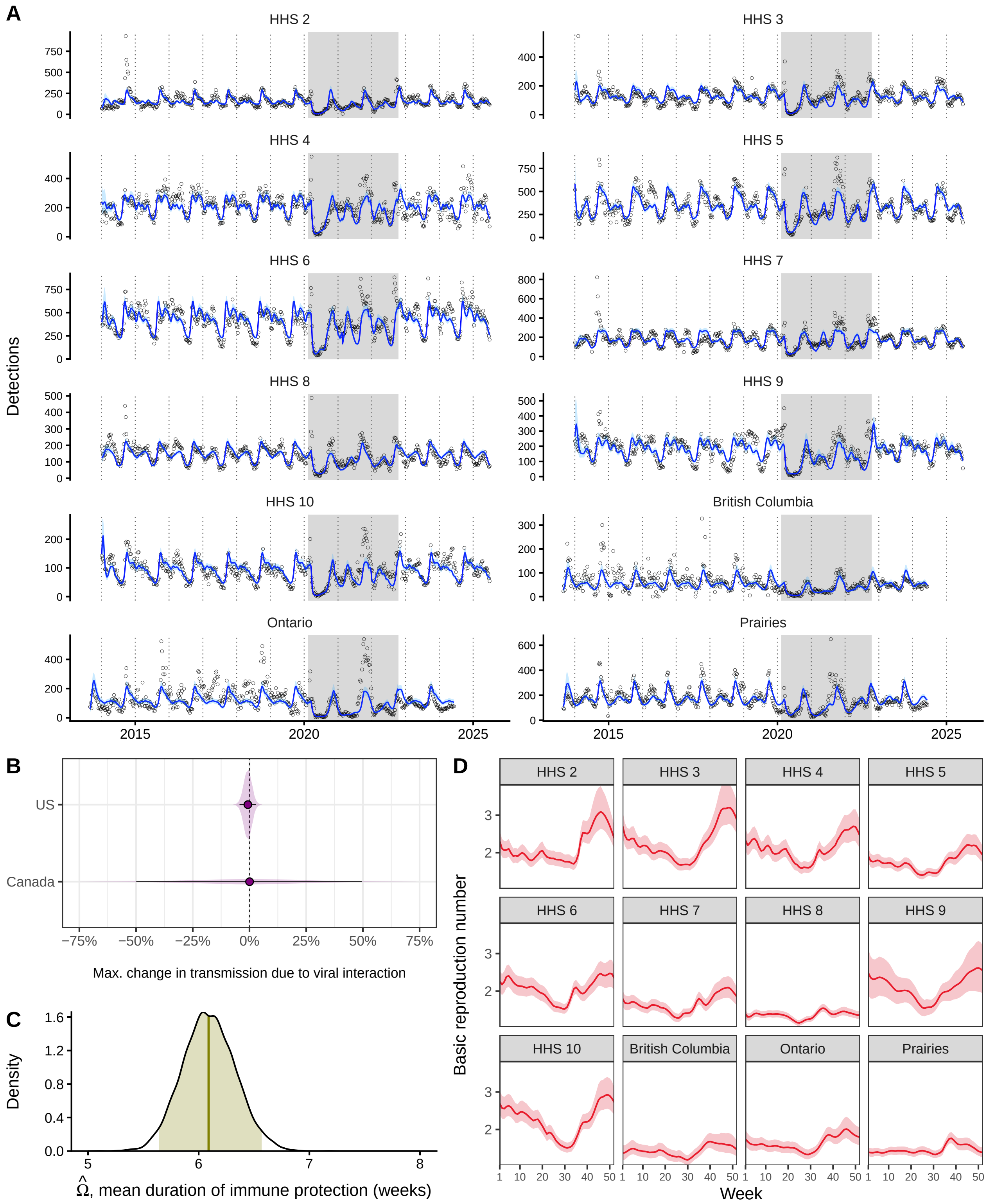

Figure S21: **Joint model fit across all locations.** Using IAV current incidence, we also fitted the model simultaneously to all regions/provinces, constraining the rate of immune waning $\omega$ to be shared across all locations and the viral interaction parameter $\phi$ across locations within a given country (US vs. Canada). See more details in **supporting information §S3**. (A) Model fit to RV/EV detections (black points). (B) Posterior distributions (with median values (points) and 95% CrIs (segments)) of the maximum change in RV transmission due to IAV interaction for US and Canadians locations. (C) Posterior distribution of the mean duration of immune protection $\Omega$ (same for all locations). (D) Estimated profiles of RV basic reproduction number $\mathcal{R}_0$. Lines denote median values and envelopes denote 95% CrIs. Grey backgrounds indicate the pandemic period.

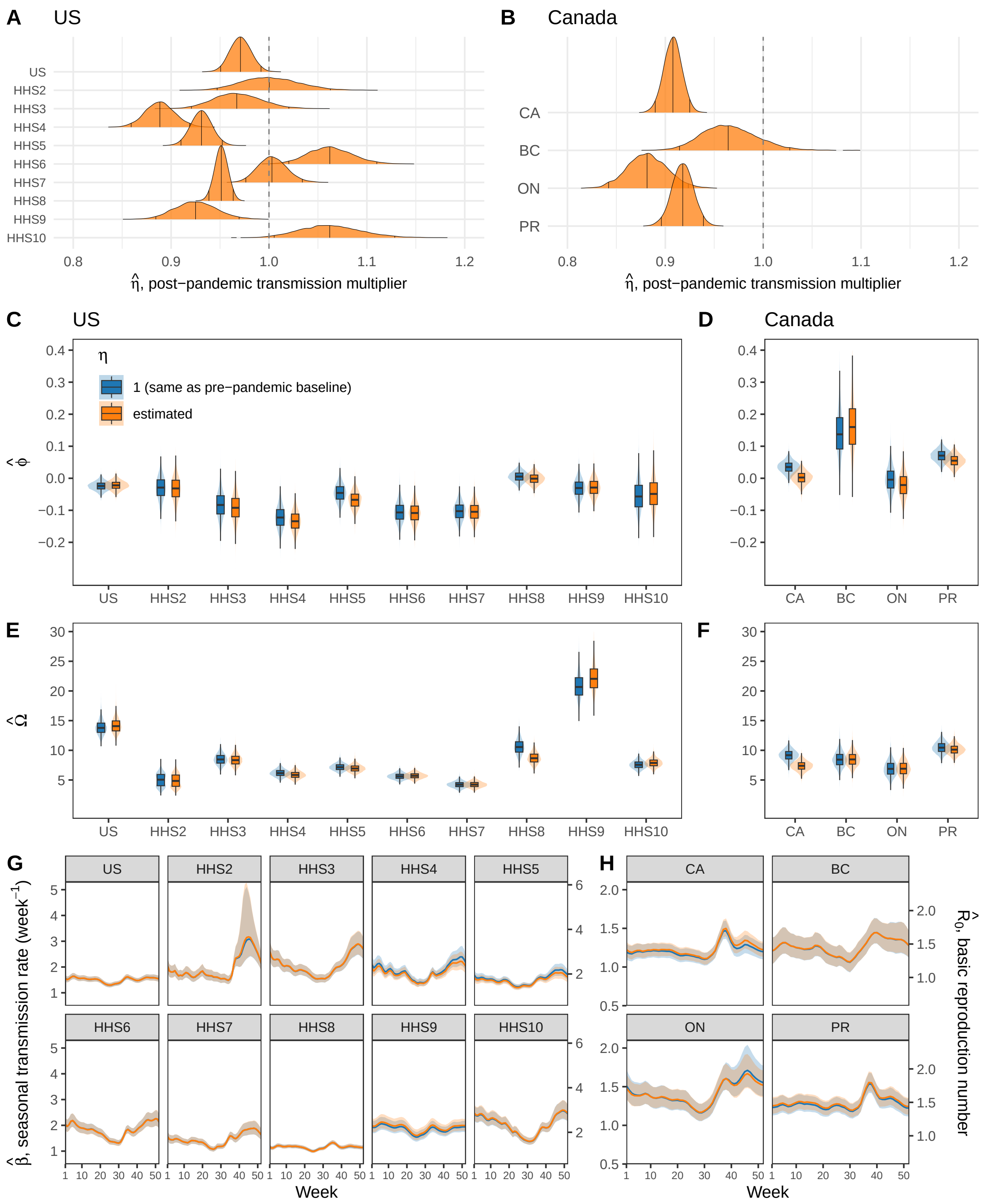

Figure S22: **Sensitivity to post-pandemic behavioral changes.** Google COVID-19 mobility data were no longer reported after October 15, 2022. In the main analysis, we assumed $c = 0$ (i.e., contacts are the same as in the pre-pandemic period) after this date. However, as NPIs were relaxed, new behavioral norms may have stabilized at non-baseline levels. We assessed the robustness of our results to this assumption by estimating a (constant) post-pandemic transmission multiplier, $\eta$, such that the seasonal transmission rate becomes $\eta\,\beta(t)$ after October 15, 2022 (in the main analysis, $\eta = 1$), with prior $\eta \sim \text{Log-}\mathcal{N}(0, 0.4)$. Estimated values were close to 1 (i.e., similar to pre-pandemic baseline) and had little impact on the estimates of the other parameters. (A-B) Posterior distributions of $\eta$; vertical black lines represent 2.5%, 50% and 97.5% quantiles, respectively. (C-H) Comparison of the posterior distributions of (C,D) viral interaction parameter $\phi$, (E,F) mean duration of immune protection $\Omega$ (expressed in weeks) and (G,H) of median (lines) and 95% CrIs (envelopes) of seasonal transmission profiles. CA=Canada (national), BC=British Columbia, ON=Ontario, PR=Prairies.

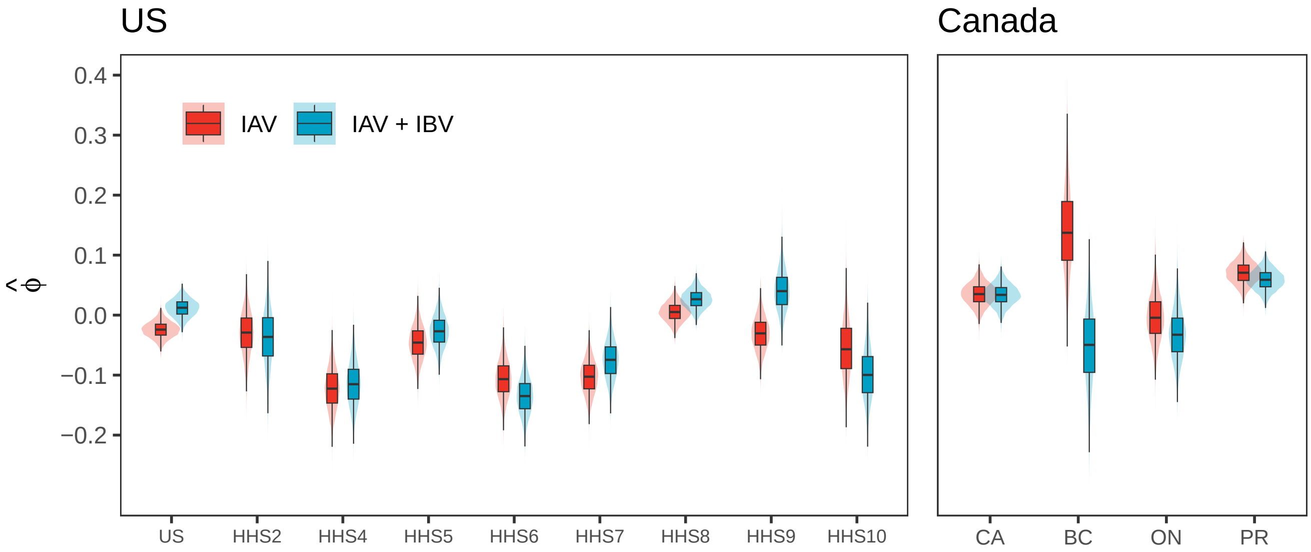

Figure S23: **The effect of including IBV on viral interaction estimates in the US and Canada.** Posterior distributions of the viral interaction parameter $\phi$ estimated using IAV current incidence (same as in the main text, in red) and combined IV current incidence (IAV+IBV, in blue). CA=Canada (national), BC=British Columbia, ON=Ontario, PR=Prairies.

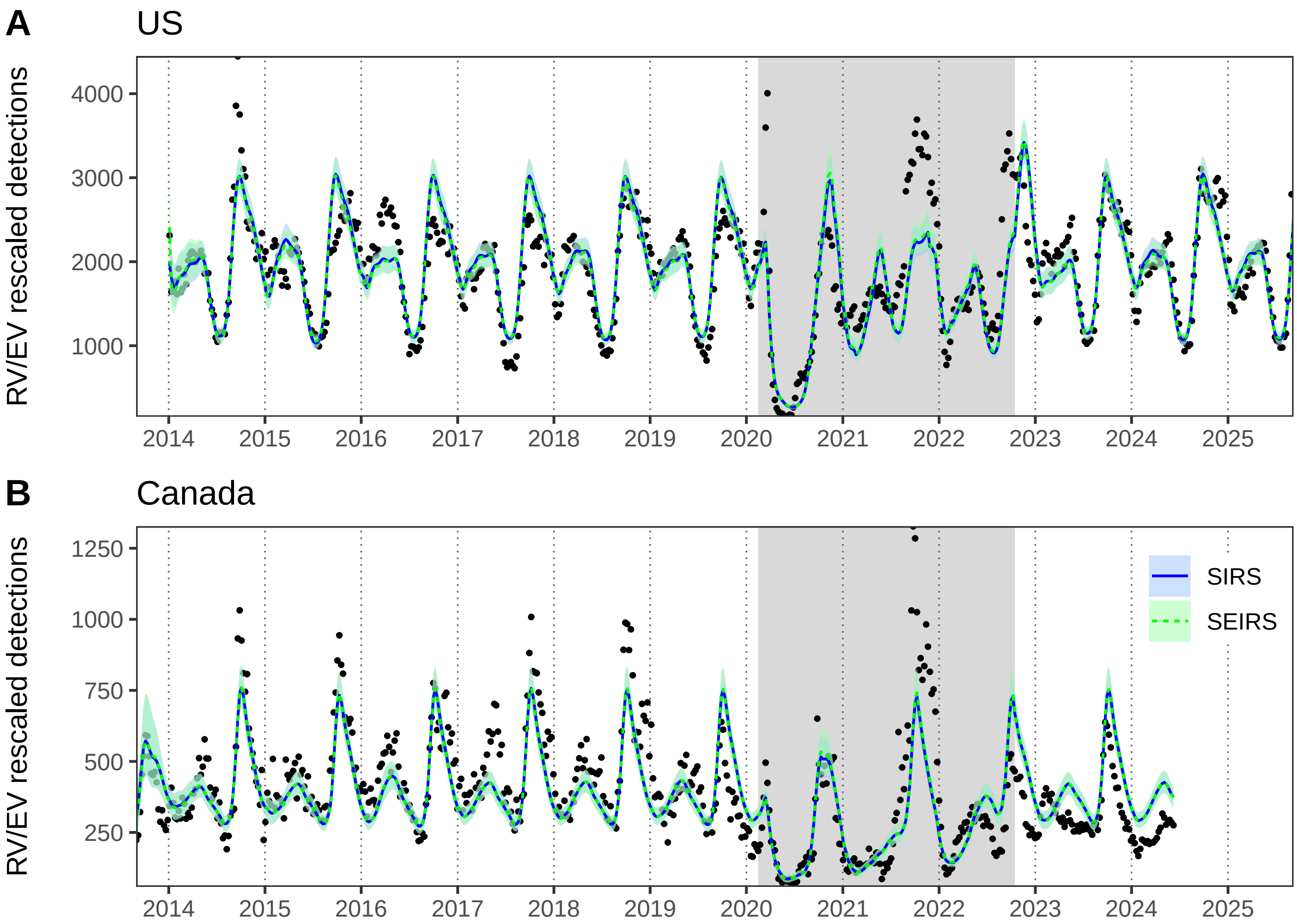

Figure S24: **Comparison of SIRS and SEIRS model fits at the national level.** Built upon the SIRS model, the SEIRS model includes an exposed (i.e., infected but not yet infectious) stage $E$. Because the incubation period (i.e., mean sojourn time in E) is shorter than a week, we used a smaller time step of $\Delta t = 0.25$ week to fit the discretized SEIRS model. We fixed the transition rate from $E$ to $I$ to 5 week$^{-1}$ ($\approx$ 2.45 days for incubation) and the recovery rate to 0.975 week$^{-1}$ ($\approx$ 8 days for recovery) [6]. We fitted both models to rescaled national RV/EV detections (points) in (A) the US and (B) Canada. Lines represent median values of posterior distributions and envelopes represent 95% CrIs; grey backgrounds indicate the pandemic period. For both models, the estimated viral interference parameter $\hat{\phi}$ was not significantly different from 0.

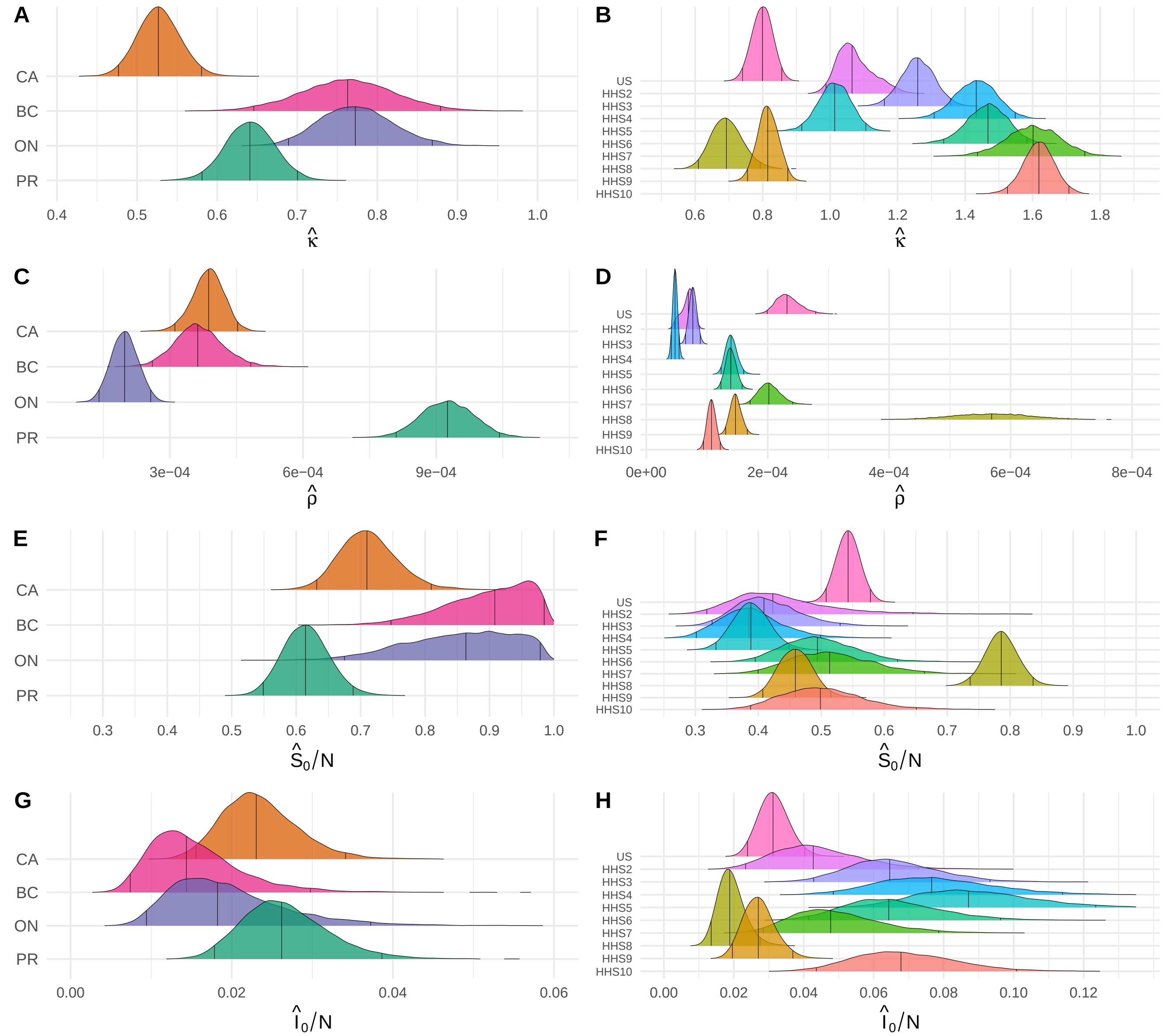

Figure S25: **Posterior density distributions of the remaining estimated parameters.** We show posterior distributions obtained with the model with no viral interaction ($\phi = 0$) using time series from (first column) Canada and (second column) the US. Vertical black lines represent 2.5%, 50% (median) and 97.5% quantiles, respectively. See **Fig. 5** in the main text for estimates of RV weekly transmission rates and mean duration of immune protection, and see **Table 1** for notations. CA=Canada (national), BC=British Columbia, ON=Ontario, PR=Prairies.

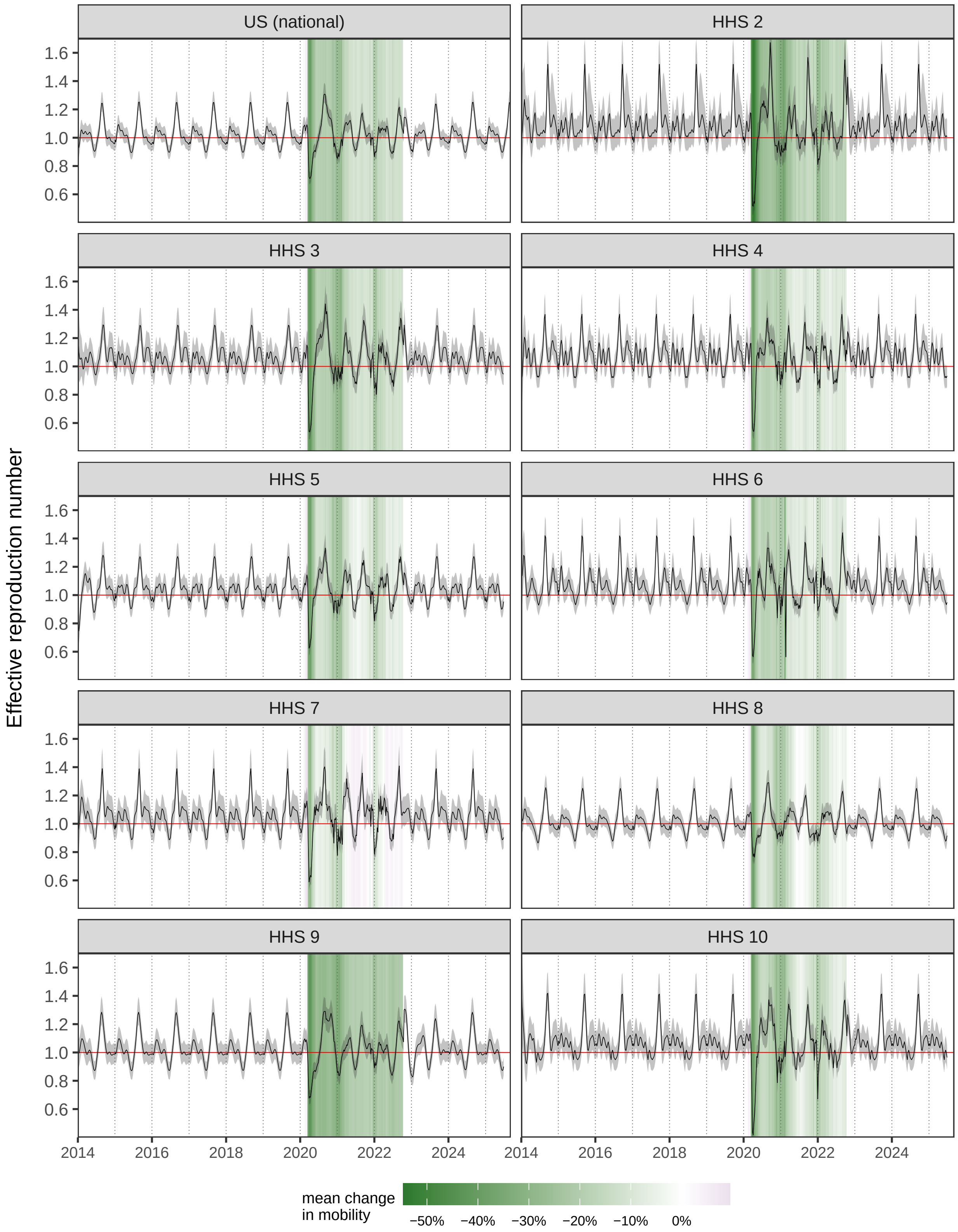

Figure S26: **Dynamics of the estimated effective reproduction number in the US.** We only show results obtained with the model with no viral interaction ($\phi = 0$). Median (black lines) and 95% CrIs (shaded envelopes) of RV/EV effective reproduction number $\mathcal{R}$ were computed from parameter posterior distributions. The epidemic grows as soon as $\mathcal{R} > 1$ (horizontal red line). Mean change in mobility (colored background) during the COVID-19 pandemic were computed from Google COVID-19 Community Mobility Reports.

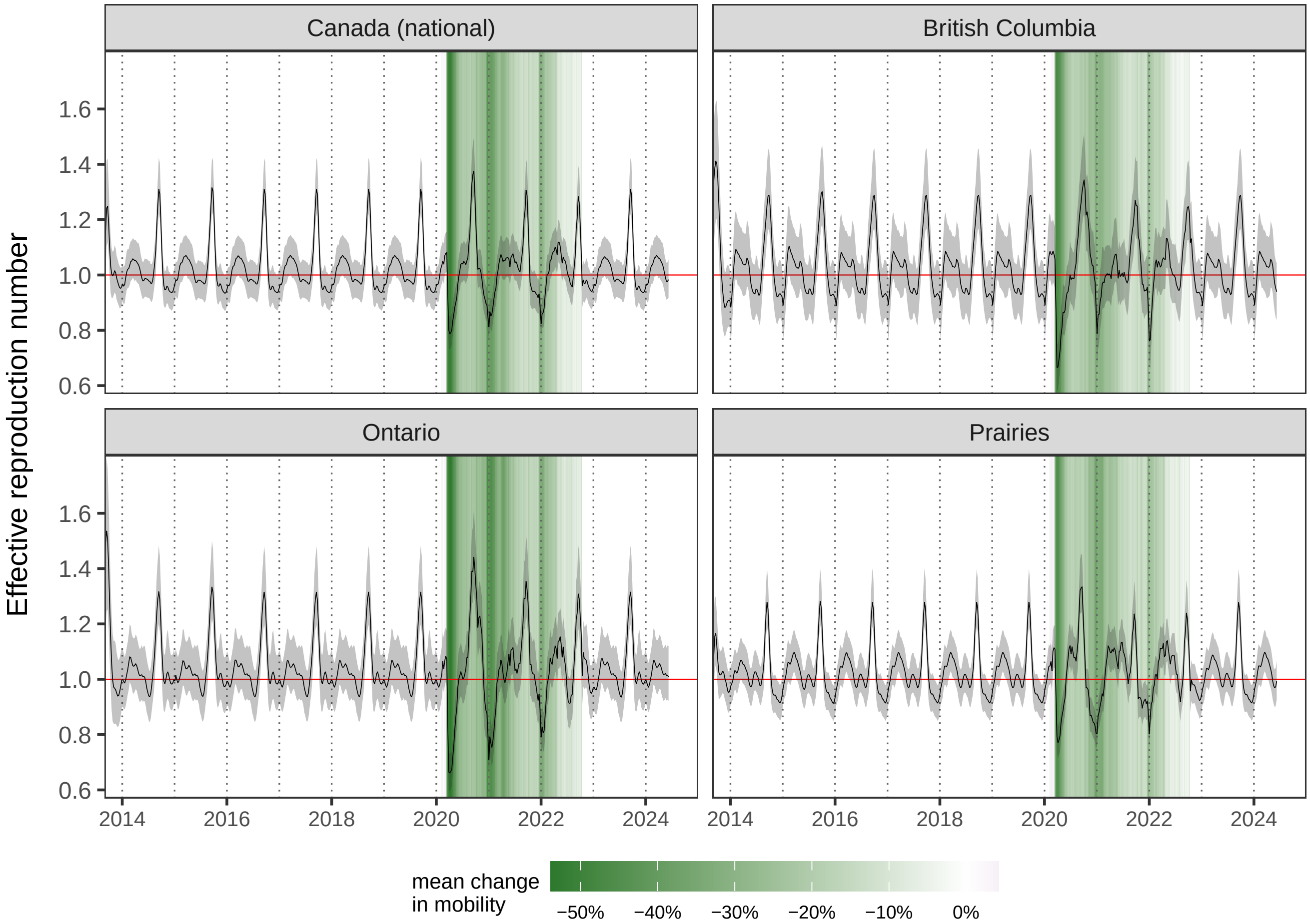

Figure S27: **Dynamics of the estimated effective reproduction number in Canada.** We only show results obtained with the model with no viral interaction ($\phi = 0$). Median (black lines) and 95% CrIs (shaded envelopes) of RV/EV effective reproduction number $\mathcal{R}$ were computed from parameter posterior distributions. The epidemic grows as soon as $\mathcal{R} > 1$ (horizontal red line). Mean change in mobility (colored background) during the COVID-19 pandemic were computed from Google COVID-19 Community Mobility Reports.

### S5 Supplementary tables

Table S1: **Parameters for the simulation study.** Virus parameters are based on existing literature and/or to reproduce the observed epidemiological characteristics of RV and IAV. With these parameter values, IAV exhibits a single annual peak during winter, whereas RV shows two seasonal peaks in fall and spring. The basic reproduction number ($\mathcal{R}_0 = \beta/(\gamma + \mu)$) ranges between 1.53 and 2.17 for RV, and between 1.33 and 2.0 for IAV, which is consistent with [6–8]. The mean duration of infectiousness is set to $1/\gamma_{\text{IAV}}$ =5 days for IAV [6, 9], and $1/\gamma$ =8 days for RV [6]. The mean duration of IAV immunity is set to $1/\omega_{\text{IAV}}$ =40 weeks, reflecting temporary protection over approximately only one influenza season due to antigenic drift. For the transmission rate of RV in scenario 2, we substitute $\beta(t)$ by $\beta(t)/(1 + \phi I^{\star}_{\text{IAV}}(t)/N)$ using values from scenario 1 (except for $\phi$) and where $\star$ denotes the endemic attractor, so that the force of infection of RV mimics the one in 1 in the absence of perturbation.

| Parameter | Value | Unit |
|---|---|---|
| $N$ | $5 \times 10^6$ | individuals |
| $\mu$ | $1/(80 \times 52)$ | week$^{-1}$ |
| $\sigma$ | 0.25 | – |
| **RV (scenario 1)** | | |
| $\beta(t)$ | $\beta_0 \left[1 + a_1 \cos(4\pi (t/52 - \delta_1)) + a_2 \cos(2\pi (t/52 - (\delta_1 + \delta_2)))\right]$ | week$^{-1}$ |
| $\beta_0$ | 1.6 | week$^{-1}$ |
| $a_1$ | 0.1 | – |
| $a_2$ | 0.1 | – |
| $\delta_1$ | 0.35 | year |
| $\delta_2$ | 0.6 | year |
| $\phi$ | 0 | – |
| **RV (scenario 2)** | | |
| $\beta(t)$ | See caption | week$^{-1}$ |
| $\phi$ | $-5$ | – |
| **RV (scenario 3)** | | |
| $\beta(t)$ | $\beta_0 \left[1 + a \cos(2\pi (t/52 - \delta))\right]$ | week$^{-1}$ |
| $\beta_0$ | 1.6 | week$^{-1}$ |
| $a$ | 0.2 | – |
| $\delta$ | 0 | year |
| $\phi$ | $-5$ | – |
| **RV (all scenarios)** | | |
| $\gamma$ | 7/8 | week$^{-1}$ |
| $\omega$ | 1/10 | week$^{-1}$ |
| $\kappa$ | 0.85 | – |
| $\rho$ | $1 \times 10^{-3}$ | – |
| **IAV** | | |
| $\beta_{\text{IAV}}(t)$ | $\beta_{0\,\text{IAV}} \left(1 + a_{\text{IAV}} \cos(2\pi (t/52 - \delta_{\text{IAV}}))\right)$ | week$^{-1}$ |
| $\beta_{0\,\text{IAV}}$ | 2.5 | week$^{-1}$ |
| $a_{\text{IAV}}$ | 0.2 | – |
| $\delta_{\text{IAV}}$ | 0.9 | year |
| $\gamma_{\text{IAV}}$ | 1.4 | week$^{-1}$ |
| $\omega_{\text{IAV}}$ | 1/40 | week$^{-1}$ |
| $\kappa_{\text{IAV}}$ | 1 | – |
| $\rho_{\text{IAV}}$ | $2 \times 10^{-3}$ | – |

Table S2: **Priors.** (Half-)$\mathcal{N}\left(\mu,\, \sigma^2\right)$ refers to the (half-)normal distribution with mean $\mu$ and variance $\sigma^2$, and $\mathcal{U}\,(a,\, b)$ refers to the uniform distribution between $a$ and $b$.

| **Term** | **Prior** | **Support/bounds** |
|---|---|---|
| $\ln\left(\beta_0\right)$ | $\mathcal{N}\left(\ln(2.5),\, 0.25^2\right)$ | $\mathbb{R}$ |
| $z_i$ | $\mathcal{N}\,(0,\, 1)$ | $\mathbb{R}$ |
| $\sigma_\beta$ | Half-$\mathcal{N}\left(0,\, 0.2^2\right)$ | $[0,\, +\infty[$ |
| $\kappa$ | $\mathcal{U}\left(0,\, -1/\min\left(c(t)\right)\right)$ | $\left[0,\, -1/\min\left(c(t)\right)\right]$ |
| $\phi$ | $\mathcal{N}\left(0,\, 0.2^2\right)$ | $[-1,\, 1]$ |
| $\omega$ | Half-$\mathcal{N}\left(0,\, 0.2^2\right)$ | $[0,\, 1]$ |
| $\rho$ | Beta(1, 99) | $[0,\, 0.01]$ |
| $S(0)/N$ | Beta(7, 1) | $[0,\, 1]$ |
| $I(0)/N$ | Beta(2, 98) | $[0,\, 1 - S(0)/N]$ |
| $\sigma$ | Half-$\mathcal{N}\left(0,\, 0.5^2\right)$ | $[0,\, +\infty[$ |

Table S3: **Parameter estimates.** Median and 95% CrIs of posterior distributions obtained with the model without viral interaction (see distributions in **Fig. S25**). We plot estimations of seasonal transmission profiles $\beta(t)$ and mean durations of immune protection $\Omega$ in **Fig. 5**.

| **Location** | $\boldsymbol{\Omega}$ (weeks) | $\boldsymbol{\kappa}$ | $\boldsymbol{\rho}$ ($\times10^{-4}$) | $\boldsymbol{S_0/N}$ | $\boldsymbol{I_0/N}$ ($\times10^{-2}$) |
|---|---|---|---|---|---|
| **United States** | | | | | |
| National | 14.5 [12.1, 18.3] | 0.80 [0.74, 0.86] | 2.32 [1.99, 2.79] | 0.54 [0.51, 0.58] | 3.12 [2.39, 4.03] |
| HHS 2 | 5.64 [3.50, 7.28] | 1.07 [0.99, 1.19] | 0.69 [0.47, 0.84] | 0.42 [0.32, 0.65] | 4.27 [2.34, 7.27] |
| HHS 3 | 8.56 [6.87, 10.4] | 1.26 [1.16, 1.36] | 0.77 [0.65, 0.89] | 0.41 [0.33, 0.53] | 6.46 [4.28, 9.32] |
| HHS 4 | 6.06 [4.94, 7.29] | 1.43 [1.31, 1.55] | 0.48 [0.41, 0.54] | 0.39 [0.30, 0.50] | 7.66 [4.84, 11.5] |
| HHS 5 | 7.27 [6.19, 8.58] | 1.01 [0.92, 1.11] | 1.40 [1.23, 1.60] | 0.39 [0.33, 0.45] | 8.73 [6.00, 12.6] |
| HHS 6 | 5.43 [4.54, 6.42] | 1.47 [1.34, 1.58] | 1.39 [1.23, 1.58] | 0.49 [0.40, 0.62] | 6.43 [4.14, 9.63] |
| HHS 7 | 4.50 [3.56, 5.66] | 1.60 [1.44, 1.75] | 2.02 [1.71, 2.41] | 0.51 [0.40, 0.66] | 4.77 [2.84, 7.85] |
| HHS 8 | 10.6 [8.19, 13.24] | 0.69 [0.61, 0.79] | 5.69 [4.43, 6.94] | 0.79 [0.74, 0.84] | 1.89 [1.35, 2.75] |
| HHS 9 | 20.0 [16.41, 24.43] | 0.82 [0.76, 0.87] | 1.47 [1.31, 1.66] | 0.46 [0.41, 0.52] | 2.70 [1.96, 3.69] |
| HHS 10 | 7.50 [6.26, 8.92] | 1.62 [1.53, 1.71] | 1.07 [0.94, 1.22] | 0.50 [0.39, 0.65] | 6.78 [4.36, 10.1] |
| **Canada** | | | | | |
| National | 8.68 [6.73, 10.56] | 0.53 [0.48, 0.58] | 3.87 [3.11, 4.53] | 0.71 [0.63, 0.81] | 2.30 [1.56, 3.41] |
| British Columbia | 8.03 [5.45, 10.98] | 0.76 [0.65, 0.88] | 3.63 [2.60, 4.82] | 0.91 [0.75, 0.98] | 1.44 [0.74, 2.98] |
| Ontario | 6.98 [4.68, 9.76] | 0.77 [0.69, 0.87] | 1.98 [1.40, 2.57] | 0.86 [0.67, 0.98] | 1.82 [0.94, 3.73] |
| Prairies | 10.3 [8.40, 12.42] | 0.64 [0.58, 0.70] | 9.25 [8.10, 10.4] | 0.61 [0.55, 0.69] | 2.62 [1.79, 3.86] |
